# Hemochromatosis Neural Archetype Reveals Iron Disruption in Motor Circuits

**DOI:** 10.1101/2022.10.22.22281386

**Authors:** Robert Loughnan, Jonathan Ahern, Mary Boyle, Terry L. Jernigan, Donald J. Hagler, John R. Iversen, Oleksandr Frei, Diana M. Smith, Ole Andreassen, Noah Zaitlen, Leo Sugrue, Wesley K. Thompson, Anders Dale, Andrew J. Schork, Chun Chieh Fan

## Abstract

Our understanding of brain iron regulation and its disruption in disease is currently lacking. We previously found that motor circuitry is susceptible to the cumulative neurotoxic effects of excessive iron, leading to the manifestation of Parkinson’s disease. However, beyond a few well-known genes involved in peripheral iron metabolism, the underlying molecular mechanisms regulating central iron levels remain unclear. To bridge this gap, we generated scores in neurotypical individuals based on the archetypal brain iron accumulation observed in magnetic resonance imaging scans of individuals who exhibit excessive absorption of dietary iron and hemochromatosis risk. Genome-wide analysis, using common-variant SNP-array and rare-variant exome data, demonstrated this archetypal brain iron accumulation pattern is highly heritable, revealing both known and novel loci associated with iron homeostasis, and causally driven by peripheral iron levels. Our score predicted abnormalities in gait and revealed a U-shape relationship with PD risk - identifying a group of individuals with a 3-fold increased risk for this disorder. Taken together these results establish a hormetic relationship between brain iron and PD risk, in which central iron levels are strongly determined by genetics via peripheral iron. We believe this framework combining forward and reverse genetics represents a powerful new study design to understand genomic drivers underlying high dimensional phenotypes.

## Introduction

Iron is an essential element required for the body to function properly. The role of iron in numerous biochemical reactions stems from its dual function as an electron mediator, serving as both a donor and acceptor, making it indispensable for life. In the brain, iron is particularly important for neuronal development, myelination and synthesis of neurotransmitters such as dopamine^1^. For these reasons during early life, there is a substantial demand for iron, and not meeting this requirement can lead to iron deficiency anemia^2^. More broadly iron deficiency in early life can have profound and long-term effects on motor skills, cognitive function, and socio-emotional behavior^3^, underscoring the critical role of iron in healthy development. Conversely, too much iron can be toxic and dangerous also. Through the generation of reactive oxygen species and iron dependent cell death, known as ferroptosis^4,5^, iron overload can cause widespread damage to healthy tissue throughout the body^6^. As iron depletion and overload can both cause substantial damage, the body must orchestrate a careful balancing act to regulate iron levels. Although we have a good understanding of how this is achieved systemically through the interaction of hepcidin and ferroportin^7,8^, our understanding of mechanisms controlling brain iron remains incomplete^9^. Elucidating these processes is of importance to gain insight into metabolic homeostasis in the brain.

Magnetic Resonance Imaging (MRI) provides a noninvasive method to accurately estimate iron concentrations in the brain^10^. This has enabled researchers to study trajectories of central iron across the lifespan uncovering a pattern of iron accumulation throughout regions of the basal ganglia in later life^11^ - possibly contributing to senescence. MRI and postmortem studies have also implicated iron dysregulation in numerous neurodegenerative diseases^1^, most notably in movement disorders such as Parkinson’s Disease (PD)^12^ and Neurodegeneration with Brain Iron Accumulation (NBIA)^13^. These findings have led researchers to attempt to use iron as a therapeutic target for PD and related disorders through the use of iron chelators^14^ which bind to and remove excess iron in the blood. Clinical trials of iron chelators have shown promise for some patients with movement disorders while exacerbating symptoms in others.^15–17^ Our inability to explain this heterogeneity in treatment outcomes points to a poor understanding of iron dyshomeostasis occurring in movement disorders. Recent work has aimed to understand the genetic determinants of regional iron accumulation in the brain^18^, however this has led to little further insight into abhorrent regulatory mechanisms in disease. There is therefore a critical need to define biologically grounded phenotypes when studying brain imaging signals^19^.

Here, we adopt a pioneering approach to study brain iron, its genetic determinants and neurodegenerative manifestations. In this approach we leverage the regional brain iron accumulation associated with the most prevalent genetic risk factor for excessive iron absorption in the gut, known as C282Y homozygosity. This genotype is responsible for the majority of hereditary hemochromatosis cases^20^ and in recent work we demonstrated that it exhibits an MRI pattern showcasing substantial localized iron accumulation in the brain’s motor circuits, specifically the basal ganglia and cerebellum^21^. Notably, this genotype also corresponds to a twofold increase in male-specific risk of developing movement disorders. Here we generate a novel brain endophenotype, based on C282Y, which captures a continuum of brain iron dysregulation and is predictive of risk for movement disorders. To operationalize our recent MRI findings of C282Y into an endophenotype, which we refer to as the “Hemochromatosis Brain”, we trained a classifier to predict C282Y homozygosity status from T2-weighted brain MRI scans, a modality sensitive to iron accumulation^10^. We performed this training in a subsample of 960 individuals taken from UK Biobank (UKB). We then deploy this classifier on 35,283 independent MRI images, also from UKB, none of whom are C282Y homozygotes, to generate a PolyVoxel Score (PVS)^22^ for each individual capturing the degree to which they resemble the archetypal Hemochromatosis Brain. The resulting PVS enables us to answer two questions: first, does a neural archetype learnt from the extreme case of iron dysregulation inform us about the genetic drivers of iron homeostasis of the human brain more generally? Second, does the degree of brain iron buildup, as encapsulated by the Hemochromatosis Brain, predict risk for movement disorders? This work is an important step in understanding the genetic determinants of brain iron dysregulation and in identifying putative iron-related subgroups of movement disorders.

## Results

### Learning from the Hemochromatosis Brain

Figure 1 presents a graphical workflow of the analysis performed in this paper, with Supplementary Table 1 giving a demographic breakdown of each subsample. Performing 5-fold cross validation in Subsample A of UKB, consisting of 193 C282Y homozygotes and 767 covariate-matched controls, established that the Hemochromatosis Brain classifier can predict C282Y homozygosity status with high accuracy from T2-Weighted scans (attaining an AUC=0.86). Supplementary Figure 1 shows results of hyperparameter tuning. Figure 1 displays classifier weights and features importance of classifier by brain region (see also Supplementary Figure 2). As expected, univariate statistics show strong resemblance to previous work describing C282Y homozygote effect on T2-weighted signal^33^. Feature importance of posterior effects indicate that the cerebellum, thalamus, putamen and caudate have the largest contributions to the PVS.

**Figure 1:**
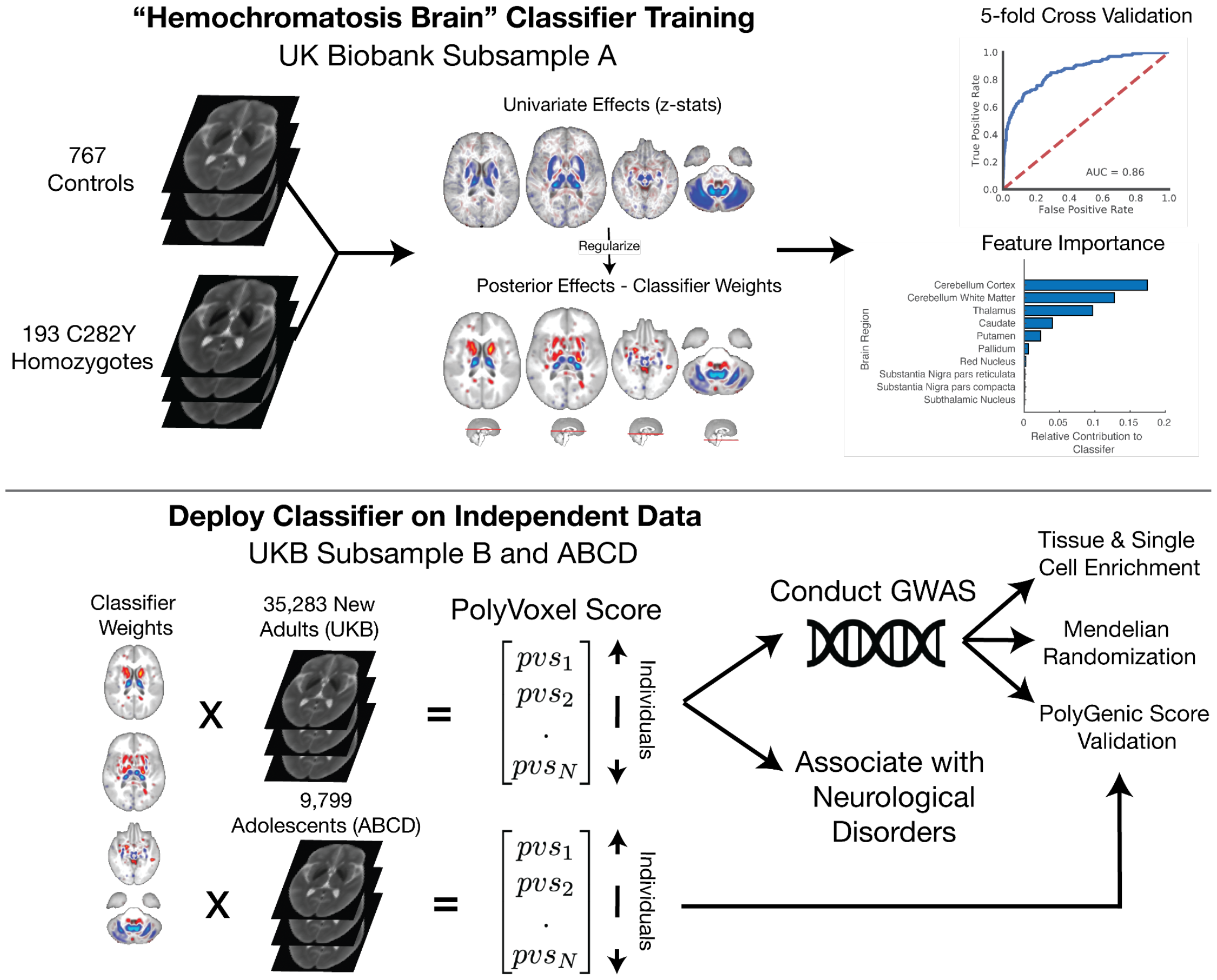
Overview of study design. Top panel: Hemochromatosis Brain Classifier training in Subsample A of UKB to differentiate controls from C282Y homozygotes from T2-weighted MRI scans, with univariate and regularized classifier weights, classifier performance (ROC) and feature importance by brain region. Lower panel: deploy classifier in Subsample B (UKB) and ABCD Study^®^ to generate PolyVoxel Score (PVS) capturing Hemochromatosis Brain liability. In Subsample B we then conduct a GWAS to find variants associated with this PVS liability scale. Using these GWAS results we perform tissue and single cell enrichment analysis, mendelian randomization and polygenic score validation against the PVS generated from the ABCD Study. Finally, we test this PVS against neurological disorders within UKB.

### Evaluating the Validity of the Hemochromatosis Brain with Genetic Analyses

We then sought to evaluate the validity of our scoring system by conducting a series of genetic analyses based on the Hemochromatosis Brain PVS in an independent set of individuals from UKB. First, we performed a Genome-Wide Association Study (GWAS) with the PVS in Subsample B as the phenotype of interest. For discovery we restricted this analysis to a single homogenous ancestry group, and used the remaining individuals for validation. This resulted in a discovery sample size of 30,709 European ancestry individuals and 4,608 non European ancestry individuals for replication. This analysis revealed 42 genome-wide significant loci (13 of these being novel for any reported trait) associated with the Hemochromatosis Brain phenotype and high heritability: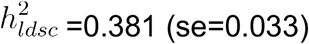 - see Figure 2A and Supplementary Figures 3 and 4. We found this heritability was substantially higher than the heritability estimated for peripheral blood iron markers (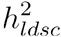 between 0.029-0.056) - see extended data tables. Among the top hits we found many that are known regulators of iron homeostasis: for example, rs6794370 (TF; p=2.43×10^−81^) transferrin is the principal glycoprotein responsible for mediating transport of iron through blood plasma; rs1800562 (HFE; p=6.93×10^−75^) is the same C282Y mutation on which the classifier was trained (here rediscovered with the heterozygotes); rs2413450 (TMPRSS6, p=2.52×10^−50^), TMPRSS6 is part of production signaling pathway of hepcidin, the key hormonal regulator of iron absorption in humans; rs13107325 (p=3.66×10^−41^) and rs12304921 (p=5.05×10^−21^) are linked respectively to metal transporters ZIP8 and DMT1, the latter being a gene highly expressed in the gut and previously linked with microcytic anemia with iron overload^23,24^.

**Figure 2:**
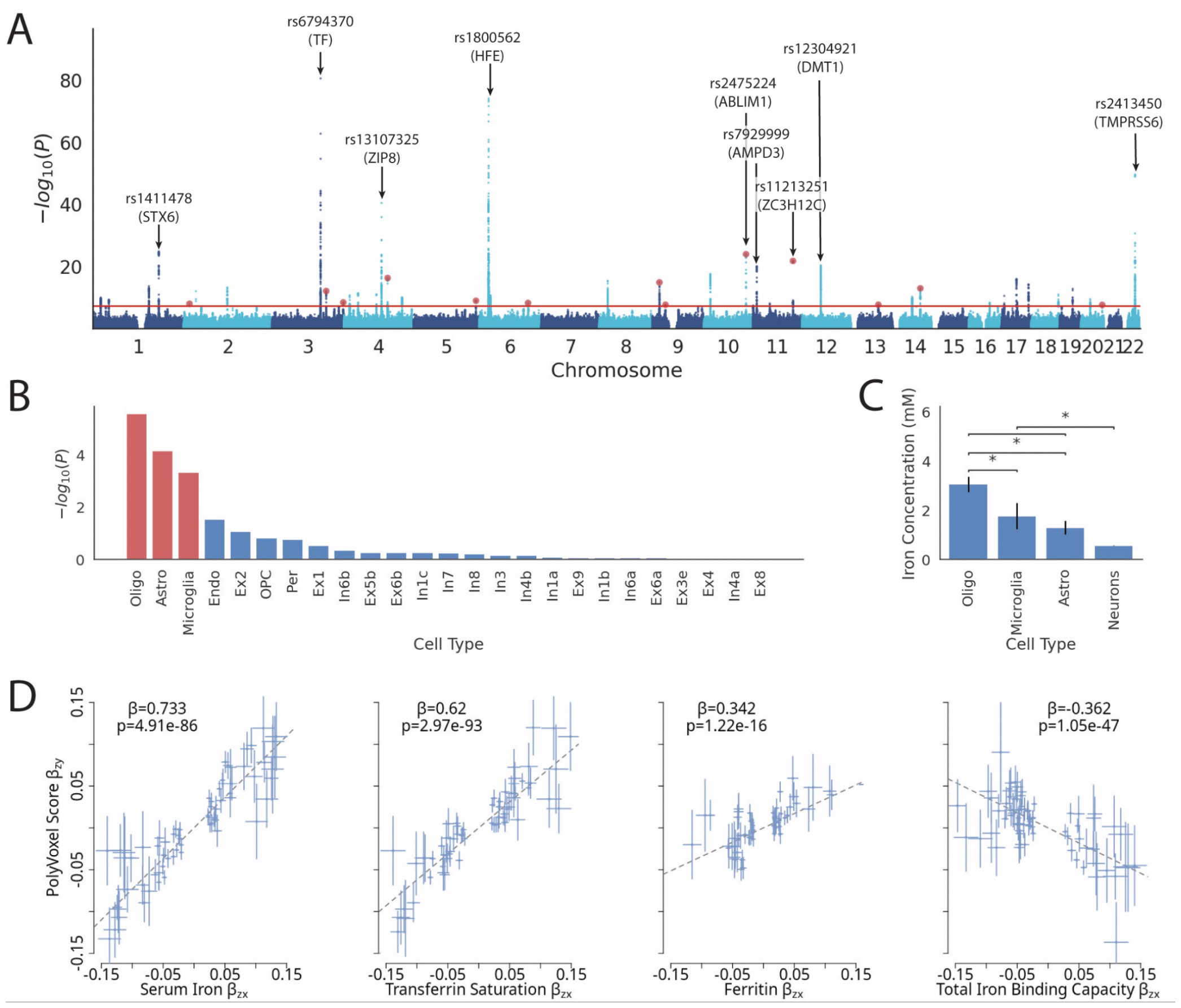
Results of GWAS on PVS of Hemochromatosis Brain in Subsample B. Panel A: Manhattan plot and QQ plot of GWAS result with peaks annotated – a total of 42 loci are discovered, see supplementary data tables for full list. Red dots indicate 13 novel loci. Panel B: Brain cell type enrichment of GWAS signal using FUMA, red bars indicate FDR significantly enriched cell types. Panel C: Iron concentration (measured by using x-ray spectrometry) of cell types in rat brain with permissions from Reinert, A et. al.^34^. Panel D: Mendelian Randomization (MR) results, using GSMR, to quantify strength of causal relationship between peripheral blood iron markers and brain iron accumulation as measured by Hemochromatosis Brain PVS. Each plot shows results of conditioning on peripheral blood iron markers as exposure and PVS as outcome, reverse direction GSMR results are shown in Supplementary Figure 6.

Among the 13 novel loci we discovered was rs73210039 (4.47×10^−9^) proximal to TFRC, transferrin receptor 1, a gene known to play a critical role in cellular iron uptake with a variable expression pattern across the brain^25^. This locus was not implicated in recent GWAS of peripheral iron markers^26^, however a nearby marker, rs3817672, (also proximal to TFRC) was found to be associated with serum transferrin and total iron binding capacity. This indicates that these polymorphisms appear to have differential effects on central vs systemic iron through TFRC. Another novel locus, rs62560298 (p=1.64×10^−15^), proximal to DENND4C, is a member of the retromer complex family that deals with recycling of metals in the body, which has been hypothesized to be implicated in neurodegeneration such as PD^27^. A further novel locus was detected at rs112593750 (p=9.82×10^−13^) intronic to CPHL1P, a pseudogene whose functioning homolog is hephaestin - a gene necessary for effective iron transport in the intestines and which has been found to mutated in sex linked anemia^28^.

We found our discovered SNPs showed greatest overlap with previously studied traits relating to iron/red blood cells, brain/cognition and non iron blood markers (Supplementary Figure 5), with only one SNP (rs13107325) previously linked with PD^29^. Analysis of rare genetic variants using exome burden testing revealed two genes associated with the PVS: TF (p=5.21×10^−8^) and OR1J4 (p=1.35×10^−6^). See extended data tables for full summary of GWAS results.

Using LDSC^30^ on common variant GWAS results we found the Hemochromatosis Brain was negatively genetically correlated only with intracranial volume (*r*_*g*_ =-0.21, z=-5.19, =1.7×10^−6^) and no other traits tested (Supplementary Figure 6). We additionally performed S-LDSC (stratified-LDSC) across 489 tissue-specific chromatin-based annotations across the body from peaks for six epigenetic marks^31^. Results revealed the hippocampus (H3K27ac, *p*_*fdr*_ =0.014 and H3K4me1, *p*_*fdr*_ =0.025) and substantia nigra (H3k27ac, *p*_*fdr*_ =0.036) displayed a significantly enriched signal. Performing cell type enrichment using FUMA^32^ of Hemochromatosis Brain GWAS using cell types defined from PsychENCODE^33^ displayed an enrichment for glial cells: oligodendrocytes (*p*_*fdr*_ =6.62×10^−5^), astrocytes (*p*_*fdr*_ =1.7×10^−3^) and microglia (*p*_*fdr*_ =1.8×10^−2^) and somewhat surprisingly no enrichment for neuronal cells– see Figure 2B. This pattern of enrichment exhibits a similar pattern followed by iron concentrations calculated using x-ray spectroscopy^34^ – displayed in Figure 2C for comparison. Mendelian randomization^35^ (MR) results between peripheral blood iron markers^26^ and the PVS revealed evidence for a strong causal relationship leading from serum iron (*β*_*std*_ =0.73, p=4.91×10^−86^) and transferrin saturation (*β*_*std*_ =0.62, p=2.97×10^−93^) to brain iron accumulation (Figure 2D) with no evidence for the reverse relationship (Supplementary Figure 7). We additionally found significant, although weaker, MR associations of total iron binding capacity (*β*_*std*_ =-0.36, p=1.05×10^−47^) and ferritin (*β*_*std*_ =0.342, p=1.22×10^−16^) to brain iron accumulation as captured by the PVS, with again no evidence for the reverse direction (Supplementary Figure 7). Taken together these indicate evidence for a causal link of peripheral serum iron and transferrin saturation leading to variability in brain iron accumulation measured by the PVS. Interestingly, despite significant phenotypic associations between PVS and PD, we did not find any significant MR associations between PVS and PD, or peripheral blood iron markers and PD (Supplemental Materials, Supplementary Figure 7).

Within our GWAS discovery we found 281 independent significant SNPs (r_LD_<0.6). We found strong evidence of replication of these SNPs within our validation cohort of 4,608 non-European ancestry individuals from Subsample B (Supplementary Table 2) with high correlation of *β* estimates between these two cohorts (r=0.90, p<1.28×10^−103^) - see Supplementary Figure 8. Sign concordance between discovery and replication cohorts was high for independent significant SNPs (94.6%, p=1.0×10^−59^) and for 13 novel loci (92.3%, p=0.003). To test the generalization of our GWAS signal we evaluated the performance of a PolyGenic Score (PGS) to predict a PVS generated in 9,799 individuals from the ABCD Study^®^ (aged 8-14 years old, i.e. more than 50 years younger than our training sample). Evaluating performance in separate ancestry strata we found the PGS significantly predicted the PVS (EUR: r^2^=0.018, z=12.67, p=8.01×10^−37^; AFR: r^2^=0.0086, z=2.73, p=6.33×10^−3^, MIX: r^2^=0.0069, z=5.47, p=4.41×10^−8^). To further understand PVS changes across the lifespan, within ABCD (full sample) and UKB (subsample B), we visualize the effect of age and sex - shown in Supplementary Figure 9. Despite the much smaller age range of ABCD vs UKB, we find a notably larger age effect in ABCD (z=53.2, r^2^=0.16, p<10^−100^) than in UKB (z=-4.00, r^2^=4.46×10^−4^, p=7.29×10^−5^) with females appearing to be phase advanced in the adolescent age range (z=27.0, r^2^=0.048, p<10^−100^). In sum, these results demonstrate that the genetics underlying the Hemochromatosis Brain PVS are detectable across the lifespan (8-80 years old) and that there are notable age-related changes in early adolescence.

## Iron Homeostasis in Human Brain and Neurological Disorders

After establishing the genetic determinants of the Hemochromatosis Brain, we next examined if this novel measure was associated with any neurological disorders including movement disorders and abnormalities of gait. We found the PVS in Subample B (containing no C282Y homozygotes) significantly predicted reduced risk for PD (OR=0.74, Z=-4.98, p=6.42×10^−7^) and Abnormalities of Gait and Mobility (OR=0.89, Z=-3.28, p=0.001) – see Figure 3A and Supplementary Table 4. The associations were specific to movement and gait disorders, as we did not find evidence of a PVS association with diagnoses of any other neurological disorders tested. Performing quantile weighted regression to predict PD status in Subsample C (Figure 3B and Supplementary Table 5) we see higher risk at the two ends of the estimated brain iron concentration spectrum: with the 1^st^ PVS quantile (OR=3.19, Z=5.61, p=2.06×10^−8^) and C282Y homozygotes (OR=2.40, Z=3.34, p=8.41×10^−4^). For Figure 3B mean iron concentrations for brain areas used to calculate PVS were estimated from T2^*^ imaging for each PVS quantile and C282Y homozygotes separately to plot values on the x axis. In contrast to C282Y homozygotes, who exhibited excessive iron accumulation and twofold increased risk for PD, the PVS exhibited an inverse relationship between brain iron concentration and PD risk, indicating the critical role of iron homeostasis in maintaining the function of motor circuits. Note for Figure 3B, although there appears to be a U-shape relationship, a far greater proportion of individuals are present in the iron deplete range than in the iron overload range; each blue point represents a quantile (∼25% of the cohort) while the orange point represents ∼0.6% of the cohort. No such U-shape relationship was observed for Abnormalities of Gait and Mobility risk (Supplementary Figure 10).

**Figure 3:**
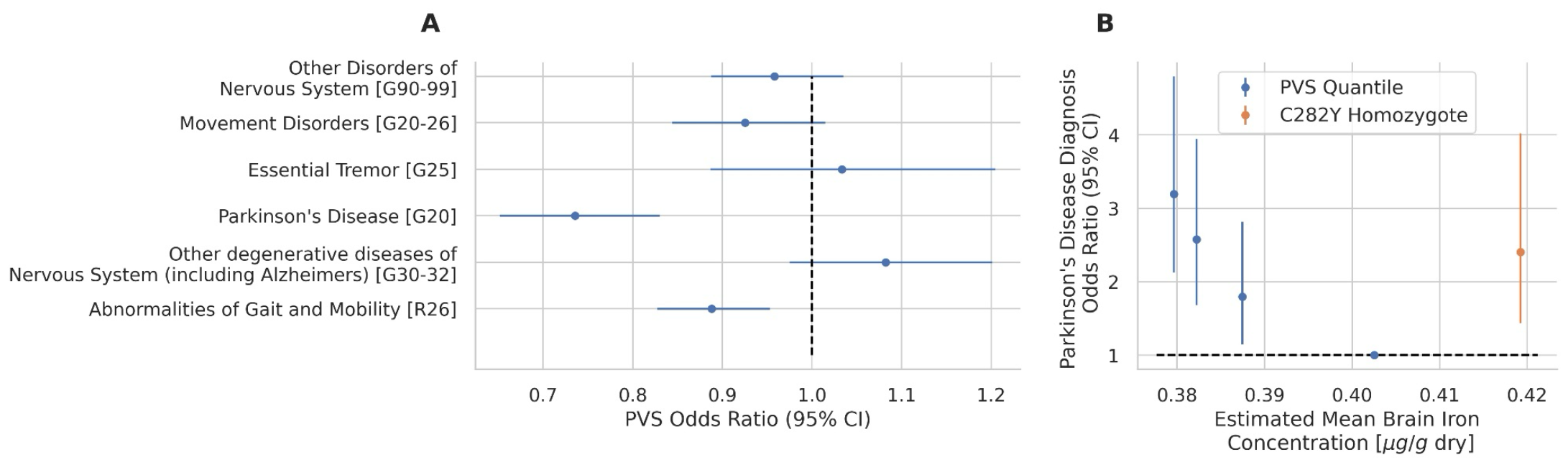
Weighted logistic regression using PolyVoxel Score (PVS) to predict neurological diagnoses in Subsample B. Inverse Probability Weights (IPW) for each disorder can be found in Supplementary Table 4. Panel A: PVS odds ratio predicting each neurological disorder. Panel B: PVS quantile weighted regression to predict PD in Subsample C, each point represents a categorical factor indicating one of four PVS quantile (blue) or C282Y homozygosity (orange). IPW of 3.89 was used for PD cases in imaging sample (blue) - see methods. Regression (y-axis) was performed using PVS from T2-W, x-axis is an estimate of mean brain iron concentration using T2* imaging for each group.

## Discussion

Here, we have presented a continuum of iron homeostasis localized to motor circuits of the brain which is predictive of risk for movement disorders. We have shown that this dysregulation spectrum is strongly associated with genetic variation 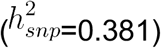, discovering both novel variants and variants proximal to known key regulators of iron homeostasis. Furthermore, the signal was enriched in gene expression patterns of glial cells – a cell group known to have highest concentrations of iron in the brain^34^. Additionally, MR analysis showed that iron markers in peripheral blood have a strong influence on brain iron accumulation differences. Yet the PD risk profile exhibited a U shape, indicating iron homeostasis is more relevant than absolute iron concentration. These results outline the genetic drivers, mediated through glial cells, influencing iron regulation in the brain and the putative consequences of disrupting this system. Taken together, these results indicate subgroups of PD that relate to the hormetic nature of iron in the brain, in which both depleted and excess levels can impart toxic effects, likely via different mechanisms ^4,36,37^.

The results from this analysis validate an established relationship between iron and movement disorders, while providing a potential novel biomarker for PD risk. Previous *post mortem* and *in vivo* imaging studies have found higher iron deposition in substantia nigra, putamen and red nucleus^38^; these regions have small but non-zero weights contributing to the Hemochromatosis Brain classifier (Supplementary Figure 2). Of note, there is a paucity of *in vivo* imaging results of PD results reporting on the cerebellum^38^ - a region which displays the largest contribution to the PVS and which has been shown to have lower iron levels in post mortem PD brain samples^38^, consistent with our findings. Additionally, deficits in iron have also been associated with movement disorders, with anemic patients and male blood donors being more likely to suffer from PD^39,40^. Furthermore, a class of rare genetic mutations result in a disease called Neurodegeneration with Brain Iron Accumulation (NBIA) which presents with a stereotyped pattern of iron accumulation in the basal ganglia similar to the univariate pattern observed in the Hemochromatosis Brain. The clinical presentation of this disorder is of dystonia and other Parkinsonism symptoms. Moreover, in the context of the MTMP model, which is the most extensively researched animal model of Parkinson’s disease (PD), it seems that ferroptosis, a form of cell death that relies on iron, has emerged as a key mechanism responsible for causing the motor symptoms associated with movement disorders^4^. The work presented in the manuscript once again underscores the link between iron dysregulation in the brain and the risk of developing a movement disorder.

Additionally, we find, to a lesser degree, the PVS predicts risk for Abnormalities of Gait and Mobility. This diagnosis represents a heterogeneous set of disorders. In contrast to IC10:G20 for PD, ICD10:G26 represents symptom-based code and does not directly indicate a specific neurological or MRI finding. Given this heterogeneity it is perhaps unexpected to find any coherent predictive signal from MRI. On the other hand, the PVS has large contributions from the basal ganglia and cerebellum, regions known to be heavily involved in gait^41,42^. Furthermore, prior work has found differences in these regions using diffusion MRI^43^ and midbrain volumetric differences^44^ patients with high level gait disorder. It may prove useful to understand the degree to which our results and those from these previous studies, using other imaging modalities, are capturing overlapping underlying biological processes.

The polygenic signal we uncovered for the Hemochromatosis Brain revealed many known master regulators of iron homeostasis. In addition we uncovered 13 novel loci (not implicated in any previous GWAS) some which were linked to genes involved in metal recycling/transport and one locus proximal to DENND4C - a gene hypothesized to be implicated in neurodegeneration and PD^27^. A question arises that if the Hemochromatosis Brain is so strongly related to iron regulation, why were we able to uncover 13 loci that were not previously reported in well powered GWAS of peripheral iron markers?^26^ Firstly, these 13 loci may be specific to central iron regulation and the emergent pattern of iron accumulation observed regionally in the Hemochromatosis Brain. Indeed, we identified two variants (rs73210039 and rs3817672) - representing distinct loci - which appeared to show differential effects on systemic vs central iron levels. These variants were both proximal to TFR1/TFRC, a gene critical to cellular iron uptake. Secondly, the Hemochromatosis Brain may represent a more stable, longterm, and heritable measure of iron in the body than what is detected through the blood. Indeed, we observe a higher heritability for the Hemochromatosis Brain than peripheral blood markers and repeated measures of peripheral blood markers appear to be unstable over repeated measures.^45^ We hope that future work can further help disentangle the genetic contribution of central vs peripheral iron regulation.

Although epidemiological, imaging and postmortem studies suggest a link between PD and iron, GWAS of the disorder have not uncovered a major contribution from iron^46^. Indeed, in our study we only find one independent significant SNP (rs13107325, p=3.66×10^−41^) overlapping with variants previously associated with PD. We also find that the Hemochromatosis Brain does not display any genetic correlation with the largest previous GWAS of PD (Supplementary Figure 5). Our interpretation of this result is that iron dysregulation may represent one potential contributor to PD risk and without this subtype being identified in current PD GWAS these unmodelled heterogeneous genetic effects lead to null findings for iron related variants. This idea of genetic heterogeneity is thought to explain the modest GWAS results of Major Depression^47^, and we aim to investigate this possibility in future work. Much of the current PD results highlight lysosomal dysfunction as a principal mechanism underlying disease pathopysiology^48^. The contribution of iron dysregulation to PD risk may still converge on lysosomal dysfunction. For example, iron depletion or overload leading to oxidative stress and cell death resulting ultimately in disease presentation; this process could be exacerbated in patients with pre-existing lysosomal dysfunction.

Iron chelators (drugs that facilitate the removal of iron from the body) have recently emerged as a potential therapeutic target for movement disorders with mixed outcomes. In the treatment of NBIA, iron chelation with deferiprone has shown promise in improving symptom progression^15,17^. Conversely, a phase 2 randomized control trial of deferiprone in 372 PD patients was stopped early due to worsening symptom progression from treatment^16^. These results may be consistent with our findings if NBIA and PD represent two ends of the iron dysregulation spectrum. With this view the more rare cases of NBIA result from iron overload thereby benefiting from iron chelation. This is in contrast to PD in the general population which may lie in the more iron-deplete range thereby being exacerbated by chelation therapy. In this interpretation NBIA would align with the risk profile of C282Y homozygotes and PD patients with the lower PVS quantiles in Figure 3B. In support of this view we see the bulk of the UKB imaging sample lies in the iron-deplete range of PD risk with each of the four blue points of Figure 3B representing ∼25% of the sample and C282Y homozygotes representing 0.06%. This indicates that future iron based interventions may benefit from MRI based patient stratification, such as through the use of the Hemochromatosis Brain PVS, enabling targeted treatment.

Single cell enrichment analysis implicated glial, and not neuronal, cells as the principal cell class associated with our Hemochromatosis Brain GWAS results. This is consistent with findings that glial cells, particularly oligodendrocytes, show larger concentrations of iron than neuronal cell types^34,49^ and is also consistent with the role of glial cells’ in maintaining brain homeostasis^50^. In net, it has been estimated that 80% of iron stores in the human brain are within glial cells^51^. Although our results indicate glial cells appear to mediate central disruption of iron homeostasis, the consequence of these disruption may be borne by other cell types. Indeed, recent work has established that microglia are highly responsive to iron perturbations and are drivers of neuronal iron-dependent death^52^. Furthermore dopamine neurons, with a strong dependency on iron for dopamine synthesis^36^ and mitochondrial function^53^, may represent a cell class particularly sensitive to this disruption. In support of this view, studies show that dopaminergic neurons are the cell class to show largest iron concentration differences between healthy and PD brains^51,54^. Further, despite glial cells being widespread across the brain, our analysis of tissue enrichment across the human body found our GWAS signal to be localized only to the substantia nigra and hippocampus. The former being rich in dopaminergic neurons and the primary site of PD pathology^55^, and the latter known to show accelerated atrophy in PD^56^. Taken together cell and tissue specific enrichment analysis indicates iron disruption of glial cells within the substantia nigra and hippocampus lead to variability in iron dysregulation as captured by the PVS. These results further support the view that the Hemochromatosis Brain represents an iron specific endophenotype of relevance to PD.

Analysis revealed a genetic signature of the Hemochromatosis Brain appears to be generalizable across a 50 year age span from adolescence (ABCD) to late adulthood (UK Biobank). Within ABCD we also observed a larger change in PVS scores as a function of age than for UK Biobank (Supplementary Figure 8) - this recapitulates previous findings of brain iron accumulation exhibiting an early exponential pattern with the largest increases observed in the first two decades of life^11^. As others have suggested^57^ this may be indicative of the development of the dopaminergic system, which relies heavily on iron. In addition, along this developmental trajectory we find evidence that females appear to phase advanced when compared to males - a pattern which is observed for other markers during adolescence.^58,59^ It is possible that early life differences in regional brain iron that we detect predisposes individuals to different neurological risk profiles later in life.

An important limitation of the current study is its observational nature. Although, T2-weighted MRI signal is known to be related to iron concentrations^60^, and the GWAS signal we detect appears to implicate iron related genes with encouragingly strong MR results, we cannot conclude that iron specifically is imparting a causal impact on disease risk for PD. It is possible that iron may be serving as a marker for another biological process that is responsible for generating the differential risk for PD. In addition, T2-weighted signal is not solely reduced by higher iron concentrations; processes of edema and gliosis also impact T2-weighted intensity^61^. Moreover, manganese similarly shortens T2-weighted signal and is known to accumulate in the basal ganglia as a result of liver cirrhosis which is common with peripheral iron overload^62^. It is possible that some of the effects captured by the PVS are in fact related to these non-iron processes. Finally, due to correlations between brain regions and the regularization process during training of our PVS some brain regions flip signs (compare univariate statistics and posterior weights in Figure 1 and Supplementary Figure 2). The PVS approach’s strength and weakness lie in its ability to condense a complex signal into one instrumental variable, indicative of a continuum of brain iron dysregulation. Although, in general, higher PVS values indicate more iron accumulation in net, this doesn’t ensure that iron levels in each individual brain region align with this direction.

Here we have showcased a continuum of neural iron dysregulation related to PD risk and which underscores the hormetic nature of this vital element. We have demonstrated that this continuum is strongly influenced by both genetic drivers and peripheral markers of iron. We hope that this dysregulation spectrum may be useful in patient stratification and in understanding heterogeneity in treatment outcomes. To this end we provide weights and code to easily calculate PVS for suitable MRI scans within new datasets - see methods. Our novel approach could be applied more broadly to other archetype learning scenarios, providing a framework for greater biological interpretation of multivariate measures.

## Data Availability

UK Biobank data was accessed under accession number 27412. Summary statistics from GWAS of Hemochromatosis Brain PVS are available for download. Researchers can apply for access to UK Biobank data at https://www.ukbiobank.ac.uk/enable-your-research/apply-for-access. The ABCD data used in this came from [NIMH Data Archive Digital Object Identifier (10.15154/1523041)]. This data is available to approved researchers, find more information at https://abcdstudy.org.

## Acknowledgments

**UK Biobank Acknowledgment:** UK Biobank’s research resource is a major contributor in the advancement of modern medicine and treatment, enabling better understanding of the prevention, diagnosis and treatment of a wide range of serious and life-threatening illnesses – including cancer, heart diseases and stroke. UK Biobank is generously supported by its founding funders the Wellcome Trust and UK Medical Research Council, as well as the Department of Health, Scottish Government, the Northwest Regional Development Agency, British Heart Foundation and Cancer Research UK. The organization has over 150 dedicated members of staff, based in multiple locations across the UK.

**ABCD Acknowledgment:**

Data used in the preparation of this article were obtained from the Adolescent Brain Cognitive Development^SM^ Study (ABCD Study^®^) (https://abcdstudy.org), held in the NIMH Data Archive (NDA). This is a multisite, longitudinal study designed to recruit more than 10,000 children age 9–10 and follow them over 10 years into early adulthood. The ABCD Study is supported by the National Institutes of Health and additional federal partners under award numbers:

U01DA041022, U01DA041028, U01DA041048, U01DA041089, U01DA041106, U01DA041117, U01DA041120, U01DA041134, U01DA041148, U01DA041156, U01DA041174, U24DA041123, and U24DA041147

A full list of supporters is available at https://abcdstudy.org/federal-partners/. A listing of participating sites and a complete listing of the study investigators can be found at https://abcdstudy.org/principal-investigators.html. ABCD Study consortium investigators designed and implemented the study and/or provided data but did not necessarily participate in analysis or writing of this report. This manuscript reflects the views of the authors and may not reflect the opinions or views of the NIH or ABCD Study consortium investigators.

The ABCD data repository grows and changes over time. The ABCD data used in this came from [NIMH Data Archive Digital Object Identifier (10.15154/1523041)].

## URLS

PLINK (https://www.cog-genomics.org/plink/2.0/)

FUMA (https://fuma.ctglab.nl)

LDSC and sLDSC (https://github.com/bulik/ldsc)

Annotations for sLDSC tissue-specific analysis (https://alkesgroup.broadinstitute.org/LDSCORE/)

## List of Supplementary Materials

### Materials and Methods

Supplementary Table 1: Demographic breakdown of subsamples of UK Biobank

Supplementary Table 2: Demographic breakdown of subsamples of ABCD Study

Supplementary Table 3: Regions of interest and MRI atlases used

Supplementary Table 4: Regression analysis of PolyVoxel Score (PVS) with neurological disorders

Supplementary Table 5: Weighted quantile regression analysis of PVS with Parkinson’s Disease

Supplementary Table 6: Terms used to define categories for overlap of discoveries in Supplementary Figure 4

Supplementary Figure 1: Hyperparameter tunning curve of “Hemochromatosis Brain” classifier

Supplementary Figure 2: PolyVoxel Score (PVS) weight breakdown by each region of interest

Supplementary Figure 3: QQ plot of GWAS discoveries of PVS

Supplementary Figure 4: Overlap of GWAS discoveries with previous studies grouped by categories in Supplementary Table 6

Supplementary Figure 5: Genetic correlation of PolyVoxel Score with traits of interest

Supplementary Figure 6: Mendelian Randomization results

Supplementary Figure 7: Replication of GWAS results in non European ancestry sample

Supplementary Figure 8: Effect of age on PolyVoxel Score across 8-80 years old from ABCD and UKB samples

