## Supplementary Materials for "Hemochromatosis Neural Archetype Reveals Iron Disruption in Motor Circuits"

### **Materials and Methods:**

#### UK Biobank Sample

Genotypes, MRI scans, demographic and clinical data were obtained from the UK Biobank under accession number 27412, excluding 206 participants who withdrew their consent. All participants provided electronic signed informed consent and the study was approved by the UK Biobank Ethics and Governance Council. The recruitment period for participants was from 2006 to 2010, and participants had to be 40-69 years old during this period to be included in the sample. For the current study we analyzed data from a total sample of 38,937 individuals (20,435 females) with a mean age of 64.3 years (standard deviation 7.6 years) for analysis. This sample was made up of individuals who had qualified imaging (36,243 individuals) and/or who were C282Y homozygotes (2,888 individuals). We split the sample of individuals with qualified imaging into two non-overlapping groups: Subsample A for C282Y homozygote classifier training and Subsample B for deploying this classifier. A final group, subsample C, was used for weighted quantile regression (see below) which was Subsample B with the addition of all 2,888 C282Y homozygotes from the whole UKB sample. Supplementary Table 1 summarizes demographics of these three subsamples. Genotype, health record and neuroimaging data was collected from January 2006 to May 2021. Data analysis was conducted from January 2022 to October 2022. This study follows the Strengthening the Reporting of Observational Studies in Epidemiology ([STROBE](https://www.equator-network.org/reporting-guidelines/strobe/)) reporting guideline for cross-sectional studies^1^.

##### UK Biobank Image acquisition

T1 weighted and diffusion weighted scans were collected from three scanning sites throughout the United Kingdom, all on identically configured Siemens Skyra 3T scanners, with 32-channel receiver head coils. For diffusion scans, multiple scans with no diffusion gradient were collected (b=0 s/mm^2^) to fit diffusion models. The average of these b=0s/mm^2^ scans was used as voxel-wise measures of T2-weighted intensities. Diffusion-weighted scans were collected using a SE-EPI sequence at 2mm isotropic resolution. T1-weighted scans were collected using a 3D MPRAGE sequence at 1mm isotropic resolution. Voxelwise T2* values were estimated as part of the susceptibility-weighted imaging protocol at a voxel resolution of 0.8 × 0.8 × 3 mm and with 2 echoes (echo times, 9.42 and 20 milliseconds). To reduce noise, T2* images were spatially filtered (3 × 3 × 1 median filtering followed by limited dilation to fill missing data holes). If a person had multiple (longitudinal) scans we used the first scan. Further details of image acquisition can be found here^2^.

#### ABCD Sample^®^

The ABCD study is a longitudinal study across 21 data acquisition sites following 11,878 children starting at 9 and 10 years old. This paper analyzed the baseline and year 2 follow up sample from data release 4.0 (NDA DOI:10.15154/1523041). The ABCD study used school-based recruitment strategies to create a population-based, demographically diverse sample with heterogeneous ancestry. Genotype data was imputed using the TOPMED imputation server^3–5^ and genetic principal components were estimated using PC-AIR^6^ as described elsewhere^7^. We selected individuals who had passed neuroimaging and genetic quality control checks. We calculated participants’ continental genetic ancestry as calculated using SNPweights^8^ and aligning with 1k Genomes Project^9^, and indigenous reference panels^10^. Each individual was assigned an ancestry proportion to 4 continental groups: European (EUR), African (AFR), Native American (AMR), South Asian (SAS) or East Asian (EAS). From this participants were categorized into one of the three largest ancestry groups of European (EUR), African (AFR) and admixed (MIX). Individuals were categorized as EUR or AFR if they exceeded 80% ancestry within one of these continental ancestry groups and MIX if they were less than 80% ancestry across all groups. This resulted in 5,977 EUR, 687 AFR, and 3,135 MIX individuals for performing ancestry stratified analysis - see Supplementary Table 2 for demographic details.

##### ABCD Image acquisition

T1-weighted (T1w) and diffusion-weighted MRI (dMRI) scans were collected using Siemens Prisma and Prisma Fit, GE Discovery 750 and Phillips Achieva and Ingenia 3T scanners. Scanning protocols were harmonized across 21 acquisition sites. Full details of harmonization routines have been described elsewhere^11,12^. T1w images were acquired using a 3D MPRAGE scan at 1mm isotropic resolution. dMRI scans were acquired in the axial plane at 1.7mm isotropic resolution, with seven b=0s/mm^2^ frames. The average of b=0s/mm^2^ scans was taken as voxel-wise measures of T2-weighted intensities.

#### Image Preprocessing

Scans were corrected for nonlinear transformations provided by MRI scanner manufacturers^13,14^. T2-weighted and T2* scans (for UKB) were registered to T1-weighted images using mutual information^15^. Intensity inhomogeneity correction was performed by applying smoothly varying, estimated B1-bias fields^12^. Images were rigidly registered and resampled into alignment with a pre-existing, in-house, averaged, reference brain with 1.0 mm isotropic resolution^12^. Atlases used to define regions of interest for classifier feature importance analysis are shown in Supplementary Table 3.

#### C282Y Homozygote Classifier and PolyVoxel Score Generation

We aimed to train a classifier to predict C282Y homozygosity status from MRI scans. For training we generated a sample derived from the 193 C282Y homozygotes with qualified imaging and found covariate matched controls at a ratio of 4:1 (cases to controls). Controls were matched for sex, age, scanner and top ten principal components of genetic ancestry. This was performed as described elsewhere with the exception that controls were selected as those with no C282Y mutations (i.e. C282Y heterozygotes were excluded from being cases or controls). This resulted in a subsample of 193 C282Y “cases” (112 female) and 767 “controls” (463 female), i.e. no C282Y mutations, for training the classifier – we refer to this sample as Subsample A - see Supplementary Table 2 for description. For fitting, hyperparameter tuning and evaluating the classifier we employed a 5 fold cross-validation scheme in Subsample A.

The C282Y homozygote classifier was fitted using a PolyVoxel Score (PVS) framework^16^ as follows. Let
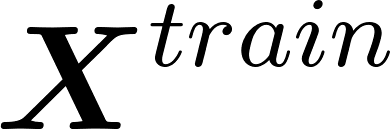
<https://www.codecogs.com/eqnedit.php?latex=%5Cboldsymbol%7BX%7D%5E%7Btrain%7D#0> represent the pre-residualized imaging matrix with *N* rows of individuals and *M* columns of voxels in the training sample. Each element of this matrix,
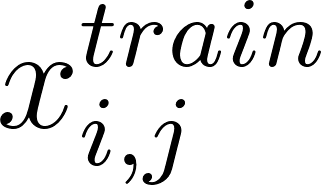
<https://www.codecogs.com/eqnedit.php?latex=x_%7Bi%2Cj%7D%5E%7Btrain%7D#0> represents the voxel intensity for the i-th individual at the j-th voxel for a T2-weighted scan. Here each column in
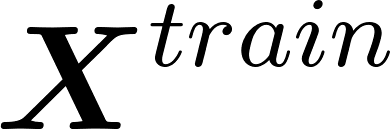
<https://www.codecogs.com/eqnedit.php?latex=%5Cboldsymbol%7BX%7D%5E%7Btrain%7D#0> was quantile transformed to a normal distribution and pre-residualized for covariates of age, sex, MRI scanner and top 10 principal components of genetics. Let
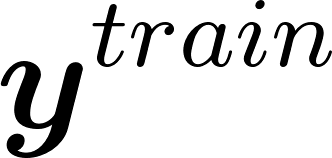
<https://www.codecogs.com/eqnedit.php?latex=%5Cboldsymbol%7By%7D%5E%7Btrain%7D#0> represent the 1-dimensional vector such that:

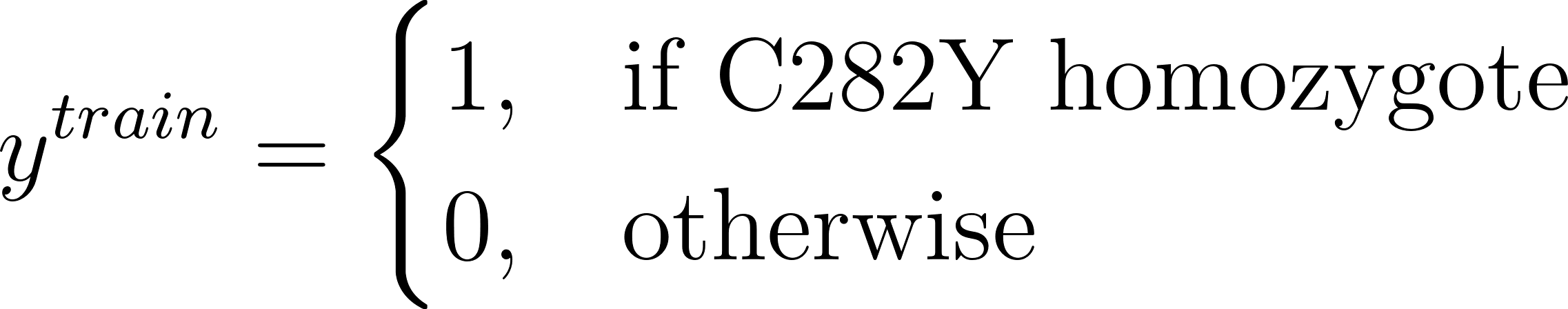
<https://www.codecogs.com/eqnedit.php?latex=y%5E%7Btrain%7D%3D%5Cbegin%7Bcases%7D%201%2C%20%26%20%5Ctext%7Bif%20C282Y%20homozygote%7D%20%5C%5C%5C%5C%200%2C%20%26%20%5Ctext%7Botherwise%7D%20%20%5Cend%7Bcases%7D#0>

We then regress each column of
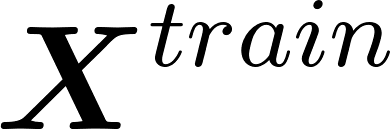
<https://www.codecogs.com/eqnedit.php?latex=%5Cboldsymbol%7BX%7D%5E%7Btrain%7D#0> with
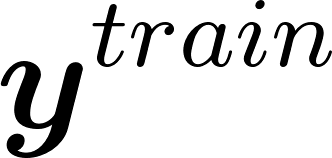
<https://www.codecogs.com/eqnedit.php?latex=%5Cboldsymbol%7By%7D%5E%7Btrain%7D#0> to generate a vector of univariate z-statistics,
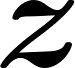
<https://www.codecogs.com/eqnedit.php?latex=%5Cboldsymbol%7Bz%7D#0>, of dimension *M* – where each element describes the univariate association of that voxel with C282Y homozygosity status. We then threshold this
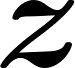
<https://www.codecogs.com/eqnedit.php?latex=%5Cboldsymbol%7Bz%7D#0> to restrict to nominally significant voxels in which p<0.01. Next, we could use the vector to generate out of sample PVS’s from a test sample’s imaging data, to estimate liability of individuals along the Hemochromatosis Brain liability scale as follows:
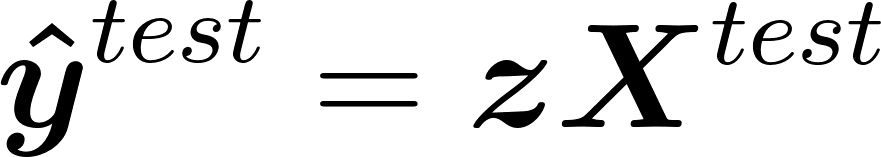
<https://www.codecogs.com/eqnedit.php?latex=%5Cboldsymbol%7B%5Chat%7By%7D%7D%5E%7Btest%7D%3D%5Cboldsymbol%7Bz%7D%5Cboldsymbol%7BX%7D%5E%7Btest%7D#0> (these univariate statistics are displayed in Figure 1). However, due to the correlation structure across voxels this gives suboptimal out of sample prediction. As such we use the correlation matrix of
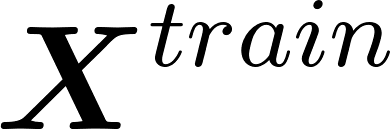
<https://www.codecogs.com/eqnedit.php?latex=%5Cboldsymbol%7BX%7D%5E%7Btrain%7D#0>,
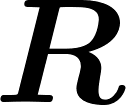
<https://www.codecogs.com/eqnedit.php?latex=%5Cboldsymbol%7BR%7D#0>, to re-weight
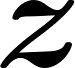
<https://www.codecogs.com/eqnedit.php?latex=%5Cboldsymbol%7Bz%7D#0> to obtain posterior effect sizes.
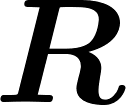
<https://www.codecogs.com/eqnedit.php?latex=%5Cboldsymbol%7BR%7D#0> can be decomposed using singular value decomposition
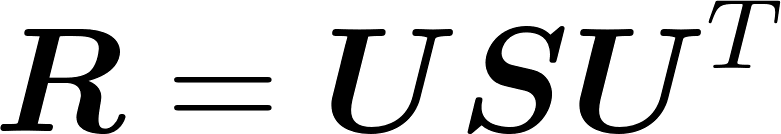
<https://www.codecogs.com/eqnedit.php?latex=%5Cboldsymbol%7BR%7D%20%3D%20%5Cboldsymbol%7BU%7D%5Cboldsymbol%7BS%7D%20%5Cboldsymbol%7BU%7D%5ET#0> (
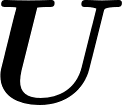
<https://www.codecogs.com/eqnedit.php?latex=%5Cboldsymbol%7BU%7D#0> – unitary matrix,
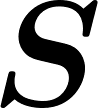
<https://www.codecogs.com/eqnedit.php?latex=%5Cboldsymbol%7BS%7D#0> – diagonal matrix with singular values on its diagonal). We consider the regularized form of
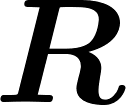
<https://www.codecogs.com/eqnedit.php?latex=%5Cboldsymbol%7BR%7D#0> as:
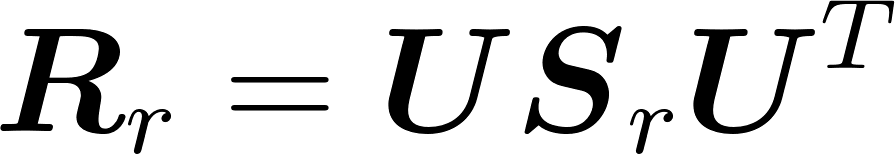
<https://www.codecogs.com/eqnedit.php?latex=%5Cboldsymbol%7BR%7D_r%20%3D%20%5Cboldsymbol%7BU%7D%5Cboldsymbol%7BS%7D_r%20%5Cboldsymbol%7BU%7D%5ET#0> , where
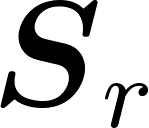
<https://www.codecogs.com/eqnedit.php?latex=%5Cboldsymbol%7BS%7D_r#0> is obtained from
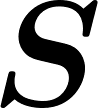
<https://www.codecogs.com/eqnedit.php?latex=%5Cboldsymbol%7BS%7D#0> by keeping
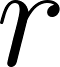
<https://www.codecogs.com/eqnedit.php?latex=r#0> largest singular values and replacing the remaining with
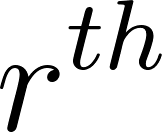
<https://www.codecogs.com/eqnedit.php?latex=r%5E%7Bth%7D#0> largest. We then decorrelate association statistics to generate PVS classifier weights (posterior effects in Figure 1) as
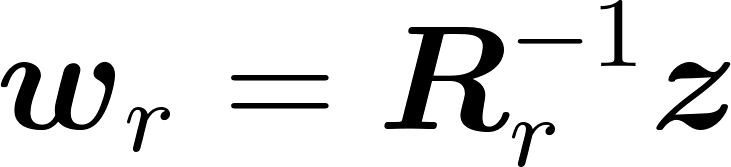
<https://www.codecogs.com/eqnedit.php?latex=%5Cboldsymbol%7Bw%7D_r%3D%5Cboldsymbol%7BR%7D_r%5E%7B-1%7D%5Cboldsymbol%7Bz%7D#0> which were in turn are used to generate PVS scores for each individual as:
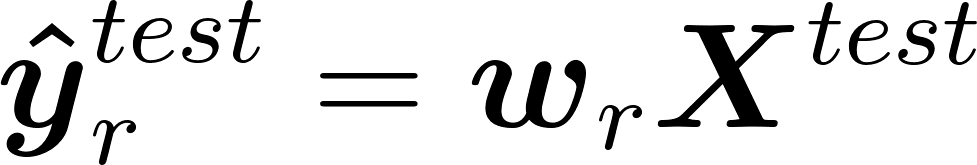
<https://www.codecogs.com/eqnedit.php?latex=%5Cboldsymbol%7B%5Chat%7By%7D%7D%5E%7Btest%7D_r%3D%5Cboldsymbol%7Bw%7D_r%5Cboldsymbol%7BX%7D%5E%7Btest%7D#0>. During cross-validation in Subsample A, nine
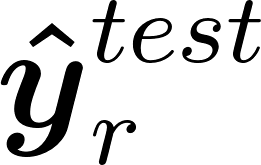
<https://www.codecogs.com/eqnedit.php?latex=%5Cboldsymbol%7B%5Chat%7By%7D%7D%5E%7Btest%7D_r#0> were generated with different values of (1, 5, 10, 20, 50, 100, 200, 500, 1000 and M (i.e. no regularization)). The
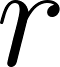
<https://www.codecogs.com/eqnedit.php?latex=r#0> value that maximized the association between and
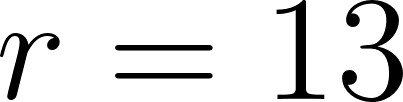
<https://www.codecogs.com/eqnedit.php?latex=r%3D13#0> was selected for displaying the receiver operator characteristic (ROC) curve in Figure 1. For downstream analysis in subsample B we used the mean optimal value,
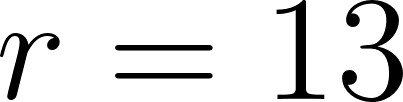
<https://www.codecogs.com/eqnedit.php?latex=r%3D13#0>, to fit posterior weights across the whole of subsample A.

To capture feature importance of the classifier and univariate weights for different brain regions (Figure 1 and Supplementary Figure 2), we normalized
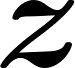
<https://www.codecogs.com/eqnedit.php?latex=%5Cboldsymbol%7Bz%7D#0> and
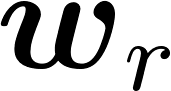
<https://www.codecogs.com/eqnedit.php?latex=%5Cboldsymbol%7Bw%7D_r#0> to be of unit length as
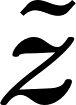
<https://www.codecogs.com/eqnedit.php?latex=%5Cboldsymbol%7B%5Ctilde%7Bz%7D%7D#0> and
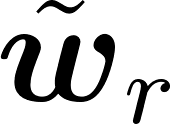
<https://www.codecogs.com/eqnedit.php?latex=%5Ctilde%7B%5Cboldsymbol%7Bw%7D%7D_r#0>. We then calculated the sum of squares of
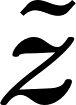
<https://www.codecogs.com/eqnedit.php?latex=%5Cboldsymbol%7B%5Ctilde%7Bz%7D%7D#0> and
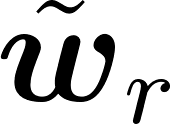
<https://www.codecogs.com/eqnedit.php?latex=%5Ctilde%7B%5Cboldsymbol%7Bw%7D%7D_r#0> for each voxel that fell within a given brain region (see Supplementary Table 3 for atlases used to define brain regions). For plotting results of weighted quantile regression (Figure 3B) we estimated mean iron concentration was calculated for each individual by taking the mean across p<0.01 voxels from T2* imaging^17^. We then used previously published estimates linking R2* (1/T2*) values to iron concentration as [μg/g dry] = C × R2*[Hz] / 3.2, where C = 2000 / 36.

#### Genome Wide Association of Hemochromatosis Brain

In Subsample B we then performed a GWAS with the Hemochromatosis Brain PVS as the phenotype of interest, covarying for age, sex and top ten components of genetic ancestry. We restricted this analysis to individuals declared as self-identified “White British” and similar genetic ancestry (using Data Field: 22006), this resulted in 30,709 individuals. 4,608 remaining individuals from Subsample B were used for replication of GWAS discoveries. Additionally, to enable the calculation of genetic correlations of related traits in UK Biobank, we performed GWAS for intracranial volume, as a measure of brain size, and 4 red blood cell traits: mean corpuscular volume, mean corpuscular hemoglobin, mean sphere cell volume and hemoglobin concentration. If there were multiple instances of a variable we took their mean value for each subject. For these measures, as well as the PVS, we quantile transformed them to a normal distribution to enforce normality and reduce leverage of outliers. We used sample and variant QC’s of: --geno 0.05, --hwe 1e-12, --maf 0.005 and --mind 0.1 using PLINK (v2.00a3.6LM)^18^. This resulted in 7,131,446 remaining variants and no individuals removed. PLINK was used for performing GWAS across these 6 traits (using –glm). For discovered variants we performed validation in 4,608 individuals who self-identified in any other category as “White British” by performing the same process for GWAS discovery. Using independent significant SNPs (see locus definition below) from the discovery set we evaluated replication in two ways: 1) compared/correlated coefficients and 2) evaluated sign concordance of coefficients between discovery and replication cohorts. For 2) we used a binomial test to assess the significance of in proportion of sign concordant SNPs.

#### Gene Burden Analysis Using Whole Exome Data

To test if rare genetic variants were associated with variability in brain iron levels captured by the Hemochromatosis Brain PVS we performed gene burden analysis using Reginie (v3.1.1)^19^. For this analysis we had 19,498 individuals from Subsample B with available genetic data. Using annotations from SnpEff^20^ (v5.0) of each observed mutation we applied a mask of mutations labeled as high or moderate consequence. Using parameters of --aaf-bins 0.1,0.05,0.01 and --bsize 200, and the same covariates described above we associated each binary gene burden score with variability in PVS. Across 18,860 genes tested we defined discoveries as those whose p-value exceeded Bonferroni significance of 0.05/18,860.

#### GWAS Locus Definition and Discovery Overlap GWAS Catalog

Summary statistics were uploaded to FUMA (FUMA: v1.4.1, MAMGA v1.08)^21^ for locus definition and gene mapping. “Independent significant SNPs” represent genome-wide significant SNPs that are in moderate to low LD with one another (r_LD_<0.6). “Genome-wide loci” represent regions in which independent significant SNPs are merged if they are: a) in moderate LD with one another (r_LD_>0.1) or b) are physically close to one another (<250kb). FUMA provides an output of independent significant SNPs and reports which of these overlap with previous studies reported in GWAS catalog (e104_r2021-09-15). We defined novel loci as those which did not contain any independent significant SNPs overlapping with associations in GWAS catalog. For SNPs that did overlap with previous GWAS catalog studies, we grouped these previously reported traits into broad categories defined in Supplementary Table 4. We allowed each significant SNP to be counted for multiple categories if it appeared in different studies across categories. However, we ensured that it was not counted more than once within a specific category, even if it appeared in multiple studies within that category. The results of this analysis are shown in Supplementary Figure 4 and extended data tables.

#### Mendelian Randomization

To assess the causal strength of association between peripheral blood markers, PD and brain iron dysregulation as measured by the Hemochromatosis Brain we conducted mendelian randomization using GSMR^22^. In order to conduct these analyses we used previous GWAS summary statistics of serum iron, transferrin saturation, total iron binding capacity, ferritin^23^, and parkinson's disease^24^. Linkage disequilibrium (LD) was estimated from the 30,709 UK Biobank individuals used for the PVS GWAS; this LD was then used to find independent significant SNPs to use as instruments for GSMR. The following parameters were used: gwas_thresh=5x10^-8^, single_snp_hedi_thresh=0.01, multi_snps_hedi_thresh=0.01, heidi_outlier_flag=T, ld_r2_thresh=0.05, ld_fdr_thresh=0.05. Bivariate GSMR was run (in both directions) to confirm the directionality of causation.

#### Genetic Correlation

LDSC^25^ was used to estimate from summary statistics using the 1k genomes European ancestry reference panel. LDSC was also used to calculate genetic correlations with GWAS of intracranial volume and four red blood cell traits (mean corpuscular hemoglobin, mean corpuscular volume, mean sphered cell volume and hemoglobin concentration) that we conducted in Subsample B, as well as publicly available summary statistics of four blood iron traits^23^, Parkinson’s disease^24^ and educational attainment^26^.

#### Single Cell Enrichment

We performed cell type enrichment using the FUMA platform^21^. Association statistics at the gene level were calculated using MAGMA (v1.08)^27^. This gene-based distribution was then associated with expression patterns of 25 cell types from PsychENCODE single-cell RNA-seq data collected from the adult brain^28^ by performing a gene-property analysis^29^.

#### Tissue-Specific Enrichment

We used stratified LDSC (S-LDSC)^30^ to examine tissue-type specific enrichment of the Hemochromatosis Brain GWAS results. We analyzed annotations derived from 489 tissue-specific chromatin assays of bulk tissue from peaks for six epigenetic marks. These data were collected as part of the Roadmap Epigenomics and ENCODE projects^31,32^, annotations were downloaded from previous analysis^33^ (see URLs). We controlled for baseline annotations as recommended by S-LDSC^30^. We reported the signed enrichment Z statistics, as well as the corresponding FDR corrected multiple comparisons adjusted p-values.

#### Polygenic/PolyVoxel Score Validation

We tested the generalization of both our genetic and neuroimaging findings of the Hemochromatosis Brain to individuals from the ABCD cohort: a cohort on average more than 50 years younger than the UKB cohort from which these findings were discovered. Within the ABCD sample we generate a PVS score for each individual at each available time point. This included 9,799 individuals at baseline (age 8.9-11.1 years) and 4,615 at 2 year follow up (age 10.58-13.67years). To generate polygenic scores we calculated posterior effect sizes using PRScs^34^ using the LD estimated from the 1k Genome European ancestry reference panel (N=503). Polygenic scores were then calculated by applying these posterior effect sizes using ‘plink –score’ (v2.00) to ABCD genetic data. Performing ancestry stratified analysis (within the groups described above) we fitted linear mixed effect models to predict the PVS of each individual with fixed effects of sex, age, top 10 PCs of genetic ancestry, and a categorical variable indicating the mri software and serial number. We included random intercepts for subject id (to account for longitudinal measures) nested with family id membership. Variance explained was computed as
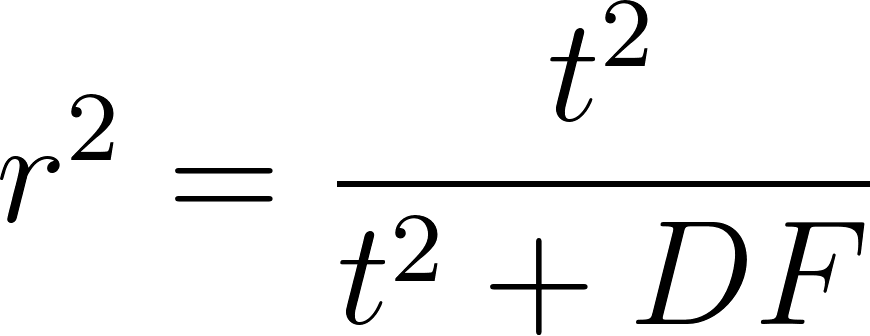
<https://www.codecogs.com/eqnedit.php?latex=r%5E2%20%3D%20%5Cfrac%7Bt%5E2%7D%7Bt%5E2%20%2B%20DF%7D#0>, where DF is the degrees of freedom.

#### Disease Enrichment of Hemochromatosis Brain

We wanted to test the utility and specificity of the Hemochromatosis Brain PVS in distinguishing cases and controls of PD and other disorders of the brain. To do this we defined Subsample B in UK Biobank who were the remainder of individuals with neuroimaging that were not in Subsample A. This resulted in 35,283 individuals (18,278 females, 0 C282Y homozygotes). For this sample we generated a PVS for each individual from their imaging data using the C282Y homozygote classifier, as described above, trained in the full Subsample A (regularization parameter set to r=13 as the mean optimal value across cross-validation). This PVS is a single score per individual that in essence quantifies the degree of iron dysregulation in motor circuits observed in C282Y homozygotes. We then associated this Hemochromatosis Brain PVS with 6 disorders/classes of disorders of the brain using weighted regression logistic models with covariates of age, sex, scanning site and top ten components of genetic ancestry. These 6 different disorders/categories were: Other disorders of the Nervous System (ICD10: G90-99), Movement Disorders (ICD10: G20-G26), Parkinson’s Disease (ICD10: G20), Essential Tremor (ICD10:G25), Other degenerative diseases of the Nervous System (including Alzheimer’s) (ICD10: G30-32) and Abnormalities of Gait and Mobility (ICD10: R26). These diagnoses were extracted from UK Biobank field 41270. We performed weighted regression using inverse probability weighting (IPW)^35^ to account for the depleted number of cases of each of these six disorders in the imaging subsample vs the whole UK Biobank sample. I.e. If the neur-imaging sample had 3 fold lower prevalence for a disorder when compared with the whole sample, we weighted cases with an IPW of 3 and controls with a weight of 1. This enabled regression across individuals in the imaging and non-imaging samples (see section below) and estimates closer to population prevalence for these disorders. IPW values for each disorder are shown in supplementary table 4.

#### Quantile Weighted Regression

Individuals were assigned to one of 4 quantiles 1-4 according to their PVS with 1^st^ being the lowest 25% of PVS, 2^nd^ being scores between 25^th^ and 50^th^ percentile and so on. We aimed to quantify the PD risk of each of these quantiles with reference to C282Y homozygotes taken from the whole UKB sample, including those that did not have neuroimaging (2,888 homozygotes taken from 488,288 individuals). In order to accomplish this we again performed weighted regression using an IPW of 3.89 for PD cases in the imaging sample and 1 for all other observations. A model was then (within sample C of Supplementary Table 1) fit to predict PD from these quantiles and C282Y homozygote status as categorical variables covarying for the same covariates described above. We then used estimated mean iron concentrations for PVS brain regions, as described above, to display the mean of this value for each group as shown on the x axis of Figure 3B.

**Data Availability:** UK Biobank data was accessed under accession number 27412. Summary statistics from GWAS of Hemochromatosis Brain PVS are available for [download](https://drive.google.com/drive/folders/1PA7Amizkq08tR5EI0FnG0VoEXOGOmVF3?usp=sharing). Researchers can apply for access to UK Biobank data at <https://www.ukbiobank.ac.uk/enable-your-research/apply-for-access>. The ABCD data used in this came from [NIMH Data Archive Digital Object Identifier (10.15154/1523041)]. This data is available to approved researchers, find more information at https://abcdstudy.org.

**Code Availability:** We have created a python command line program to generate PVS’s for T2-Weighted images registered in Montreal Neurological Institute space using classification weights presented in this analysis - <https://github.com/robloughnan/pvs>.

**Supplementary Note:**

#### Potential Spurious Locus

We identified in total 43 genomic loci associated with the Hemochromatosis Brain PVS. However one of these loci displayed a suspicious association pattern with nearby SNPs in LD - see Supplementary Figure 4.24 - and such was removed from the final loci count.

### 1. Supplementary Tables

|  | **UK Biobank Subsample** | | | **UK Biobank Full Sample** |
| --- | --- | --- | --- | --- |
|  | **A** | **B** | **C** |  |
| ***n*** | 960 | 35,283 | 38,170 | 502,413 |
| ***Age, mean (SD)*** | 64.3 (7.5) | 64.1 (7.5) | 64.3 (7.6) | 66.2 (8.13) |
| ***Female, n (%)*** | 575 (60.0) | 18,378 (52.1) | 19,972 (52.3) | 273,328 (54.4) |
| ***C282Y Homozygote (%)*** | 193 (20.0) | 0 (0.0) | 2,888 (7.56*) | 2,888 (0.57) |
| ***C282Y Heterozygote (%)*** | 0 (0.0) | 4,935 (14.0) | 4,935 (14.0) | 65,299 (13.0) |
| ***Parkinson’s Disease Cases (%)*** | 0 (0.0) | 64 (0.18) | 93 (0.24) | 3,447 (0.68) |

*Supplementary Table 1 Demographic breakdown of each subsample of UK Biobank. Subsample A was used for PVS training (and cross validation). Subsample B was used for generation of PVS scores and GWAS discovery. Subsample C was used for weighted quantile regression (main figure 3b) - it only differs from subsample B with the inclusion of 2,888 C282Y homozygotes from the entire sample. * Note this percentage represents the homozygote rate for subsample C, the homozygote rate across the whole UKB sample is 0.57%. The final column of ‘full sample’ is there for reference of PD cases but is not used directly for any analysis.*

|  | **ABCD Subsample** | | |
| --- | --- | --- | --- |
|  | **EUR** | **AFR** | **MIX** |
| ***n*** | 5,977 | 687 | 3,135 |
| ***Baseline Age, mean (SD)*** | 9.93 (0.63) | 9.91 (0.60) | 9.91 (0.60) |
| ***Year 2 Age, mean (SD)*** | 11.94 (0.64) | 11.94 (0.63) | 11.90 (0.65) |
| ***Female, n (%)*** | 2,856 47.78) | 351 (51.09) | 1,535 (48.96) |
| ***C282Y Homozygote (%)*** | 16 (0.27) | 0 (0.0) | 0 (0.00) |
| ***C282Y Heterozygote (%)*** | 684 (11.44) | 6 (0.87) | 133 (4.24) |

*Supplementary Table 2: Demographic breakdown of each ancestry strata for ABCD sample.*

| ***FreeSurfer 5.3 segmentation***  ***T1*** | ***Pauli, 2018***  ***HCP T1 & T2*** |
| --- | --- |
| *Cerebellum White Matter*  *Cerebellum Gray Matter*  *Pallidum*  *Thalamus* | *Putamen*  *Caudate*  *Substantia nigra pars compacta*  *Substantia nigra pars reticulata*  *Red nucleus*  *Subthalamic nucleus* |

*Supplementary Table 3* ***Regions of interest (ROIs) labeled using 2 different methods.*** *Column* *1) automatic segmentation using FreeSurfer 5.3 applied to each subject’s T1 image in atlas space**^36,37^**; Column 2) registration of the Pauli atlas of subcortical nuclei to the multispectral atlas**^37^*

| **Diagnosis** | **Controls** | **Cases** | **IPW for Cases** | **OR** | **CI_lower** | **CI_upper** | **Z** | **P** |
| --- | --- | --- | --- | --- | --- | --- | --- | --- |
| **Abnormalities of Gait and Mobility [R26]** | 35077 | 206 | 3.63 | 0.8881 | 0.8274 | 0.9533 | -3.2846 | 0.001 |
| **Other degenerative diseases of Nervous System**  **(including Alzheimer’s) [G30-32]** | 35223 | 60 | 5.70 | 1.0823 | 0.9754 | 1.201 | 1.4901 | 0.1362 |
| **Parkinson's Disease [G20]** | 35219 | 64 | 3.89 | 0.7356 | 0.6519 | 0.8302 | -4.978 | 6.42E-07 |
| **Essential Tremor [G25]** | 35201 | 82 | 1.95 | 1.0334 | 0.8865 | 1.2048 | 0.4201 | 0.6744 |
| **Movement Disorders [G20-26]** | 35116 | 167 | 2.65 | 0.9253 | 0.8437 | 1.0148 | -1.6487 | 0.0992 |

*Supplementary Table 4 Association results from regression in Subsample B of PolyVoxel Score (PVS) with each neurology disorder or collection of disorders.*

|  | *OR* | *z* | *P>\|z\|* |
| --- | --- | --- | --- |
| *1st Quantile* | *3.276* | *5.724* | *1.04x10^-8^* |
| *2nd Quantile* | *2.606* | *4.417* | *1.00x10^-5^* |
| *3rd Quantile* | *1.801* | *2.567* | *1.03x10^-2^* |
| *C282Y Homozygosity* | *2.470* | *3.445* | *5.72x10^-4^* |

*Supplementary Table 5 Weighted Quantile Regression to predict PD status. Results from regression in subsample C with single categorical variable indicating PVS quantile or C282Y homozygosity status. The reference group was the 4th PVS quartile group. PD cases in the imaging cohort were upweighted with a frequency of 3.89 to correct for PD case depletion within neuroimaging (see Supplementary Table 1), all other observations were given a weighting of 1.*

| ***CATEGORY*** | ***SEARCH TERMS*** | |
| --- | --- | --- |
| ***IRON/RED BLOOD CELLS*** | *iron*  *hematocrit*  *ferritin*  *red cell*  *red blood*  *bilirubin* | *hemoglobin*  *hemotcrit*  *hepcidin*  *corpuscular*  *reticulo*  *hereditary hemochromatosis* |
| ***BRAIN/COGNITION*** | *brain*  *white matter*  *intelligence*  *lentiform nucleus*  *nucleus accumbens*  *highest math class taken* | *cognitive*  *cortical*  *educational attainment*  *math ability*  *grey matter* |
| ***BLOOD MARKERS (NON-IRON)*** | *lipoprotein*  *metabolite*  *cholesterol*  *triglyceride*  *alkaline phosphatase levels*  *eosinophil*  *serum 25-hydroxyvitamin d levels*  *serum albumin level*  *lipid traits*  *monocyte*  *liver enzyme levels* | *protein*  *platelet*  *white blood cell*  *aminotransferase levels*  *globulin levels*  *testosterone levels*  *basophil*  *calcium levels*  *insulin-like growth*  *adiponectin levels*  *thyroxine concentration* |
| ***URATE*** | *urate*  *uric acid* |  |
| ***BMI/WAIST-HIP RATIO*** | *body mass index*  *waist-hip*  *body shape index*  *hip circumference* | *bmi*  *body fat*  *waist circumference*  *hip index* |
| ***BLOOD PRESSURE*** | *blood pressure*  *hypertension*  *arterial pressure*  *pulse pressure* |  |
| ***PSYCHOPATHOLOGY*** | *neuroticism*  *schizophrenia*  *depressive symptoms*  *bipolar disorder*  *anorexia nervosa* |  |
| ***HEIGHT*** | *height* |  |
| ***LEUKEMIA*** | *leukemia* |  |
| ***ALCOHOL CONSUMPTION*** | *alcohol consumption*  *alcohol use* |  |
| ***CHRONOTYPE*** | *morning person*  *daytime sleep*  *morningness*  *sleep*  *getting up in the morning* | |
| ***DIABETES*** | *diabetes* |  |
| ***CARDIOVASCULAR*** | *cardiovascular* |  |
| ***BALDNESS*** | *baldness*  *balding* |  |
| ***GRIP STRENGTH*** | *grip strength* |  |
| ***BONE/OSTEOARTHRITIS*** | *bone*  *osteoarthritis* |  |
| ***PERSONALITY/AFFECT*** | *adventurousness*  *positive affect*  *worry*  *empathy quotient* | *risk tolerance*  *life satisfaction*  *leisure sedentary behaviour* |
| ***EYE/RETINAL*** | *optic cup area*  *iris*  *vertical cup-disc ratio*  *eye*  *refractive error* |  |
| ***MEDICATION USE*** | *medication use* |  |
| ***PARKINSONS*** | *parkinson* |  |
| ***GOUT*** | *gout* |  |
| ***BIRTH WEIGHT*** | *birth weight* |  |
| ***NUTRITION*** | *consumption*  *meat-related diet* |  |
| ***ASTHMA*** | *asthma* |  |

*Supplementary Table 6: Definition of search terms to define each category when comparing GWAS discoveries with overlapping traits from GWAS catalog - used to generate Supplementary Figure 4.*

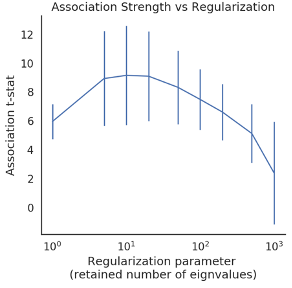

*Supplementary Figure 1 Association strength (t-stat) between “Hemochromatosis Brain” Classifier and C282Y homozygote status (y-axis) and regularization parameter (x-axis) – number of retained eigenvalues for SVD truncation (see methods). Points indicate association strength in test folds of 5-fold cross validation in Subsample B. r=13 was used for PVS generation in subsample B.*

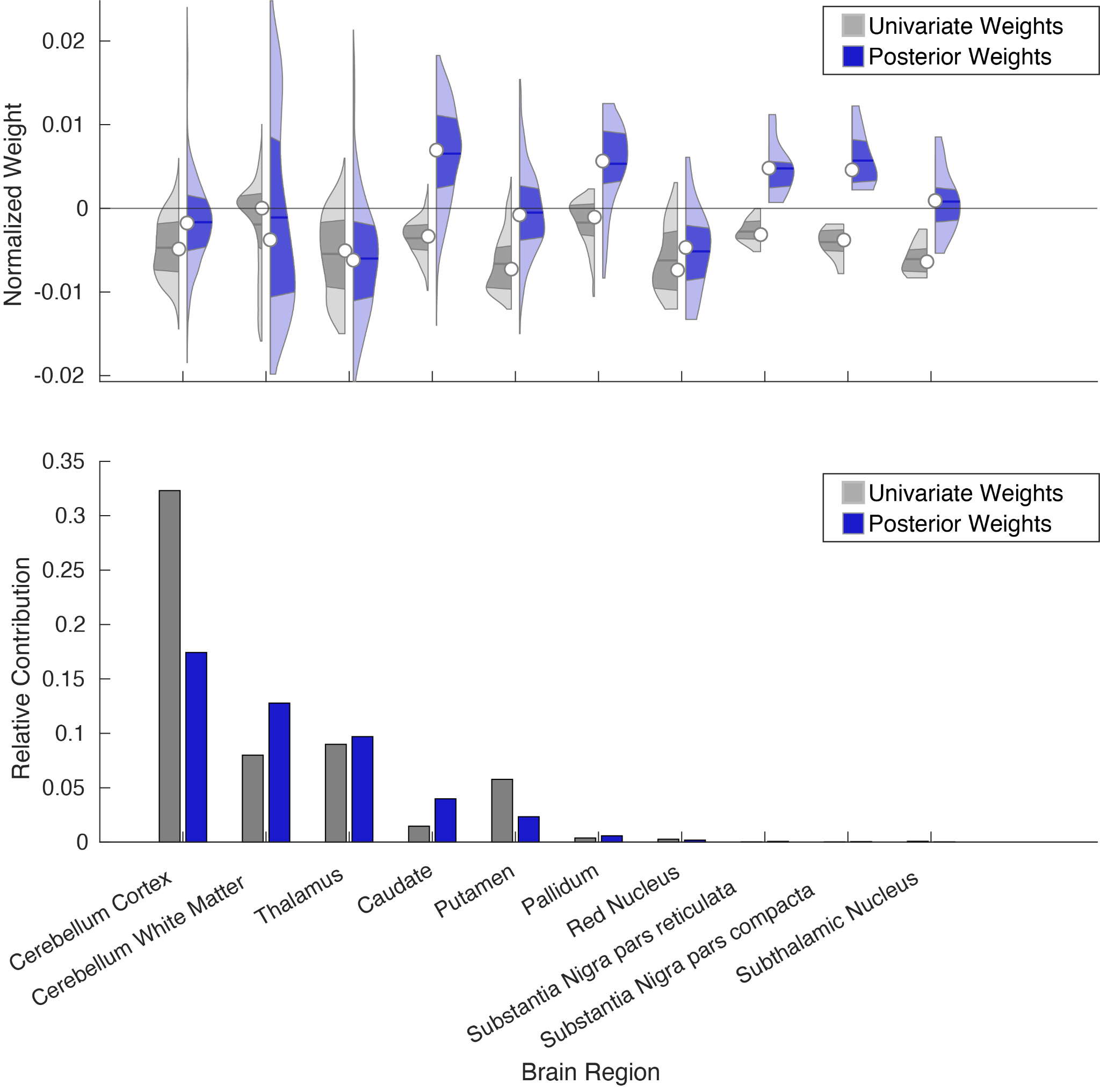

*Supplementary Figure 2: PolyVoxel Score (PVS) weight breakdown by each region of interest (ROI). Top panel indicates distribution non zero PVS weights for each region, where univariate statistics and posterior weight vectors are normalized to be of unit length. Bottom panel indicates the relative contribution to each region as the sum of squares within each brain region for univariate statistics and posterior weights. Note: univariate weights are shown for reference however posterior weights were used for PVS generation.*

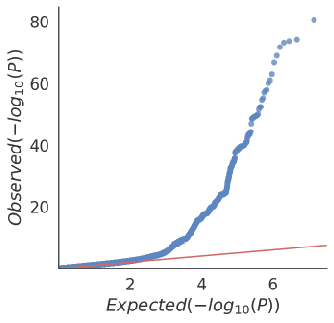

*Supplementary Figure 3: QQ plot of GWAS of PVS in European ancestry individuals within UKB subsample B (30,709 individuals).*

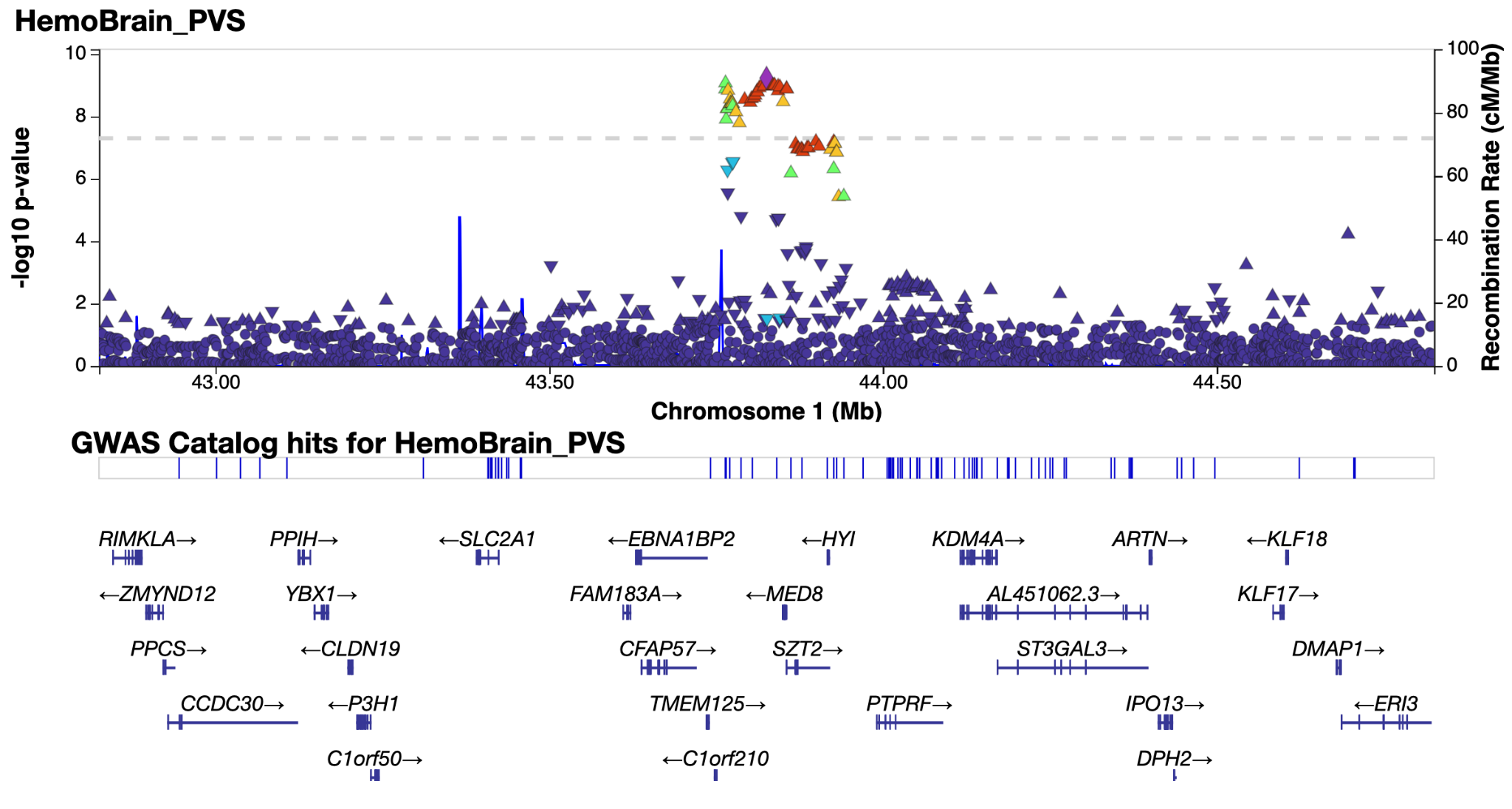

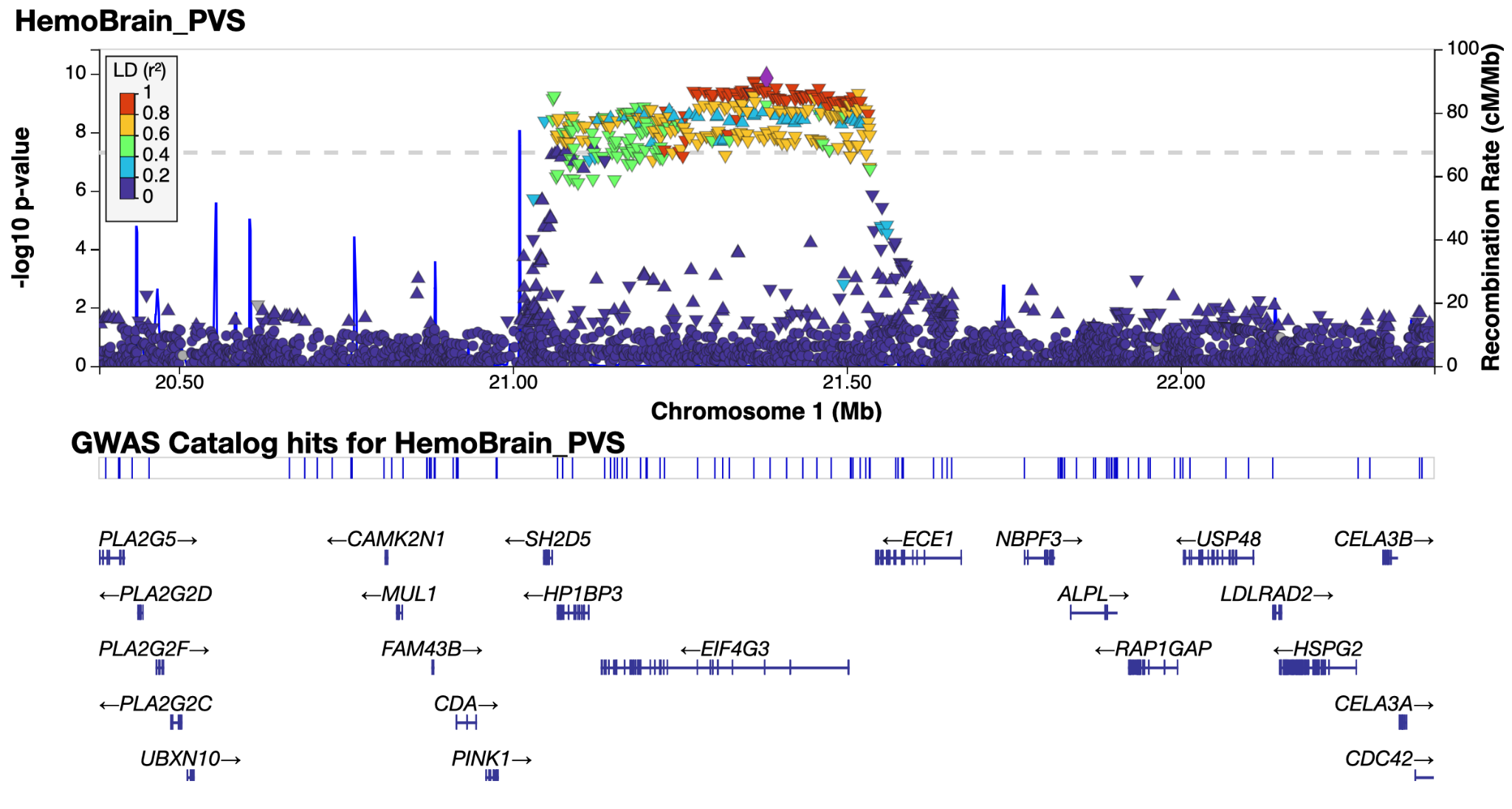

*Supplementary Figure 4.1 & 4.2: LocusZoom**^38^* *plot of genomic loci 1 and 2 - see extended data tables for genomic locus numbering.*

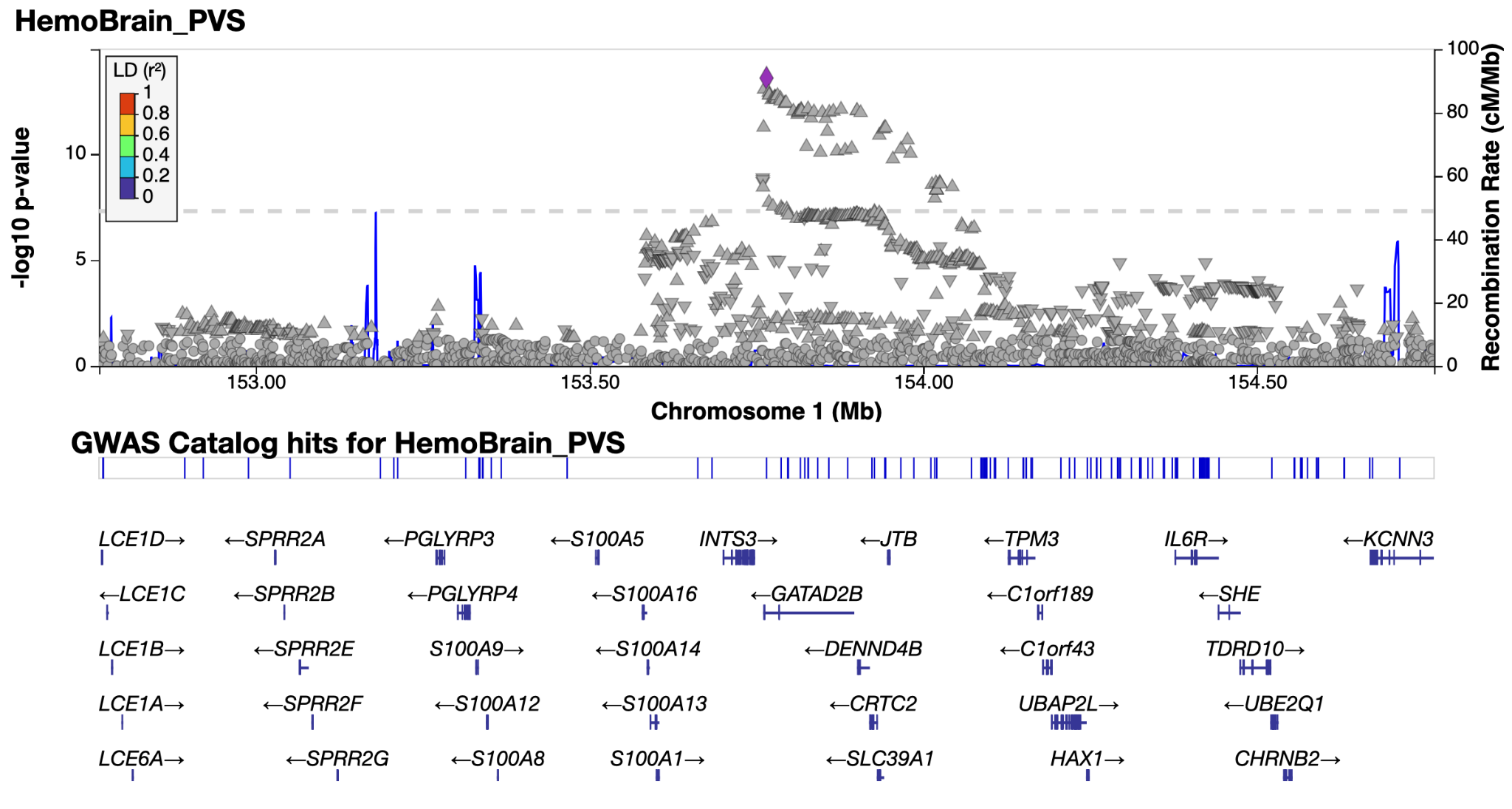

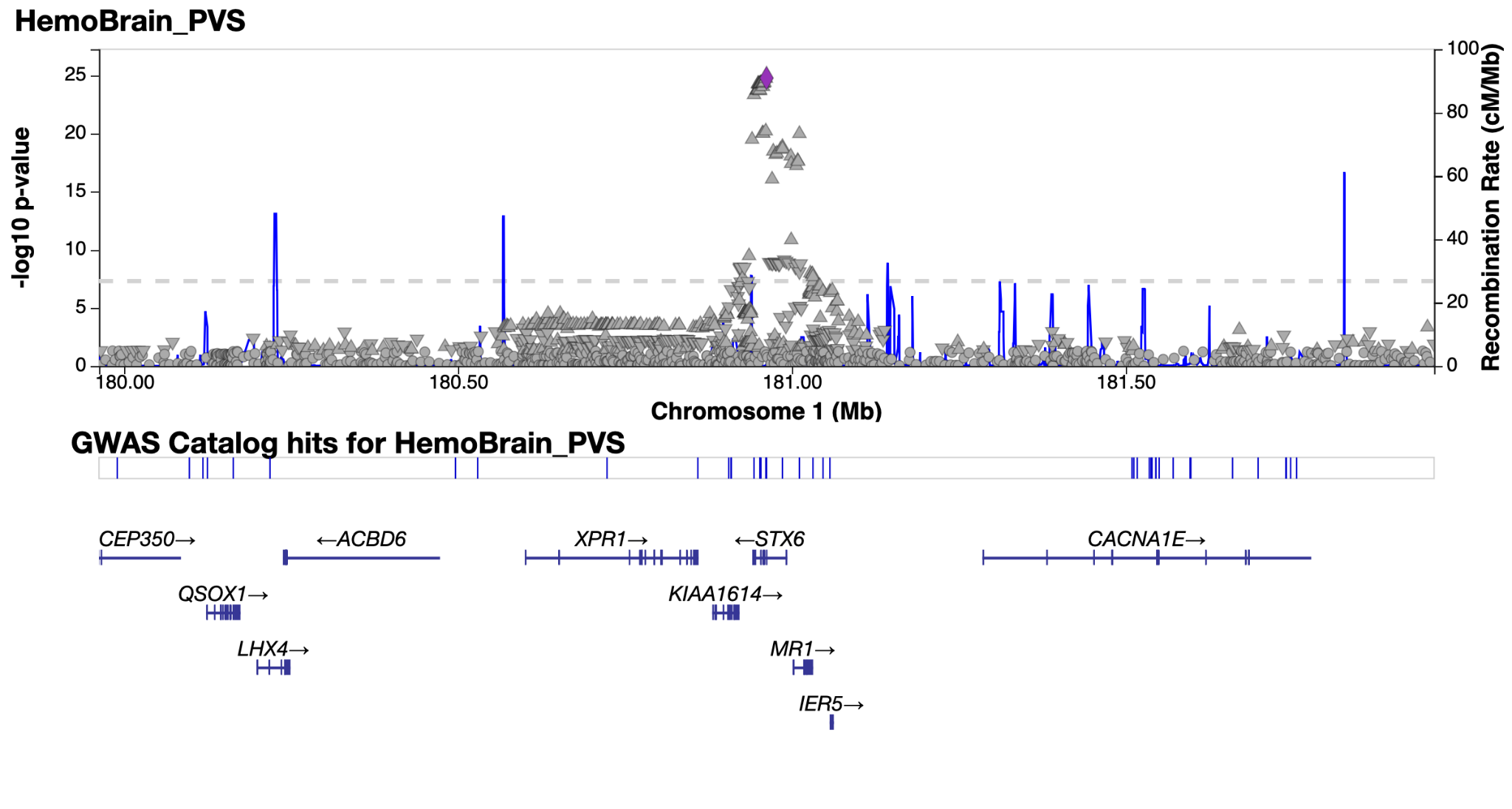

*Supplementary Figure 4.3 & 4.4: LocusZoom**^38^* *plot of genomic loci 3 and 4 - see extended data tables for genomic locus numbering. Non-lead SNP points are in gray as lead SNP is not in LD reference panel (1k genomes).*

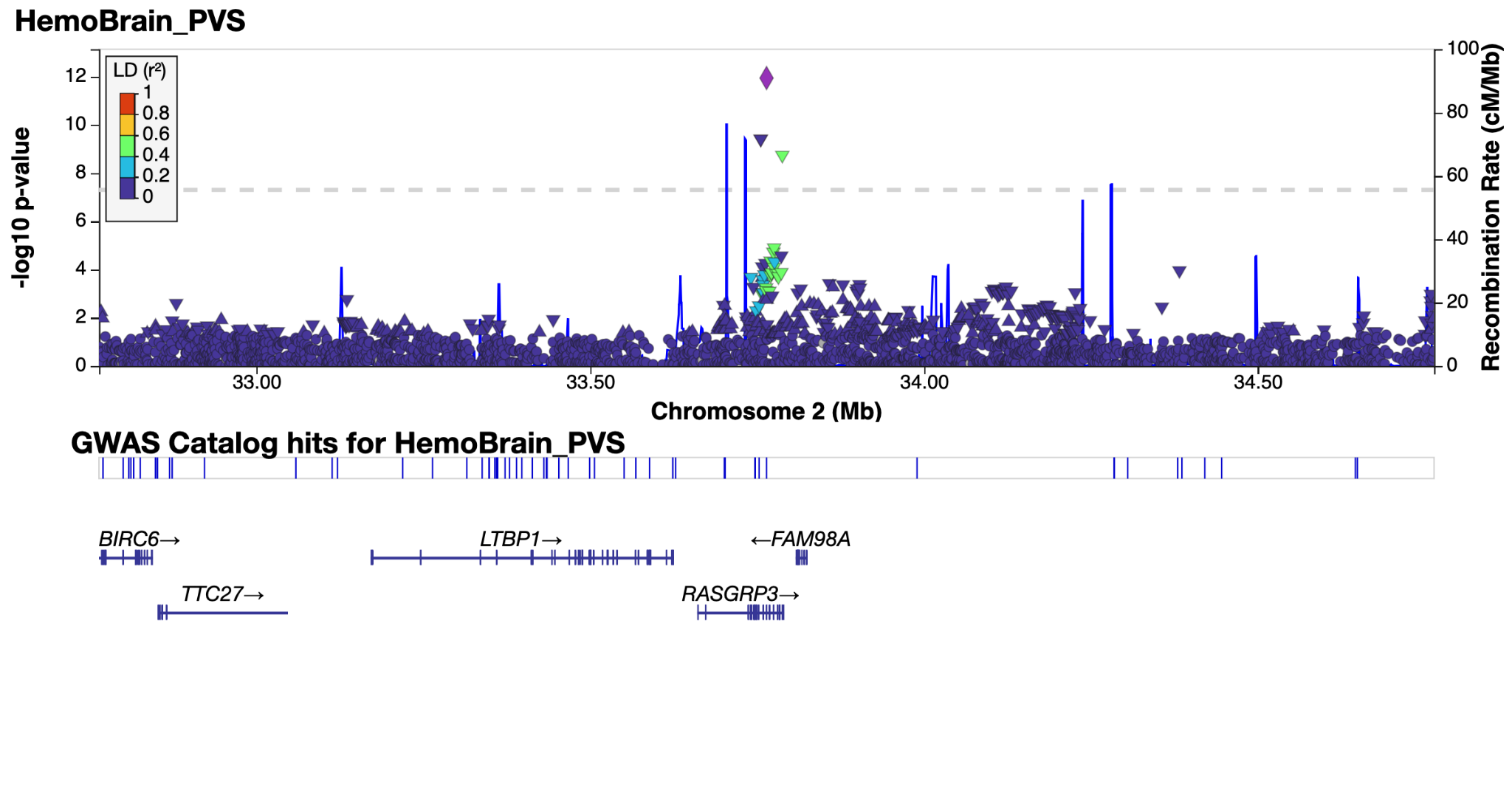

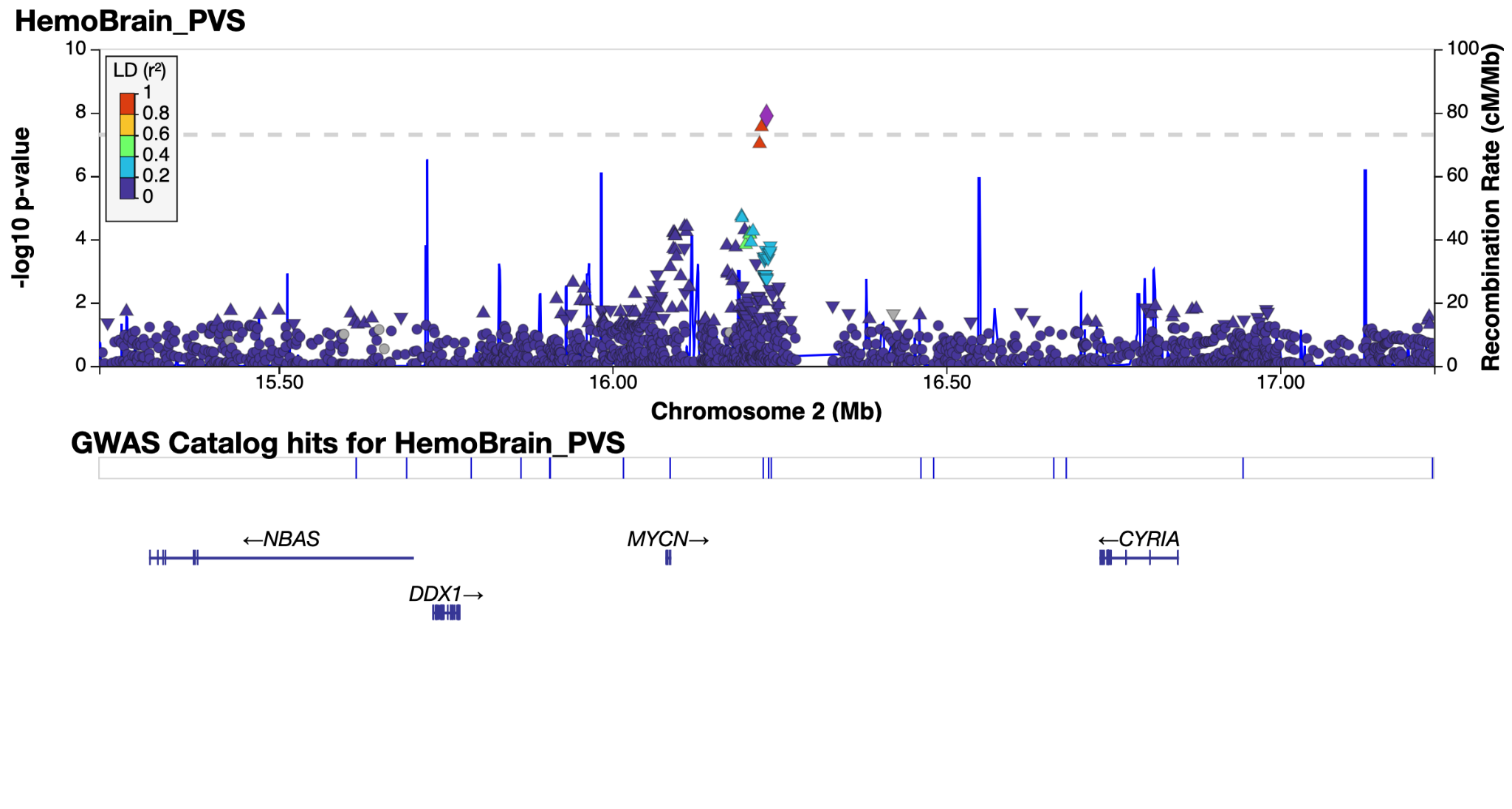

*Supplementary Figure 4.5 & 4.6: LocusZoom**^38^* *plot of genomic loci 5 and 6 - see extended data tables for genomic locus numbering.*

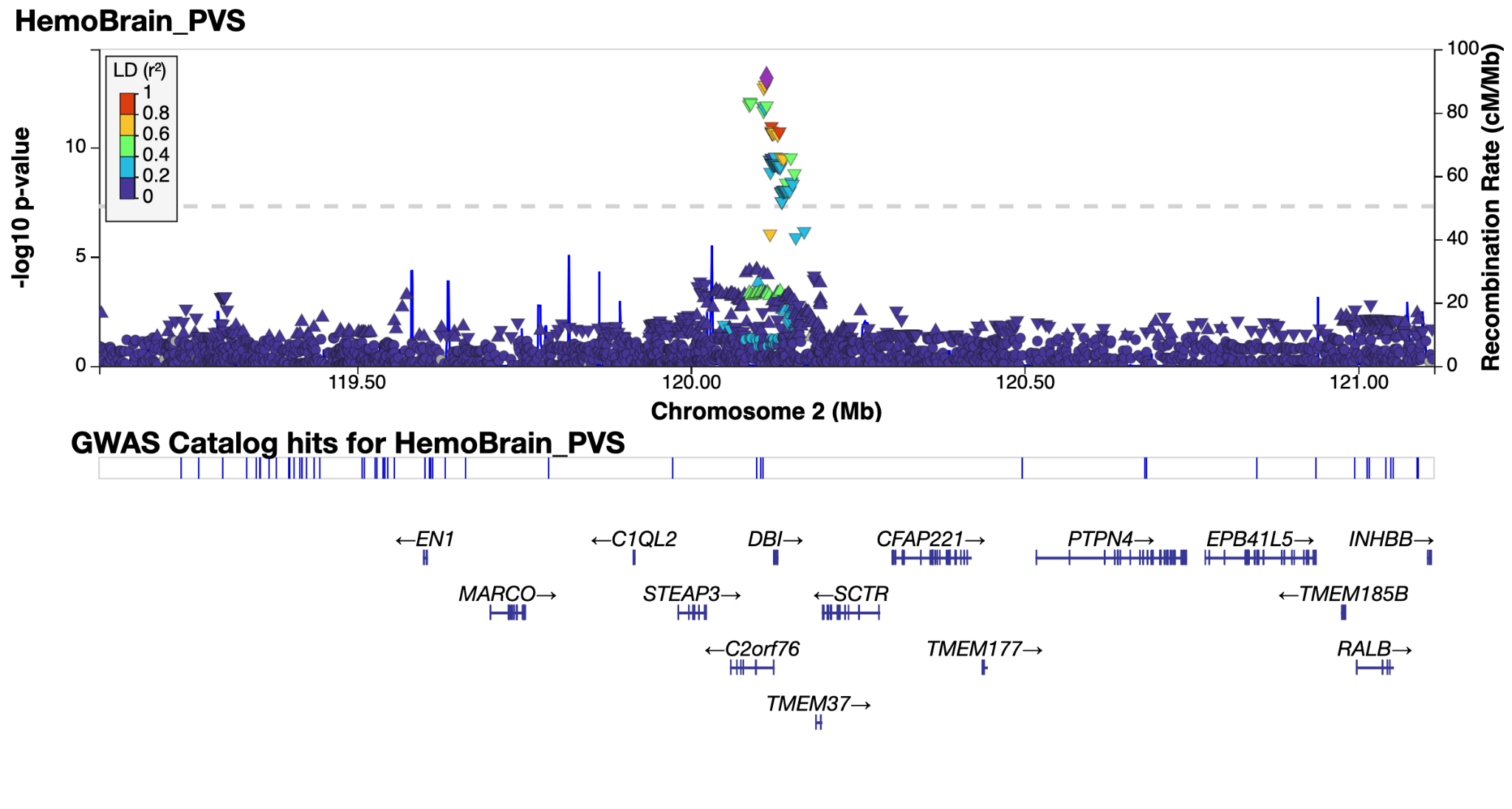

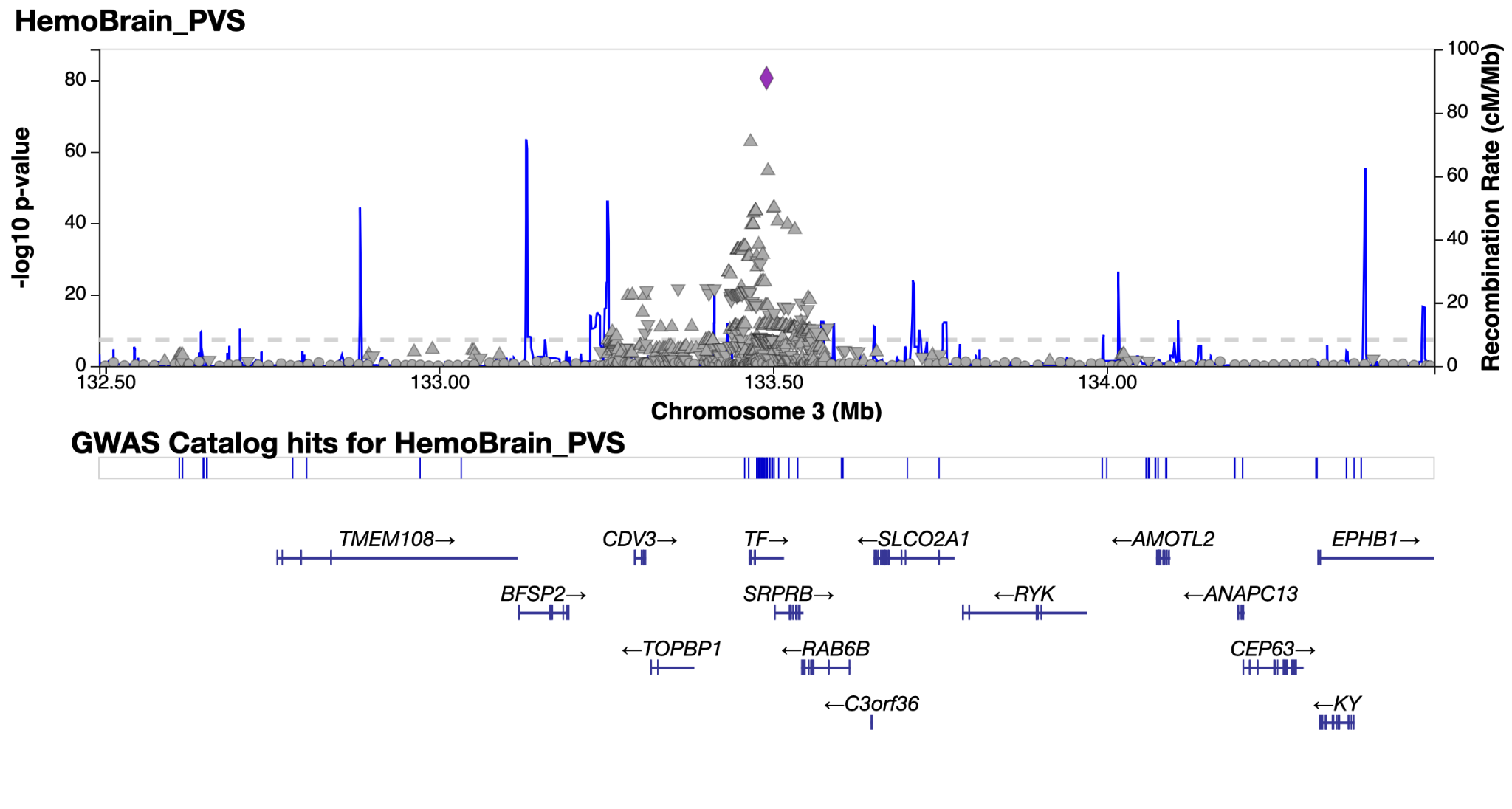

*Supplementary Figure 4.7 & 4.8: LocusZoom**^38^* *plot of genomic loci 7 and 8 - see extended data tables for genomic locus numbering. Non-lead SNP points are in gray as lead SNP is not in LD reference panel (1k genomes).*

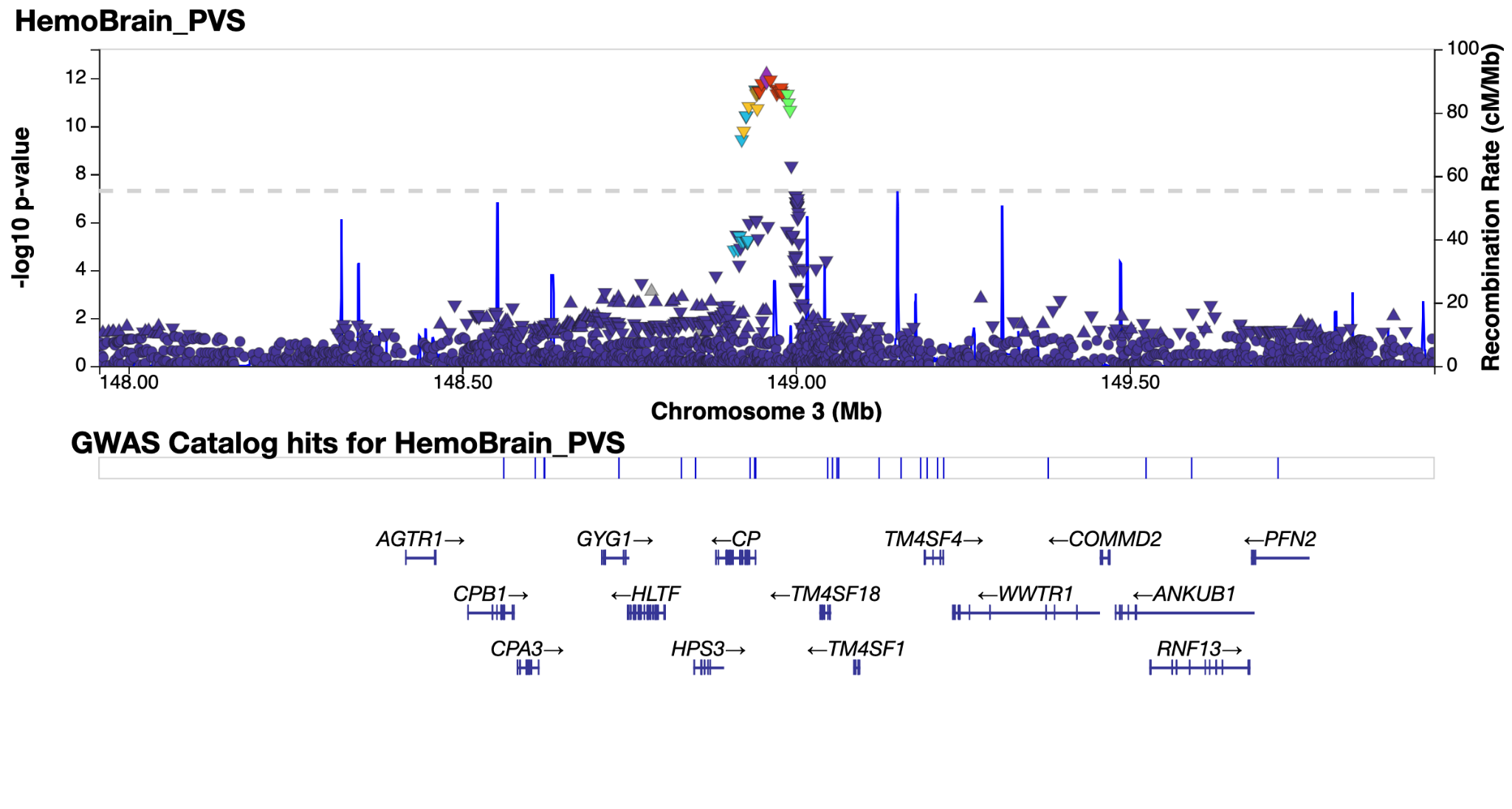

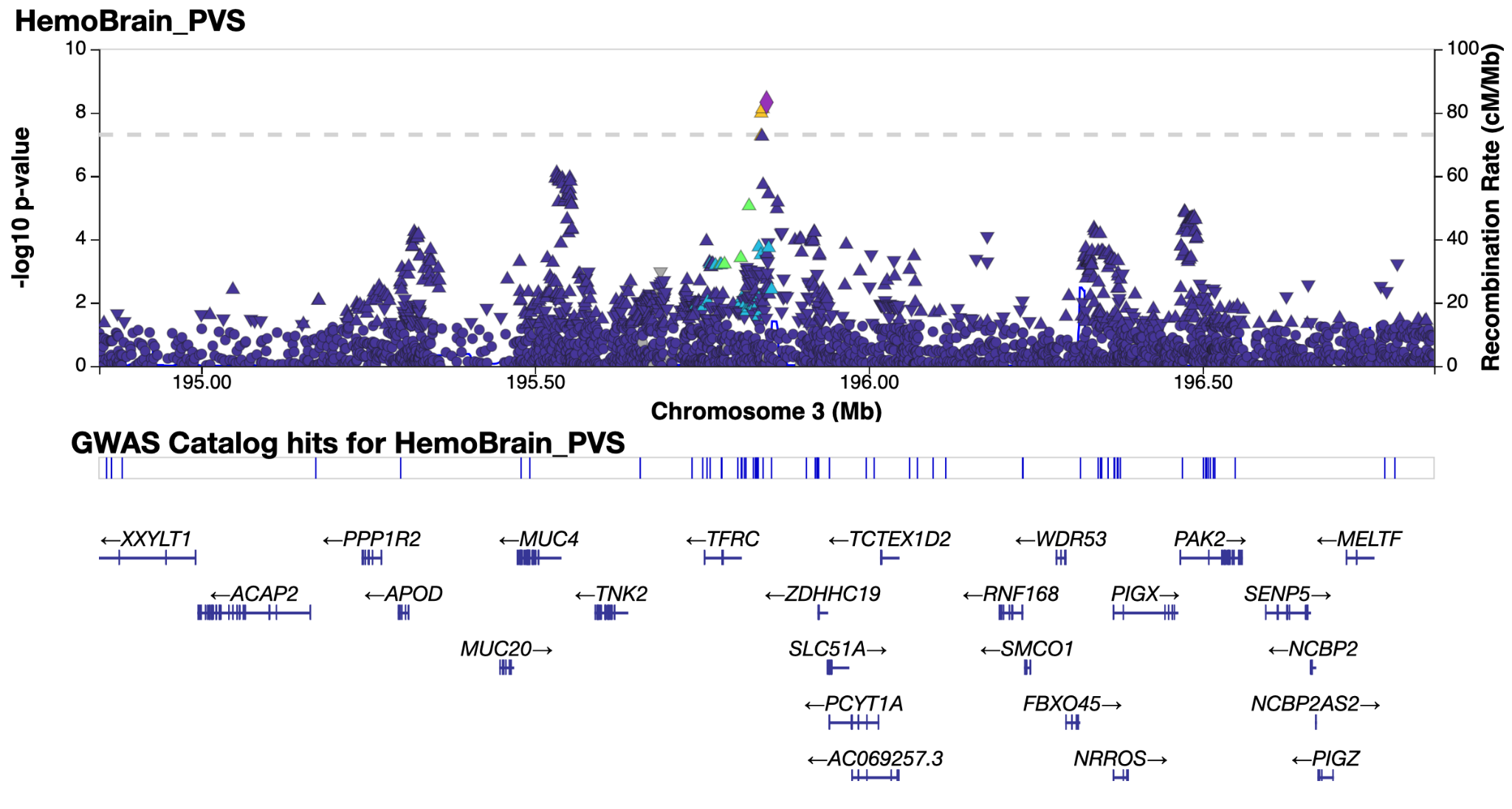

*Supplementary Figure 4.9 & 4.10: LocusZoom**^38^* *plot of genomic loci 9 and 10 - see extended data tables for genomic locus numbering.*

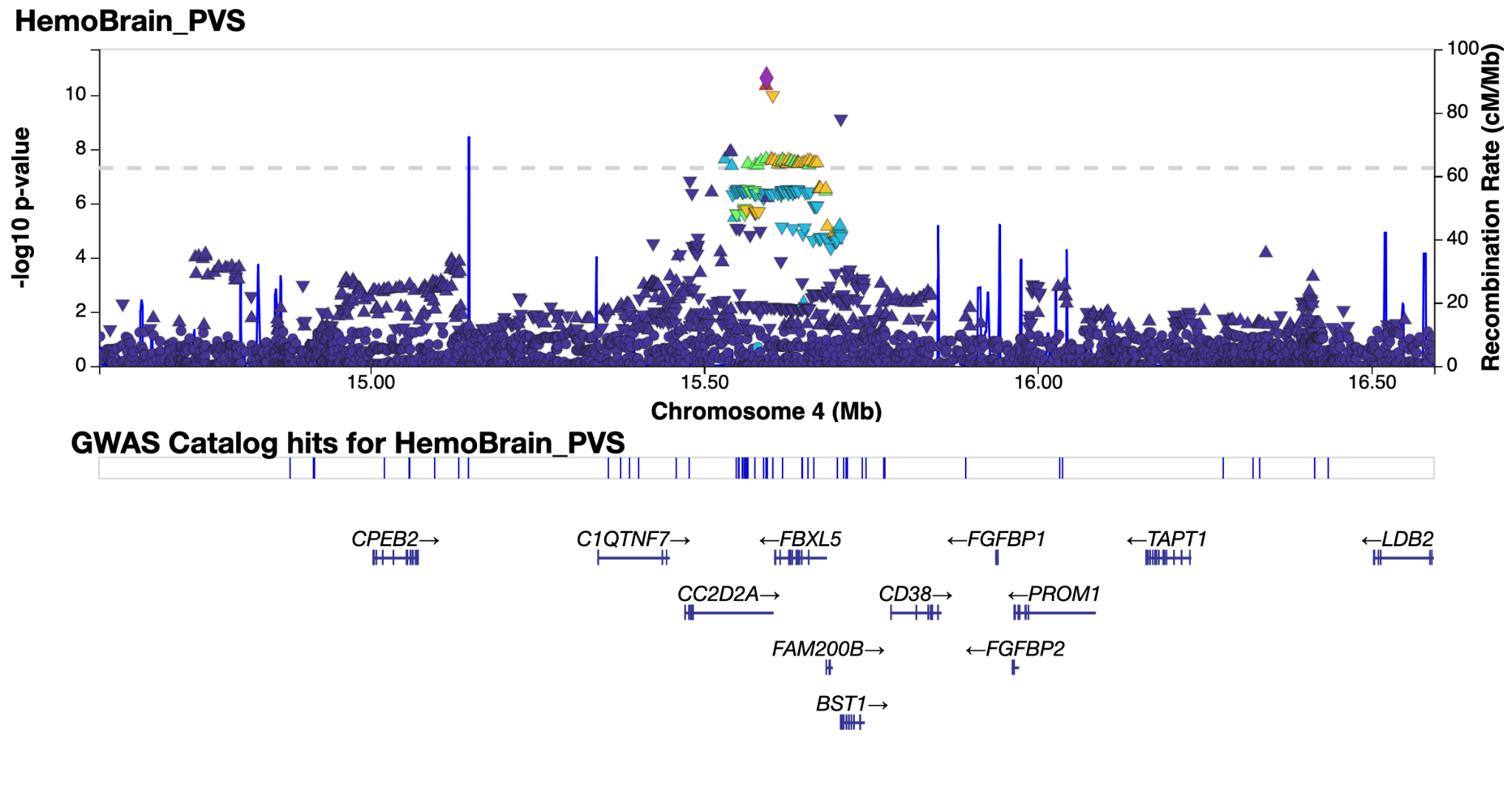

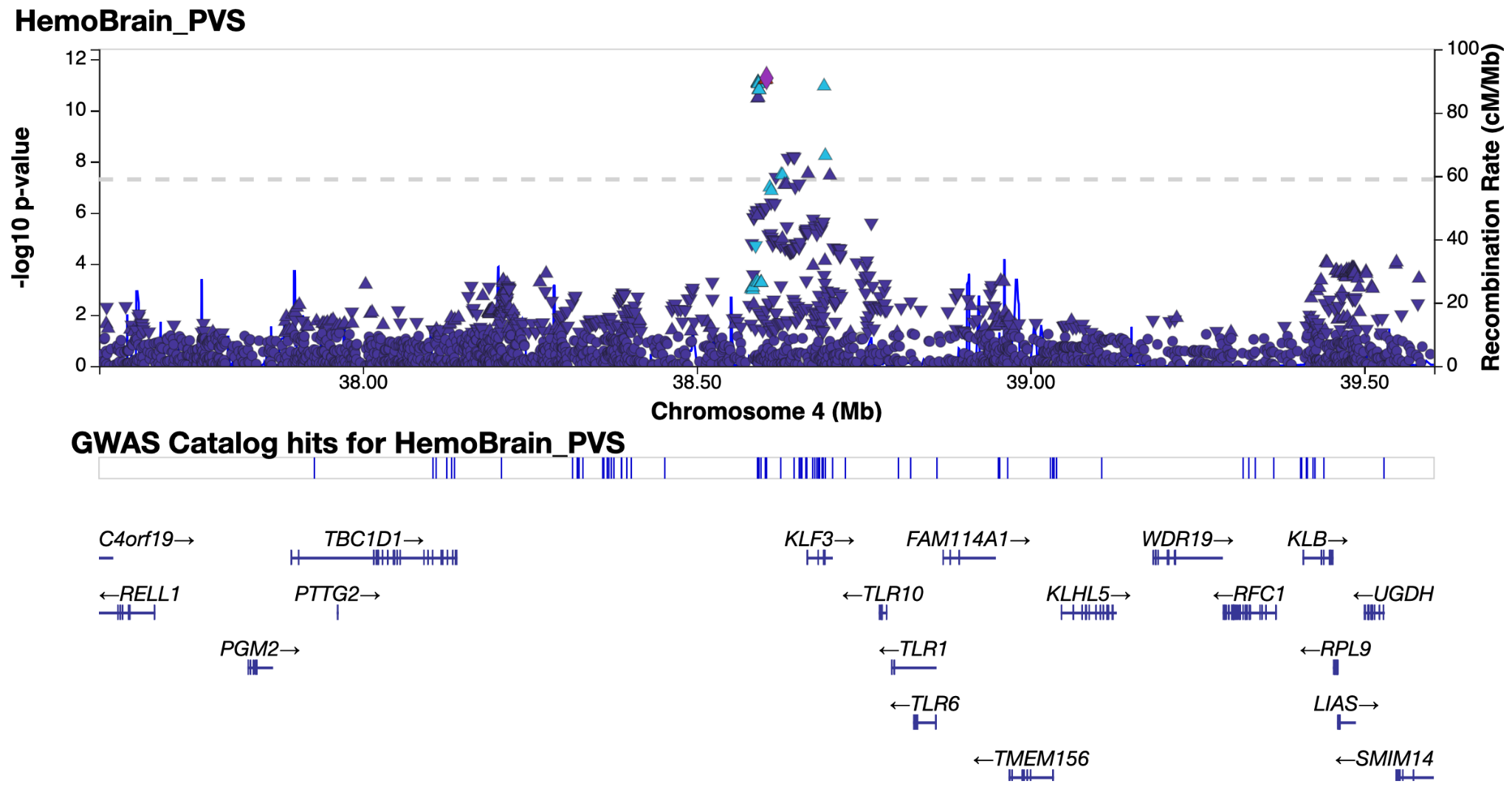

*Supplementary Figure 4.11 & 4.12: LocusZoom**^38^* *plot of genomic loci 11 and 12 - see extended data tables for genomic locus numbering.*

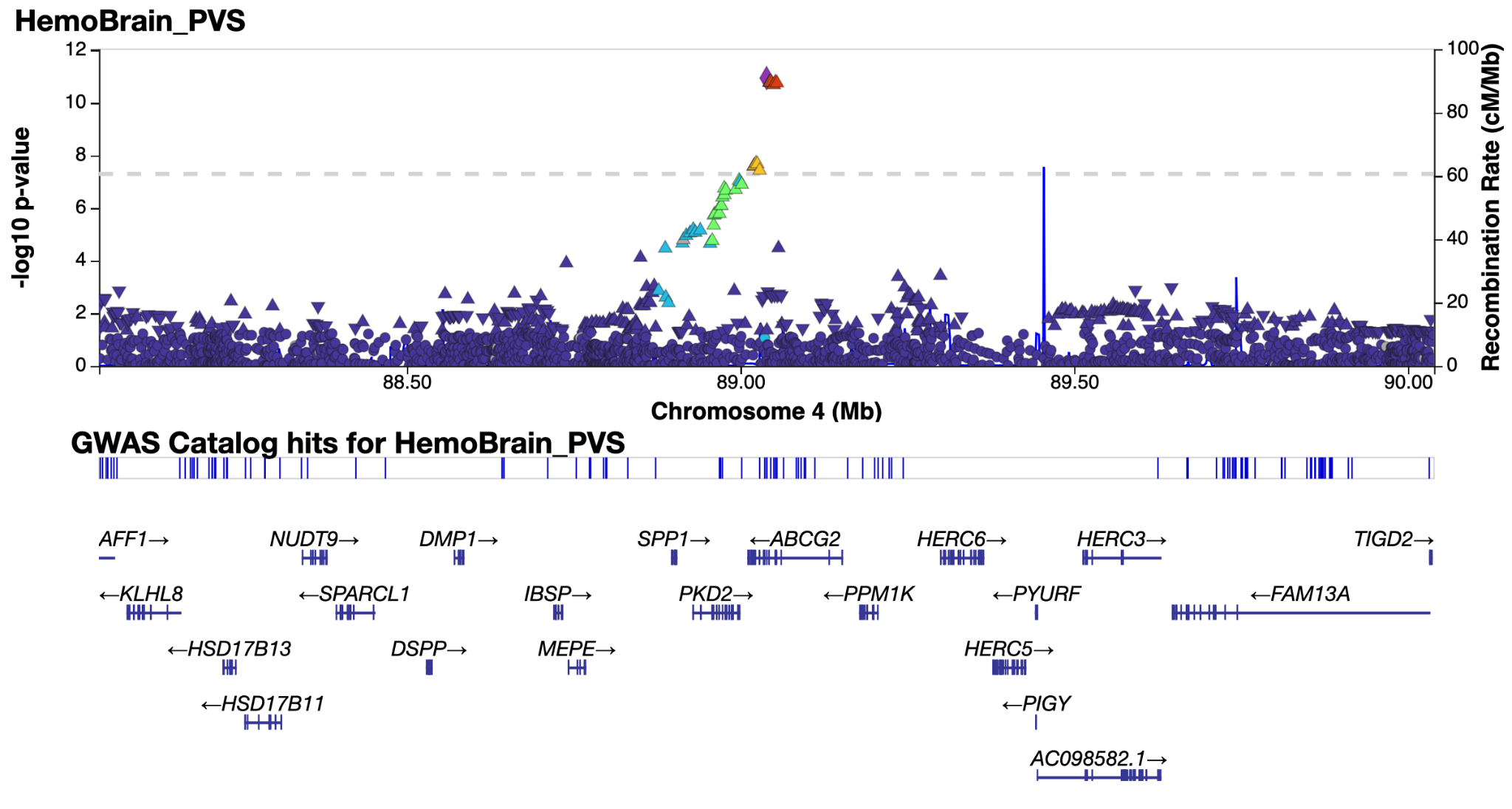

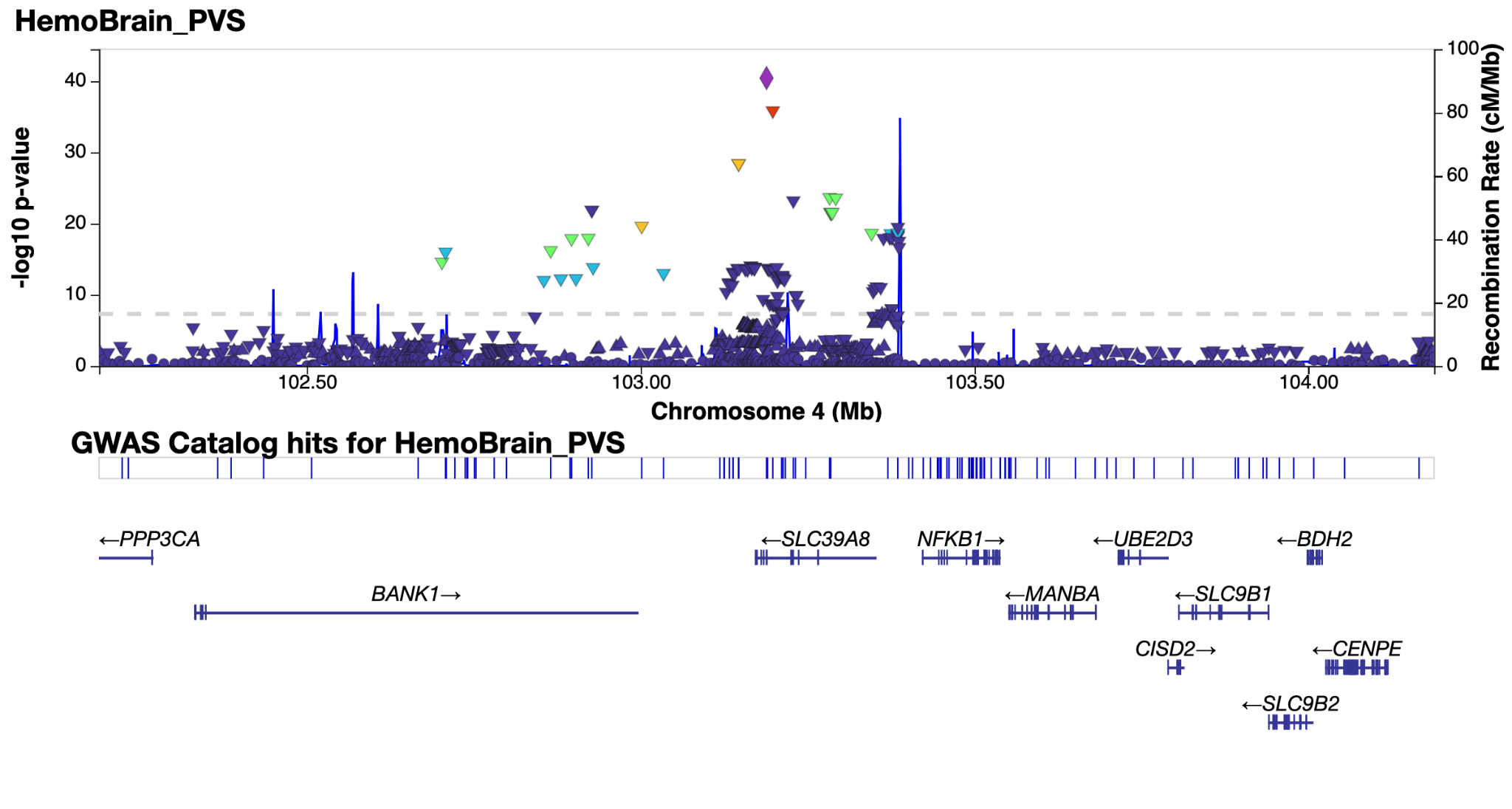

*Supplementary Figure 4.13 & 4.14: LocusZoom**^38^* *plot of genomic loci 13 and 14 - see extended data tables for genomic locus numbering.*

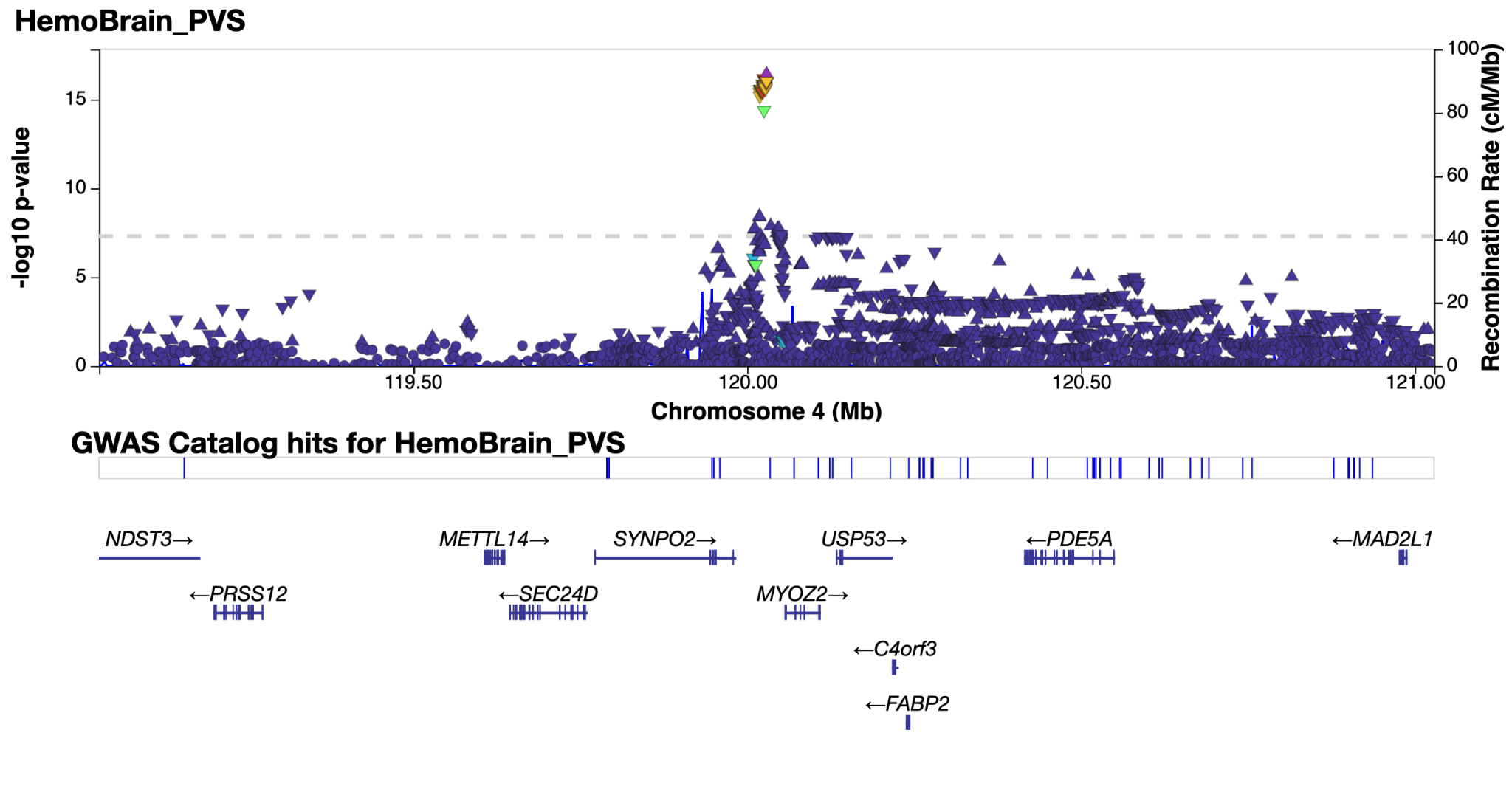

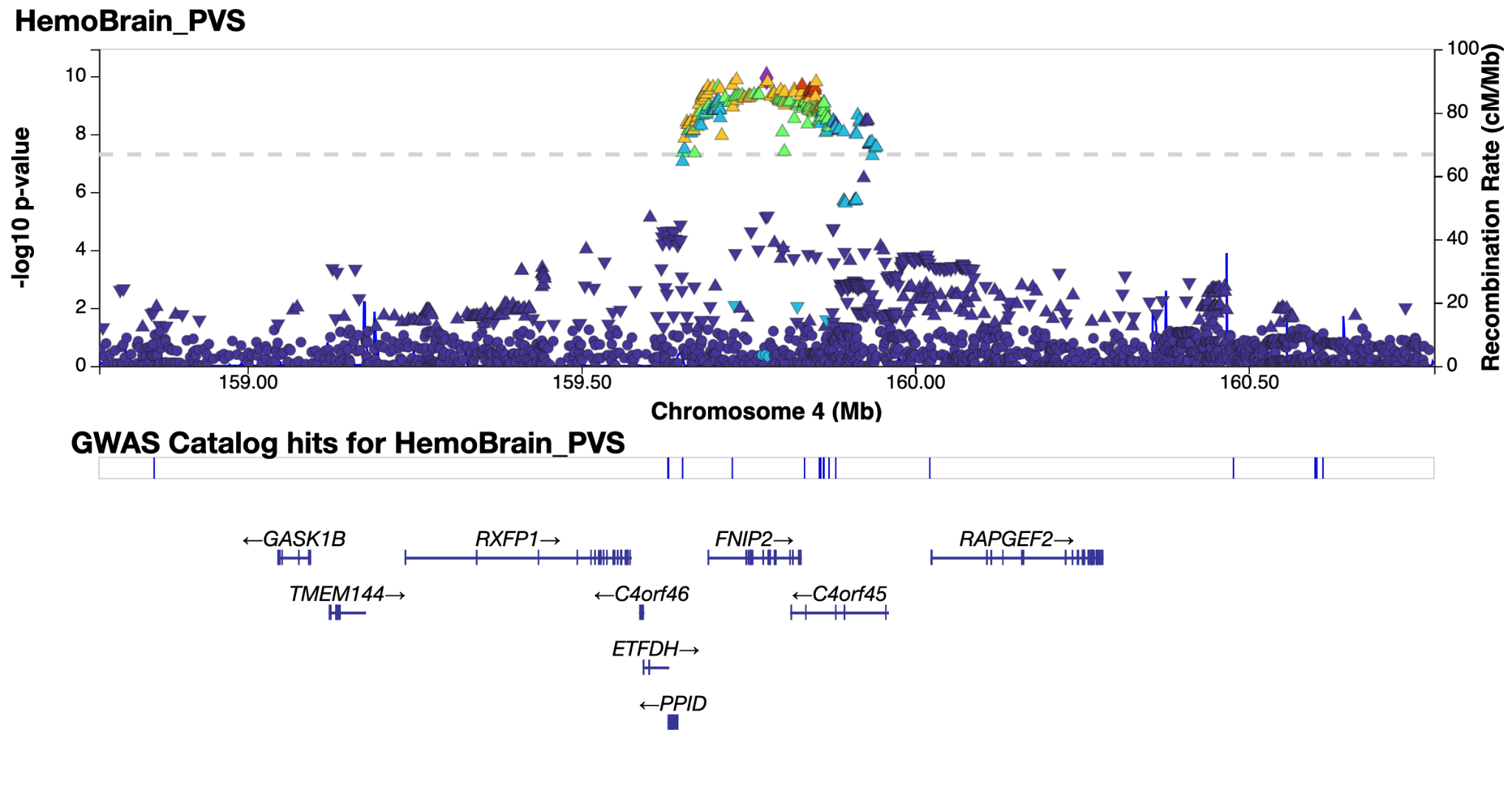

*Supplementary Figure 4.15 & 4.16: LocusZoom**^38^* *plot of genomic loci 15 and 16 - see extended data tables for genomic locus numbering.*

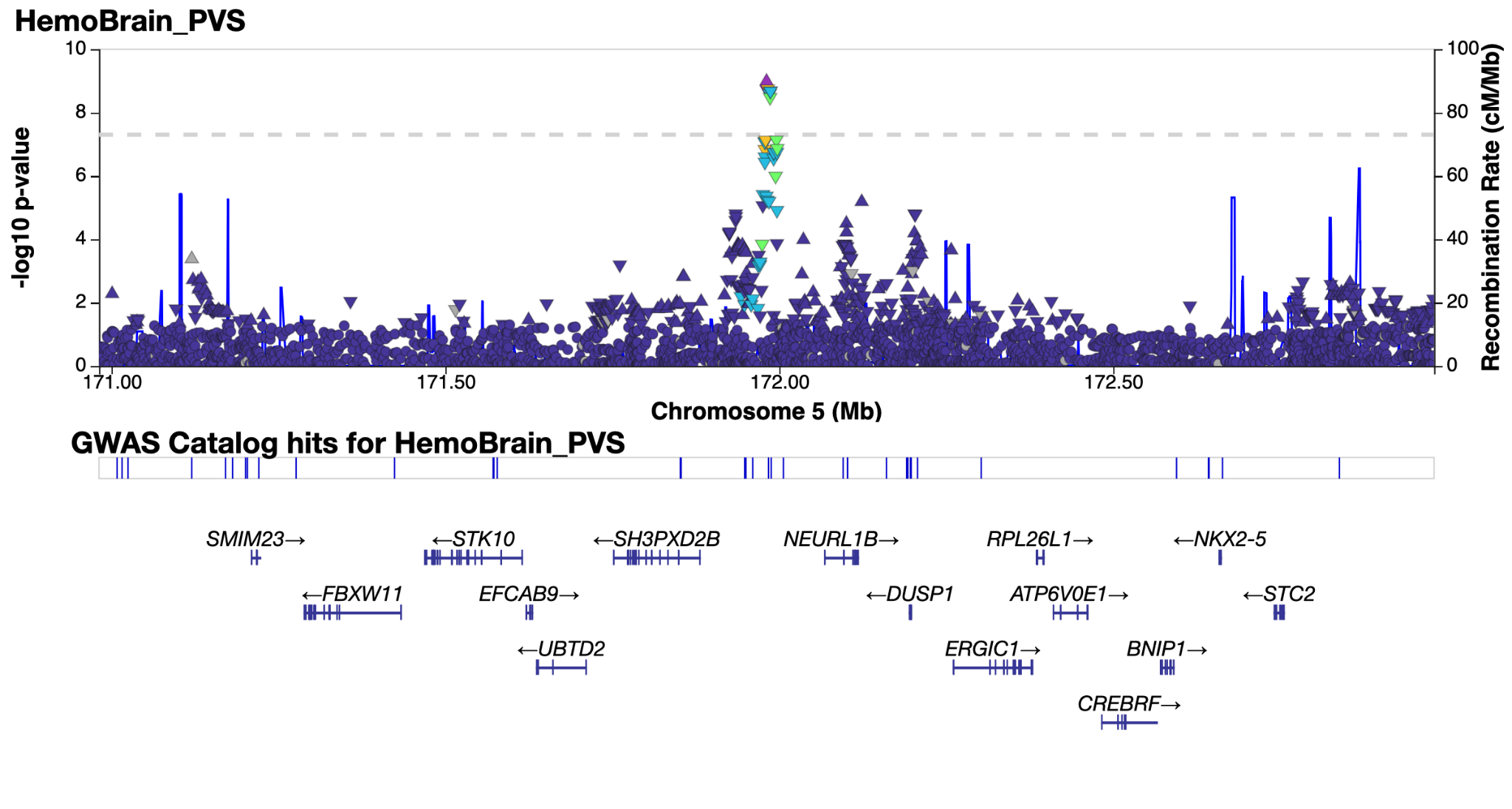

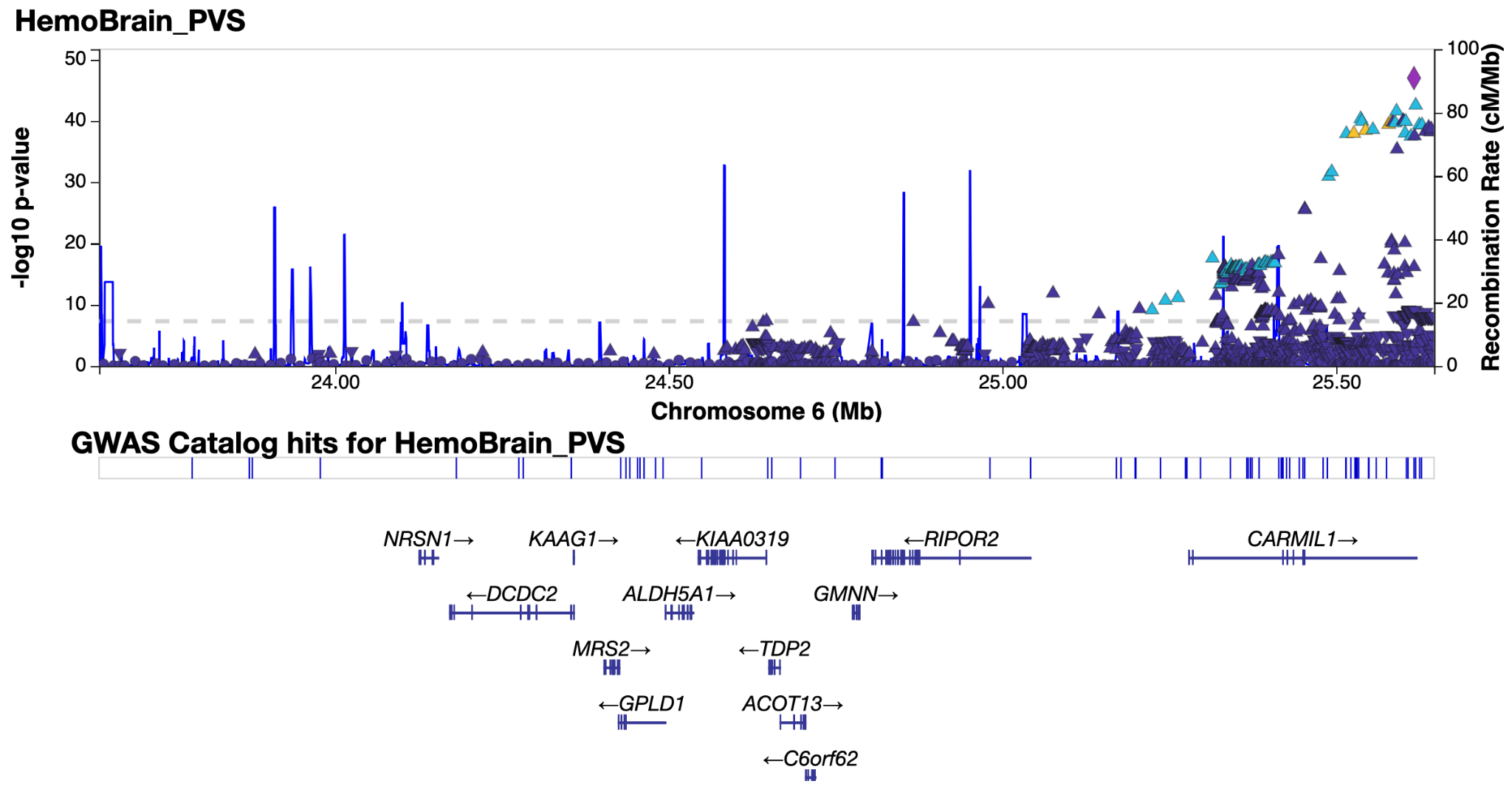

*Supplementary Figure 4.17 & 4.18: LocusZoom**^38^* *plot of genomic loci 17 and 18 - see extended data tables for genomic locus numbering.*

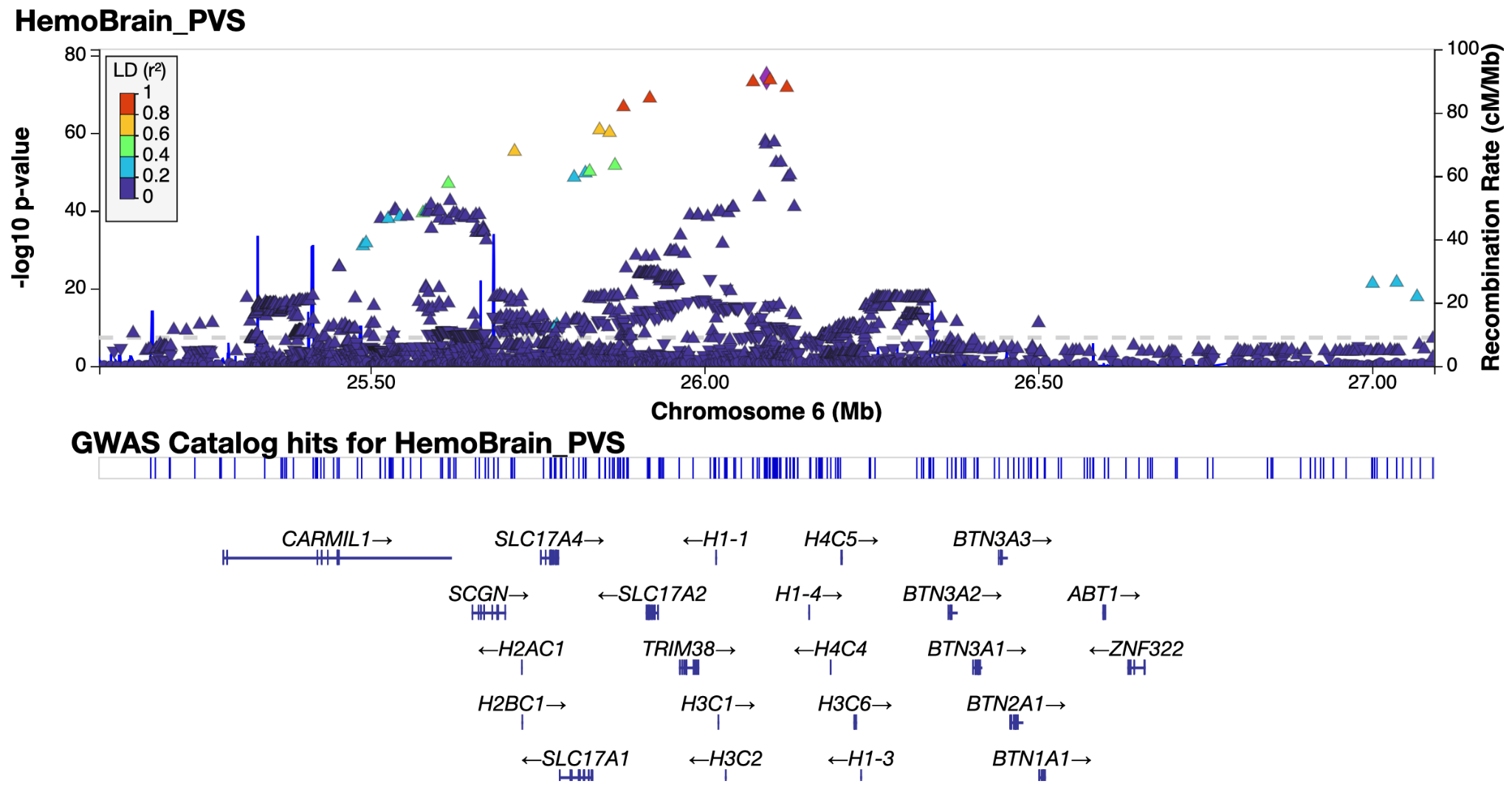

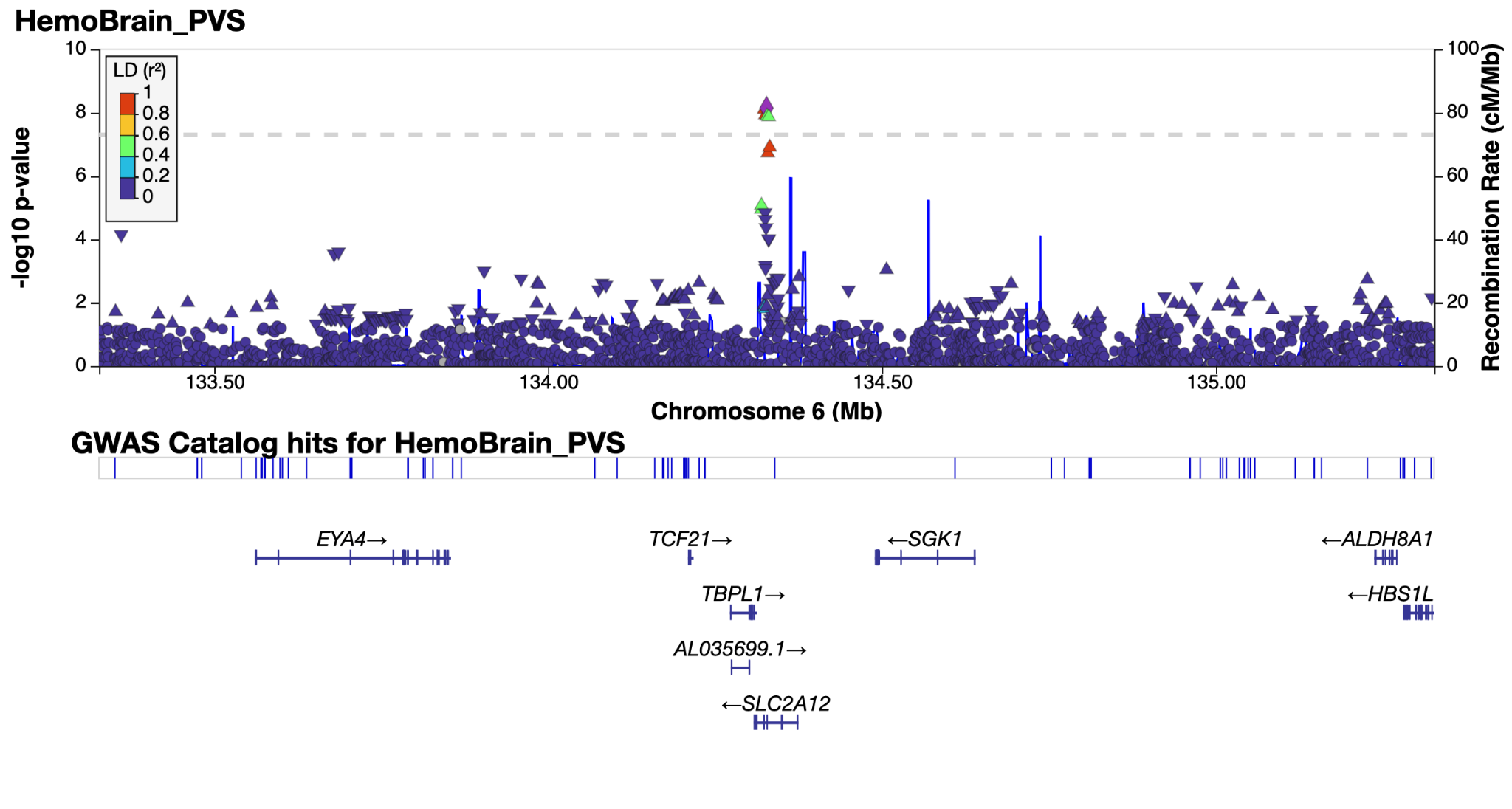

*Supplementary Figure 4.19 & 4.20: LocusZoom**^38^* *plot of genomic loci 19 and 20 - see extended data tables for genomic locus numbering.*

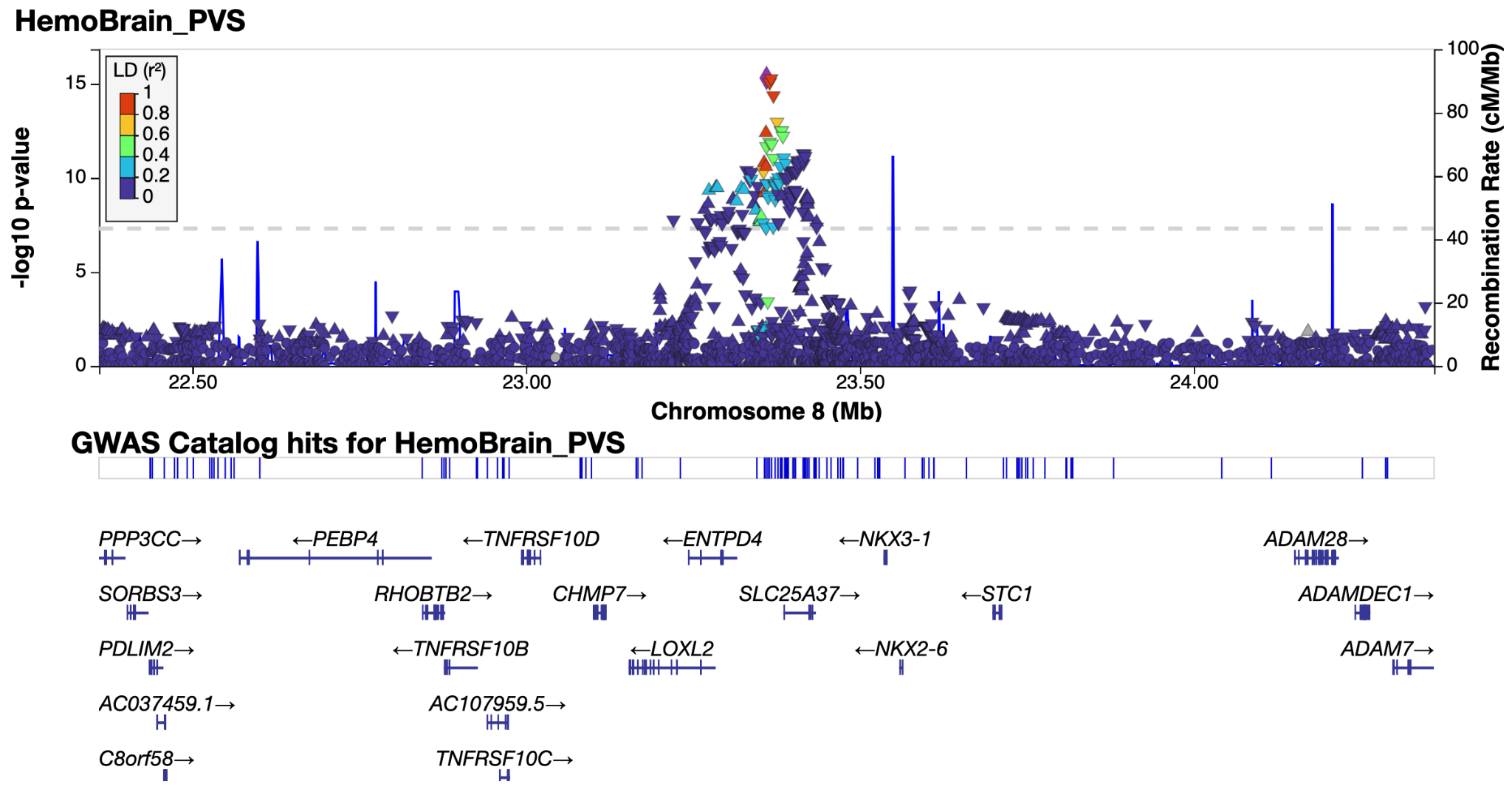

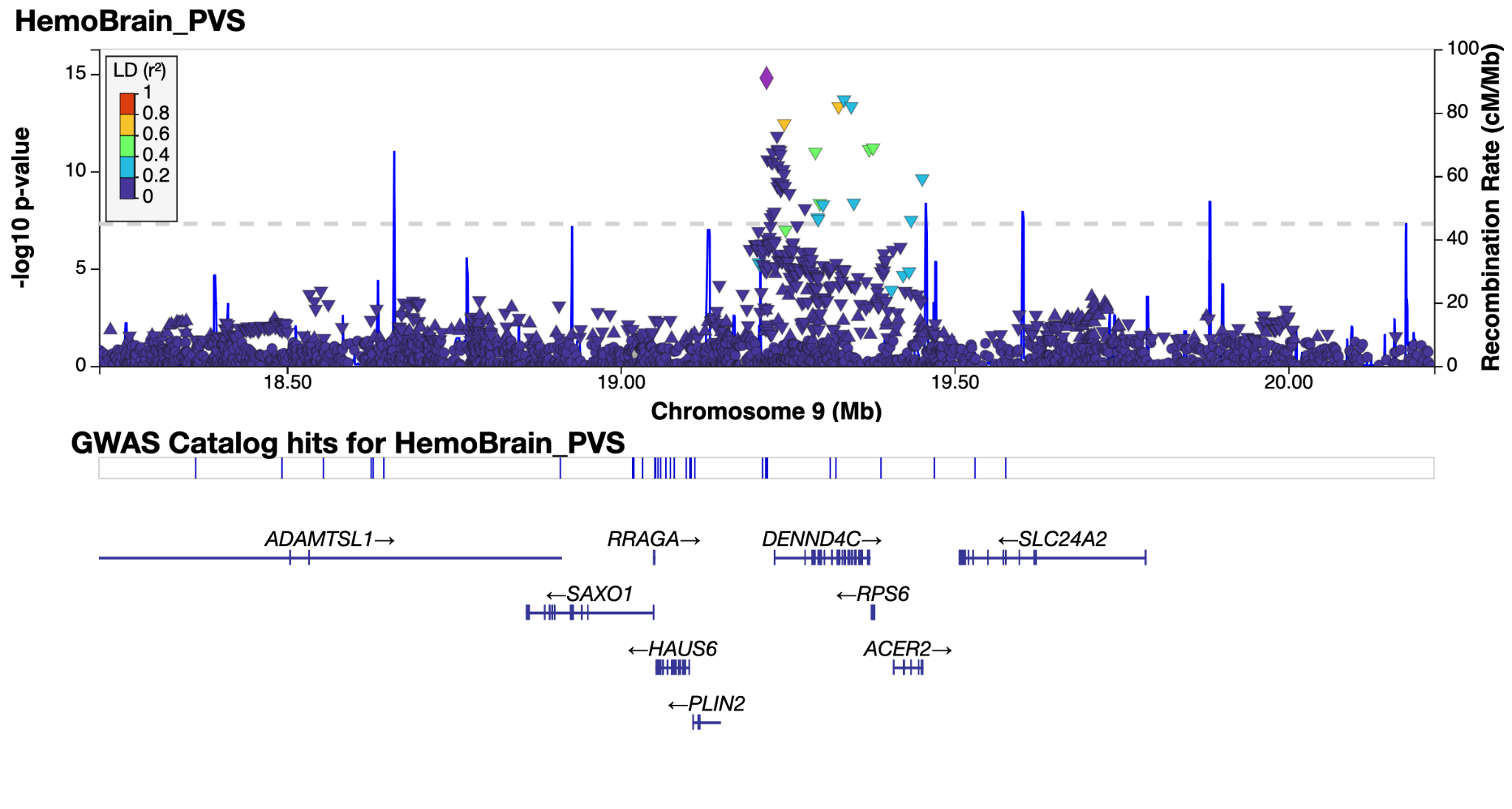

*Supplementary Figure 4.21 & 4.22: LocusZoom**^38^* *plot of genomic loci 21 and 22 - see extended data tables for genomic locus numbering.*

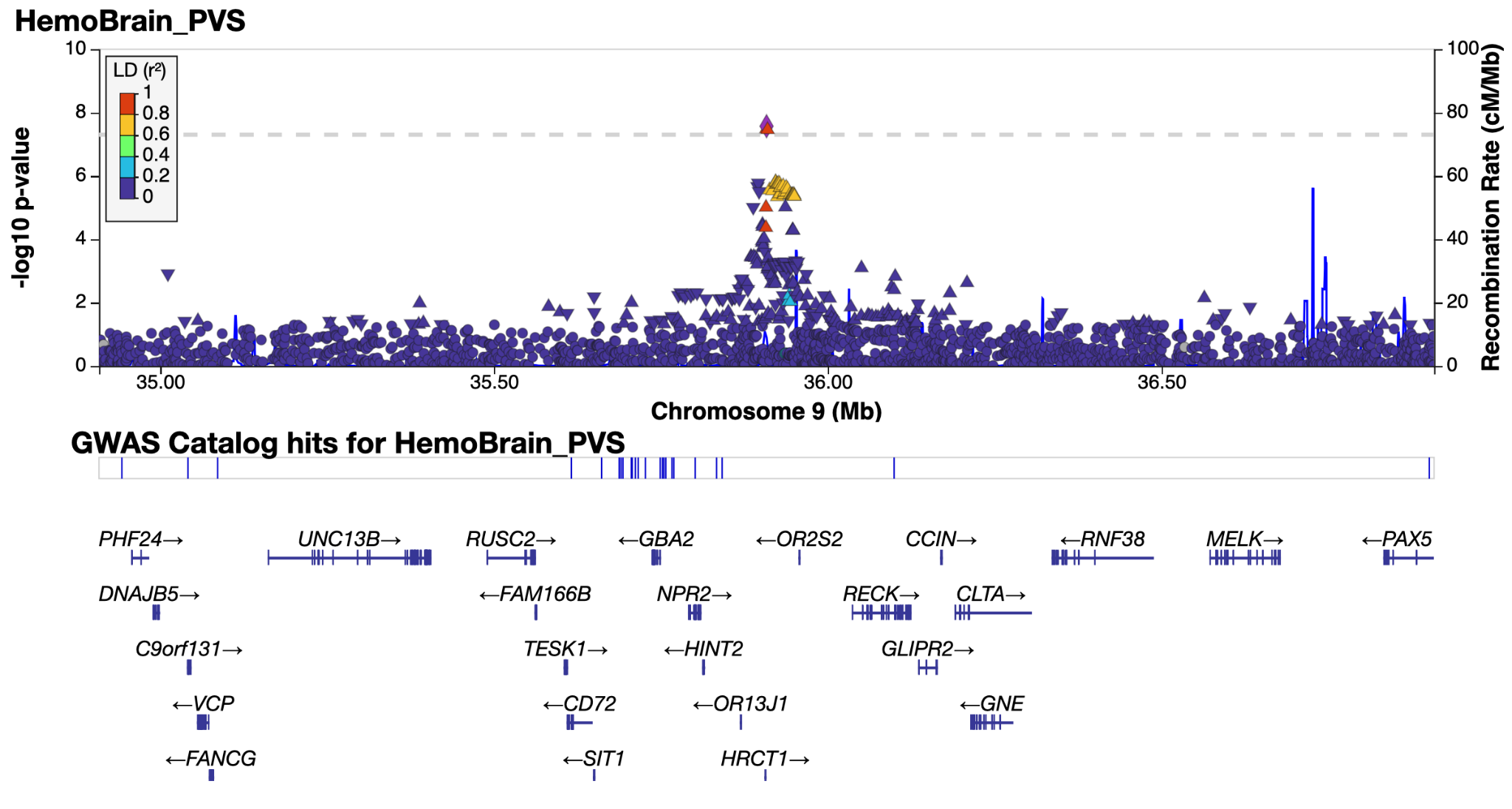

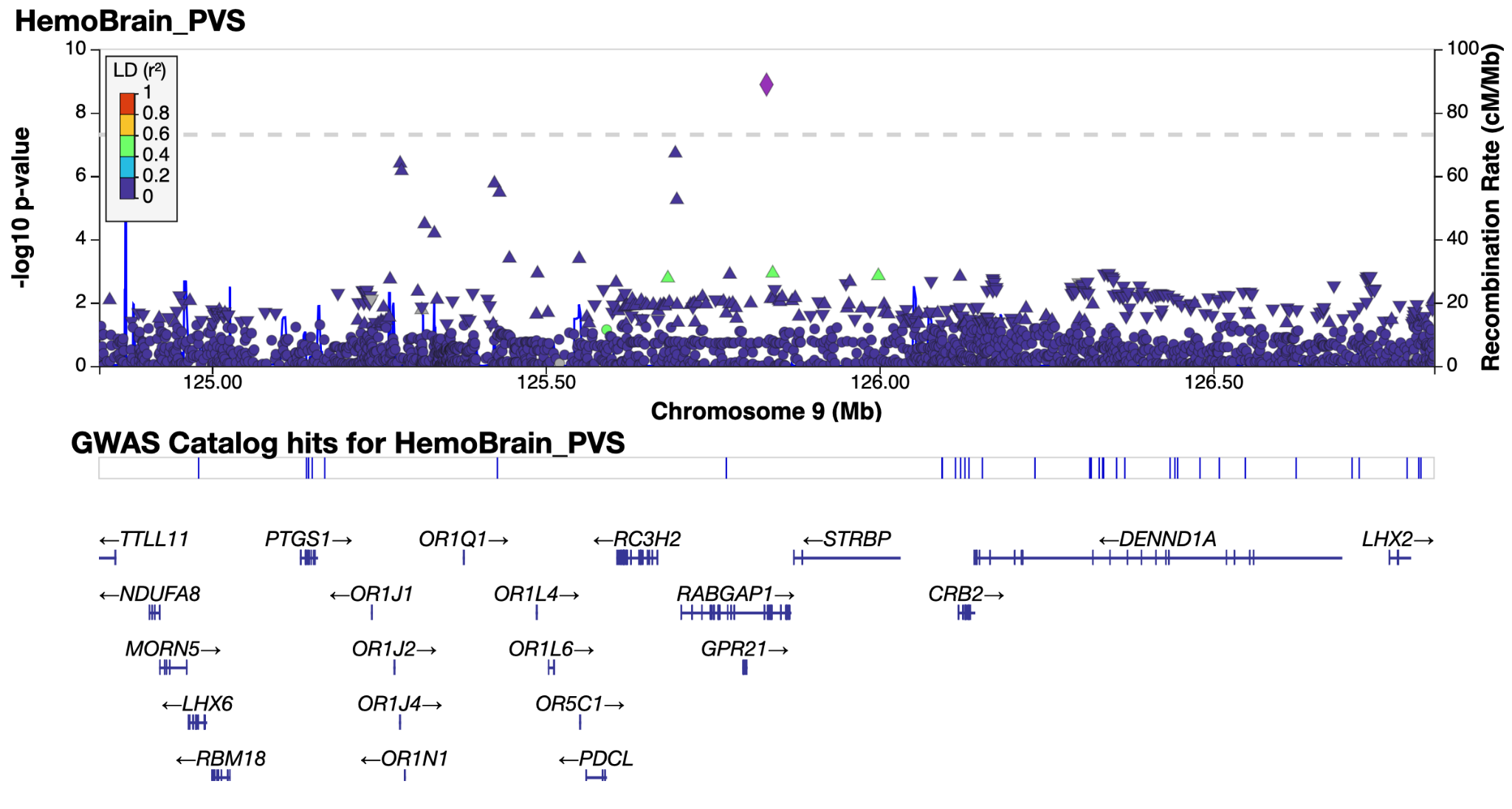

*Supplementary Figure 4.23 & 4.24: LocusZoom**^38^* *plot of genomic loci 23 and 24 - see extended data tables for genomic locus numbering. Locus 24 displayed a suspicious signal with nearby variants in LD, leading us to believe it was a spurious association and so was removed from the final locus count.*

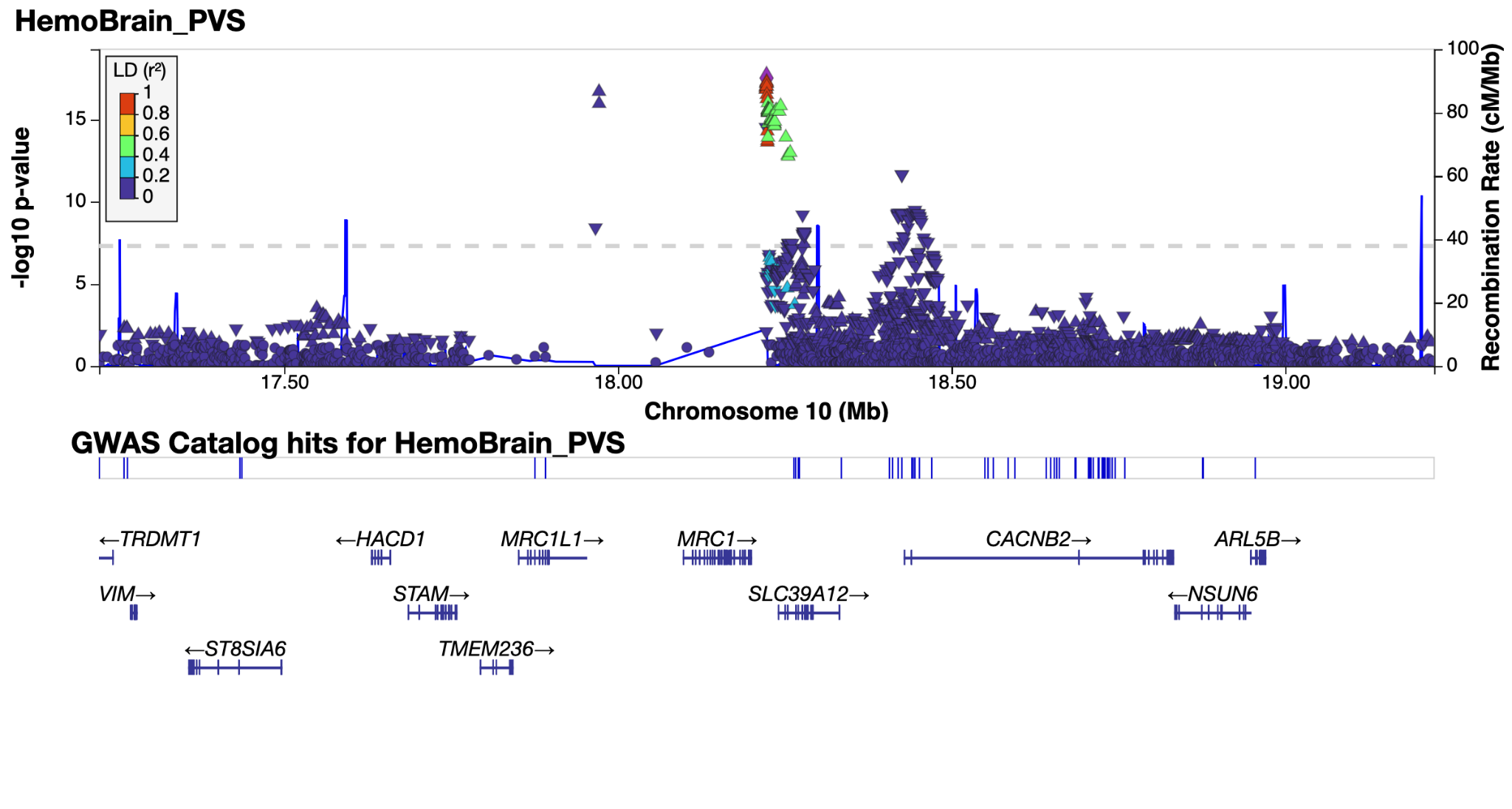

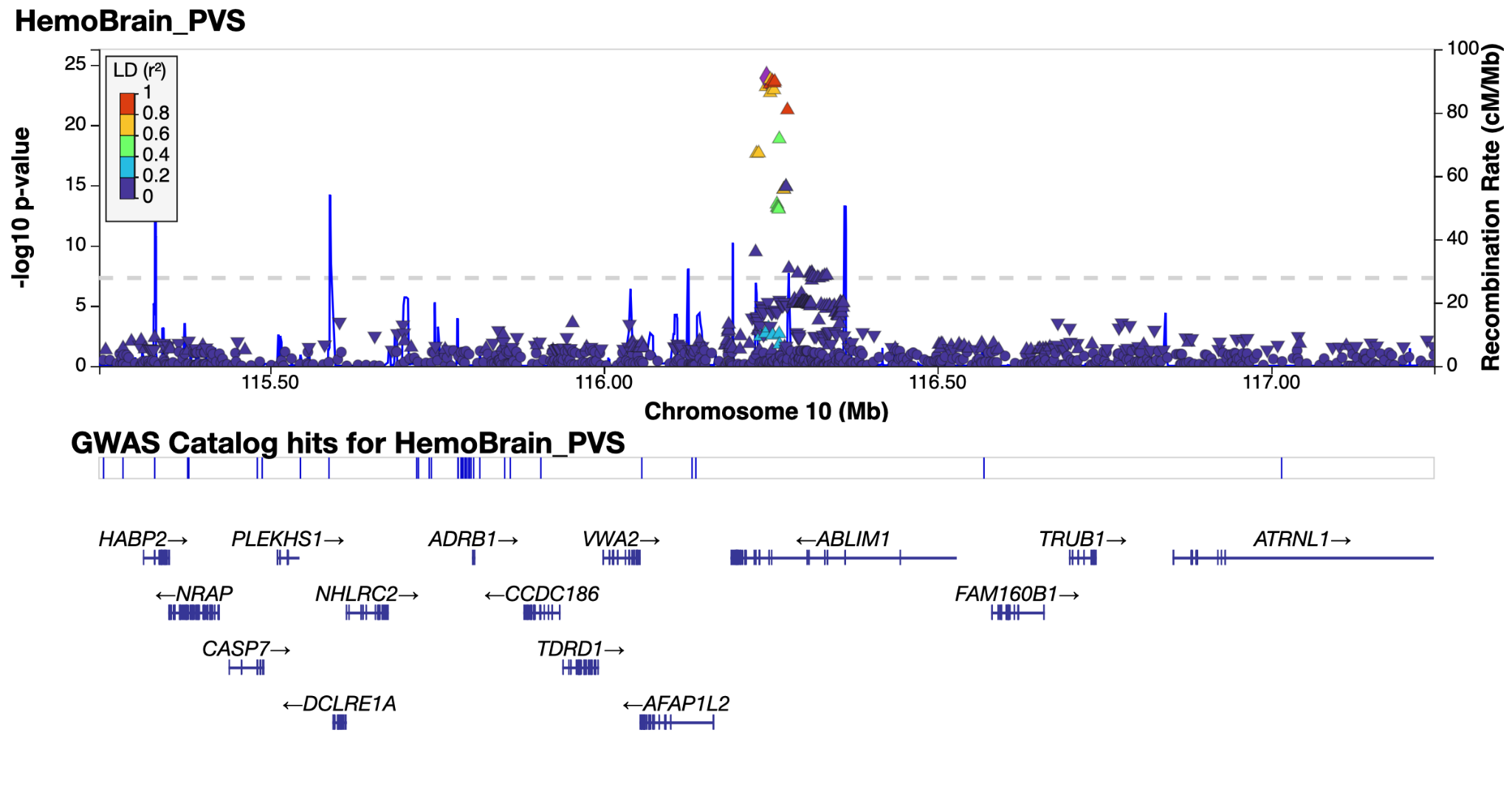

*Supplementary Figure 4.25 & 4.26: LocusZoom**^38^* *plot of genomic loci 25 and 26 - see extended data tables for genomic locus numbering.*

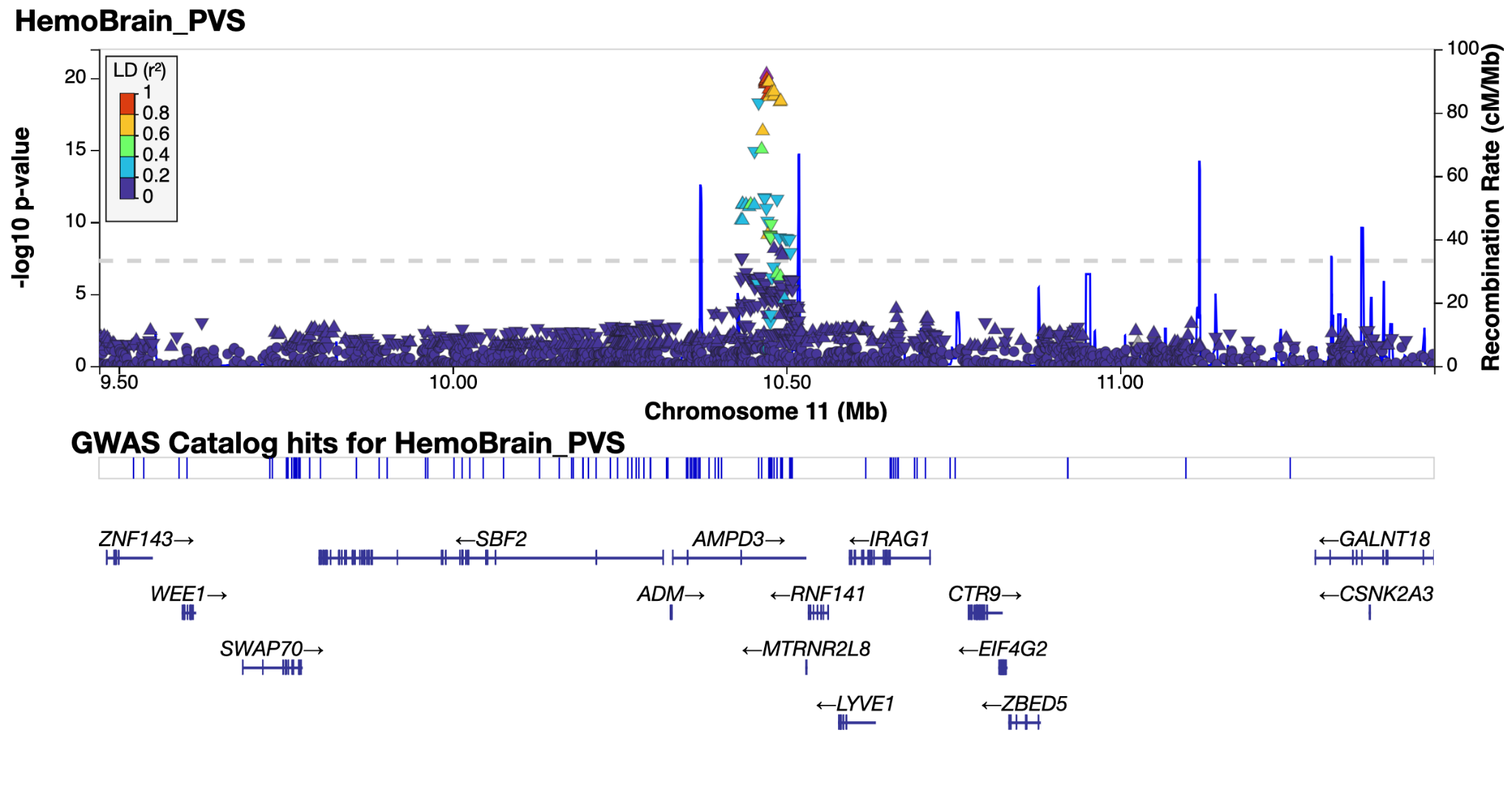

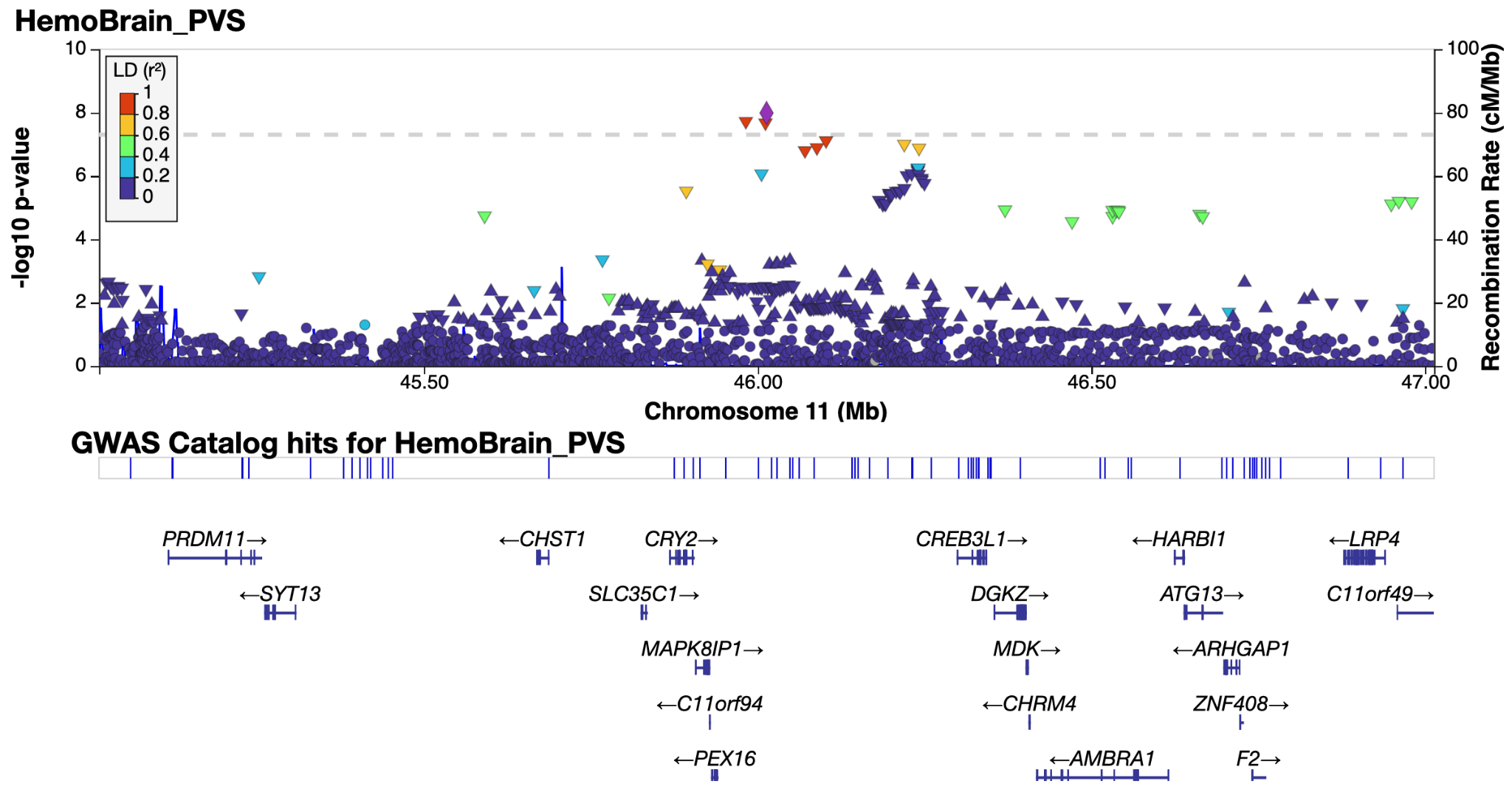

*Supplementary Figure 4.27 & 4.28: LocusZoom**^38^* *plot of genomic loci 27 and 28 - see extended data tables for genomic locus numbering.*

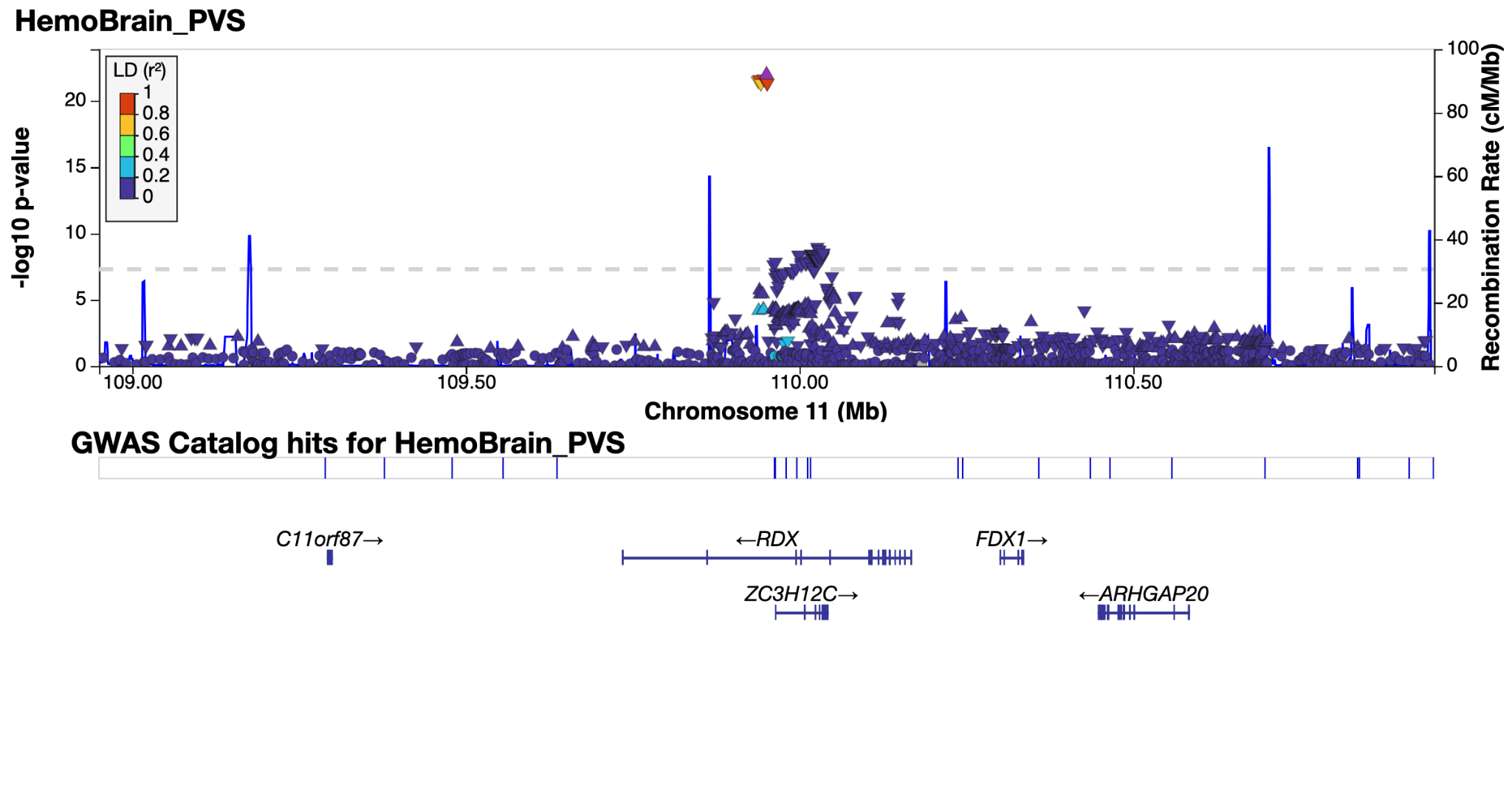

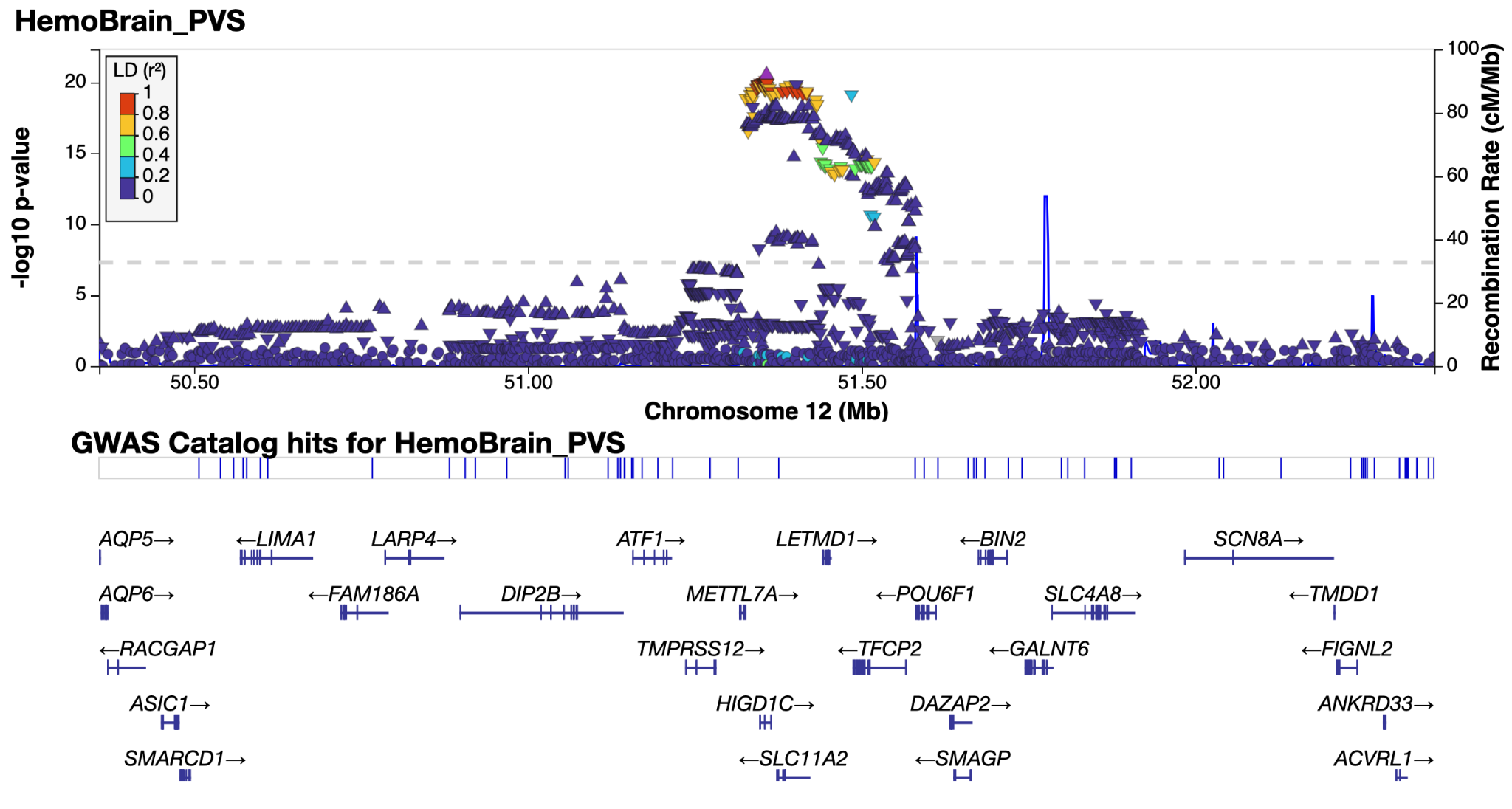

*Supplementary Figure 4.29 & 4.30: LocusZoom**^38^* *plot of genomic loci 29 and 30 - see extended data tables for genomic locus numbering.*

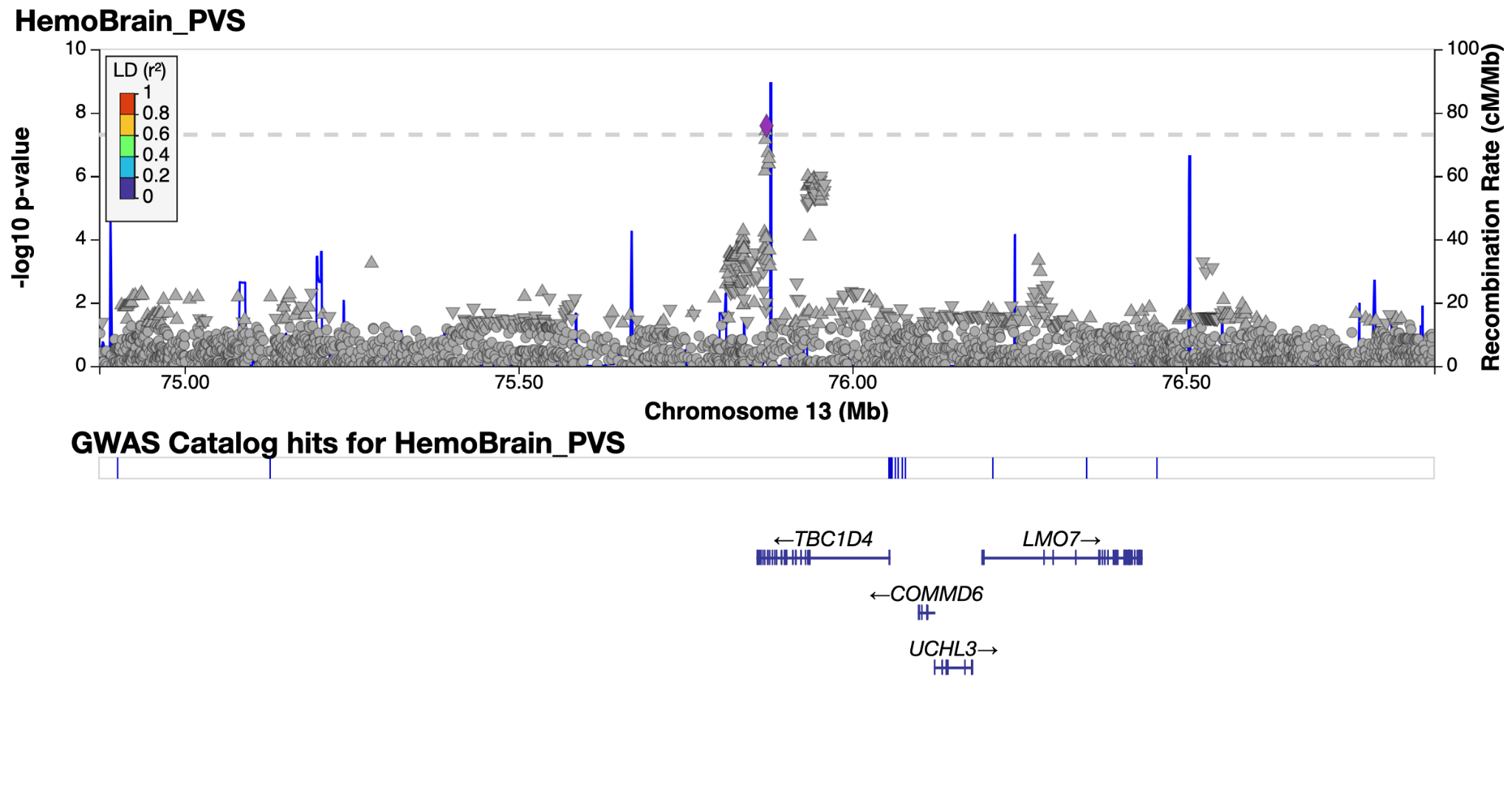

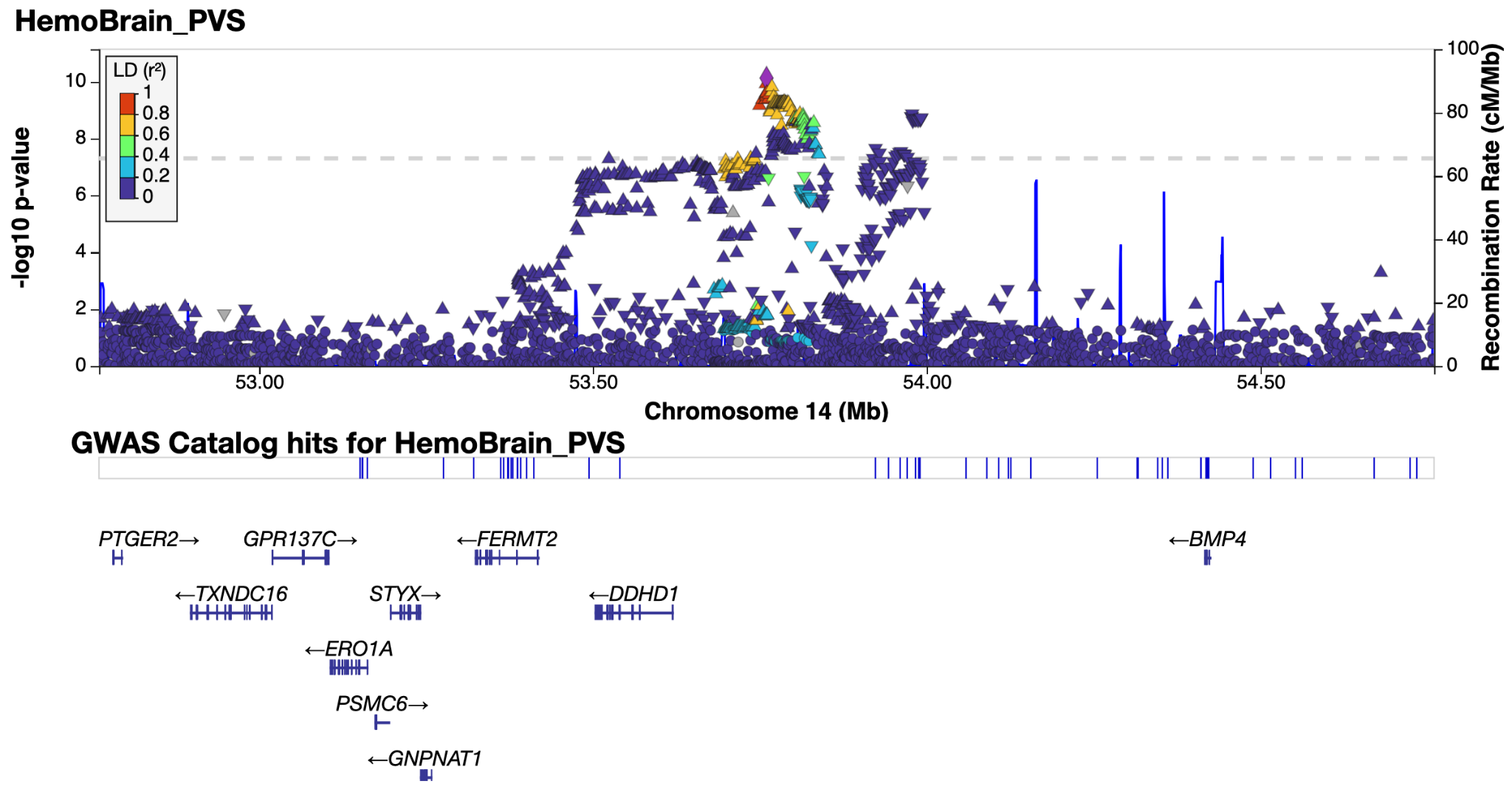

*Supplementary Figure 4.31 & 4.32: LocusZoom**^38^* *plot of genomic loci 31 and 32 - see extended data tables for genomic locus numbering. Non-lead SNP points are in gray as lead SNP is not in LD reference panel (1k genomes).*

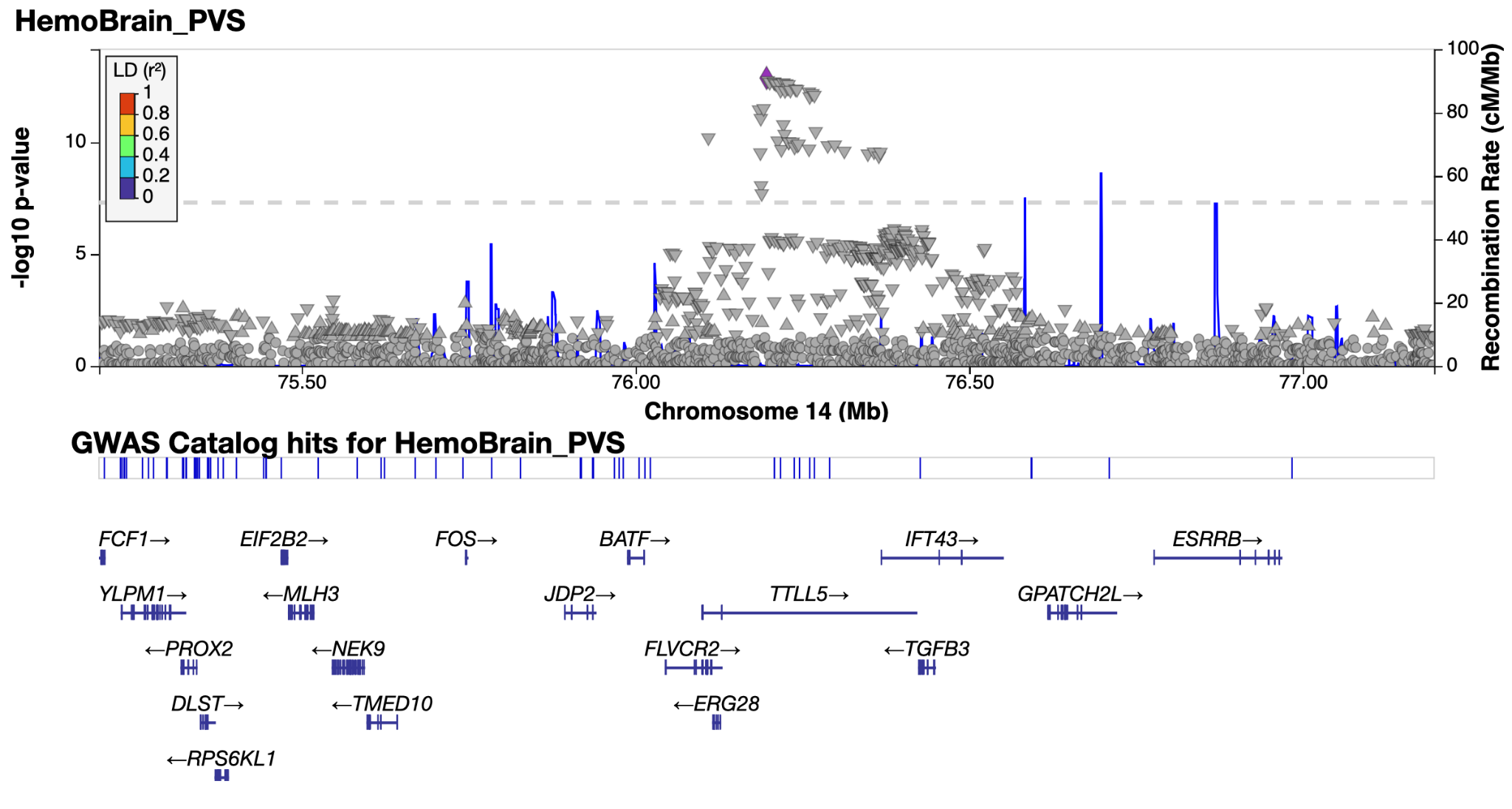

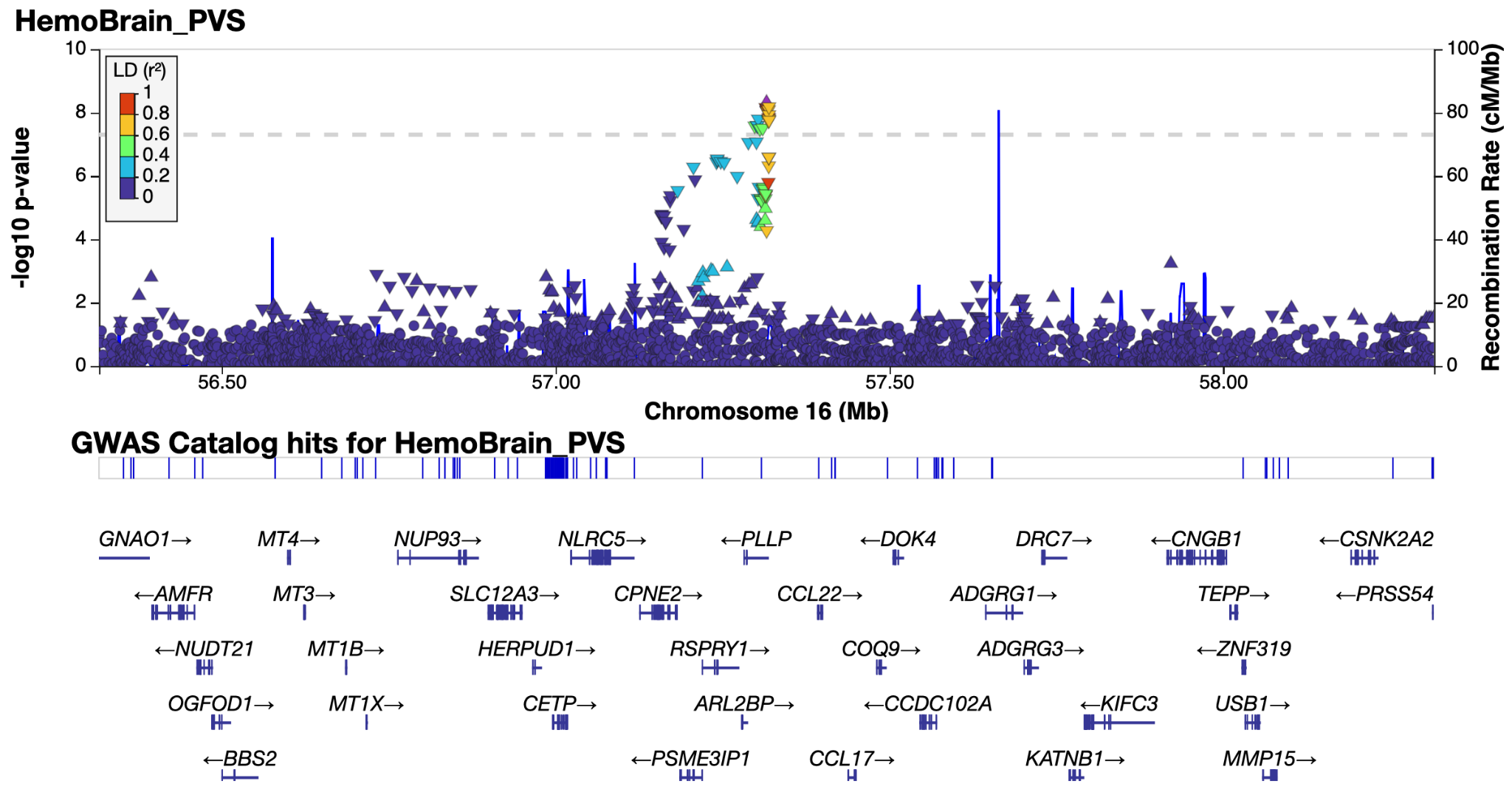

*Supplementary Figure 4.33 & 4.34: LocusZoom**^38^* *plot of genomic loci 33 and 34 - see extended data tables for genomic locus numbering. Non-lead SNP points are in gray as lead SNP is not in LD reference panel (1k genomes).*

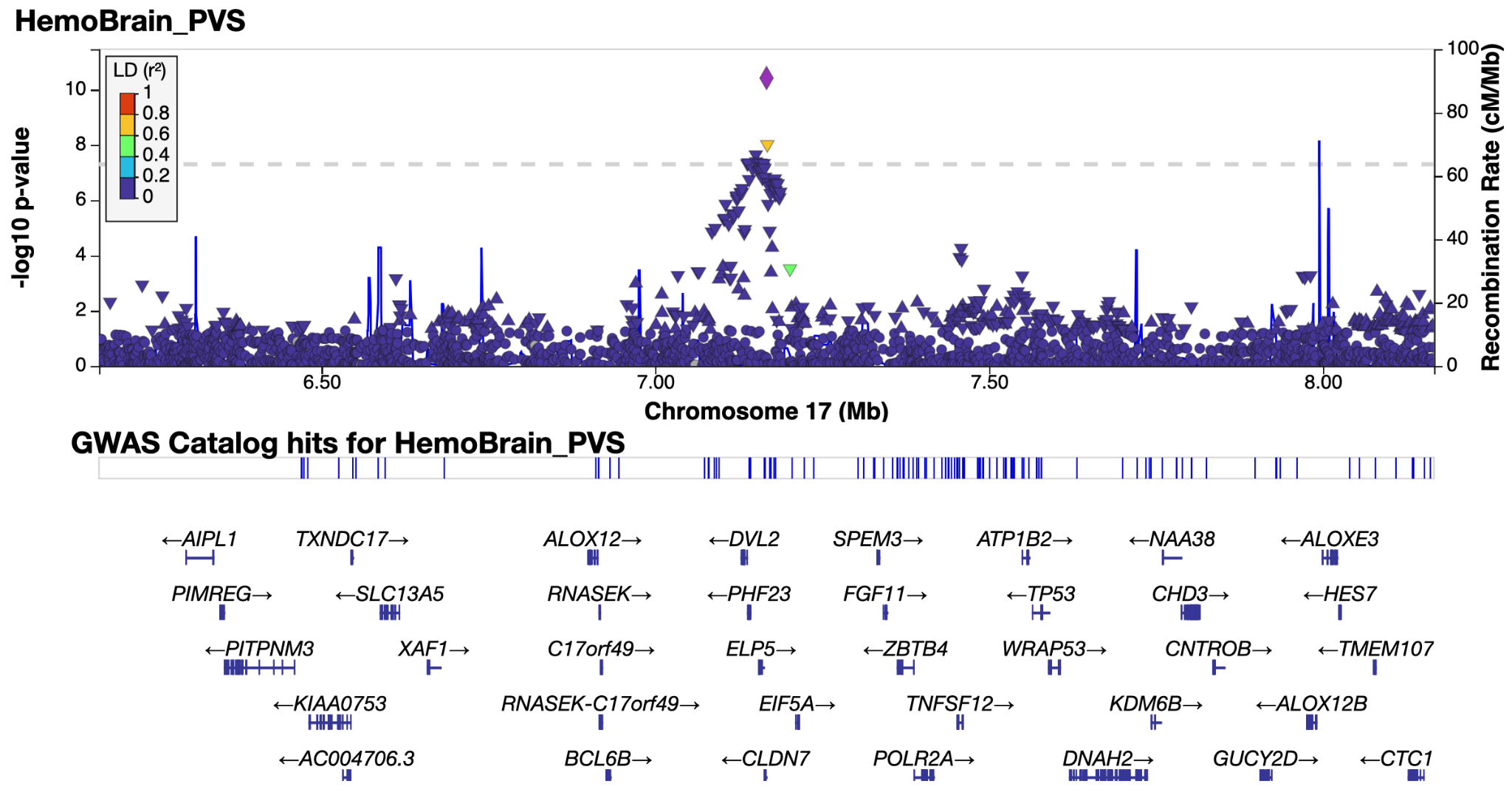

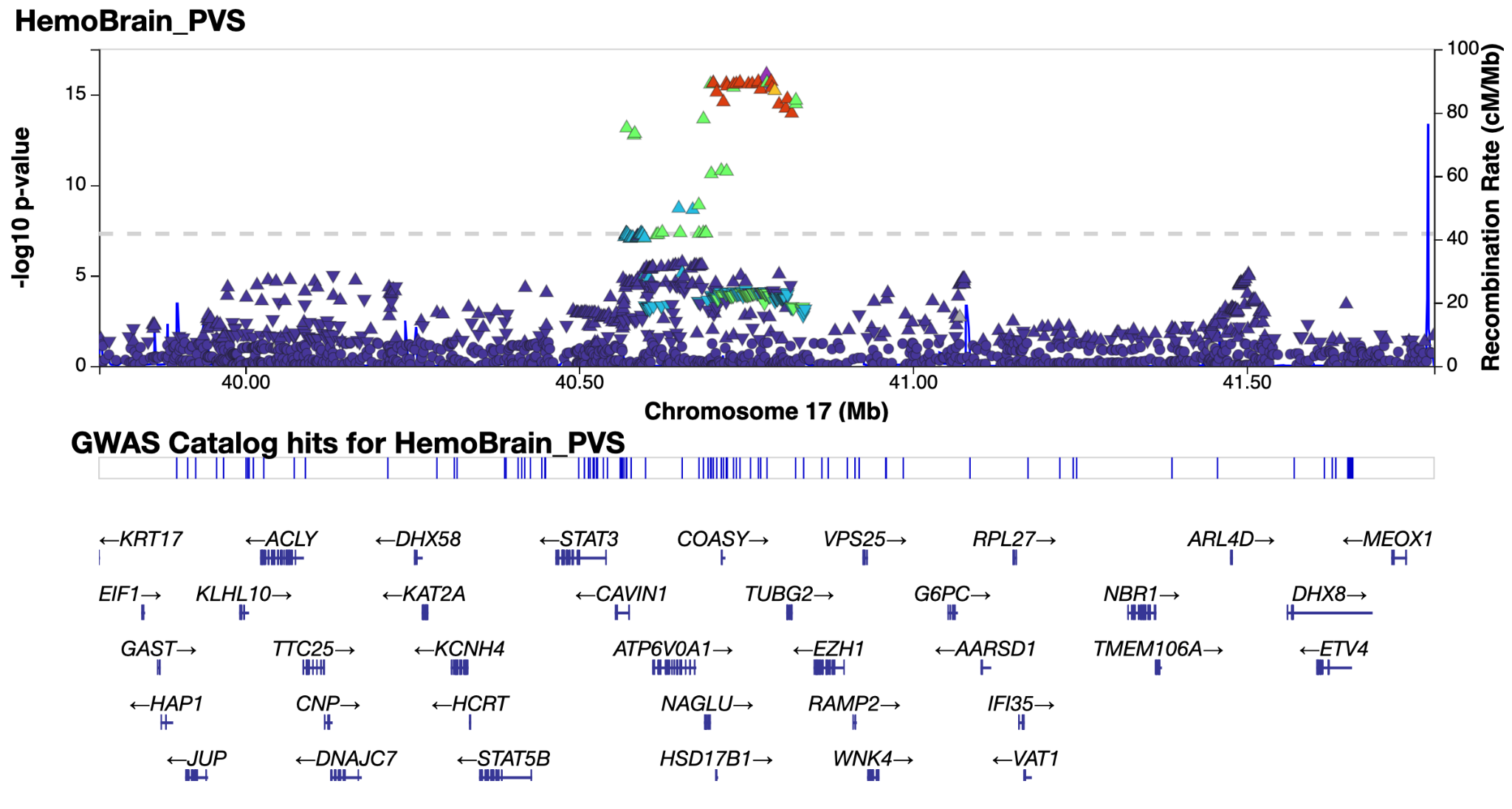

*Supplementary Figure 4.35 & 4.36: LocusZoom**^38^* *plot of genomic loci 35 and 36 - see extended data tables for genomic locus numbering.*

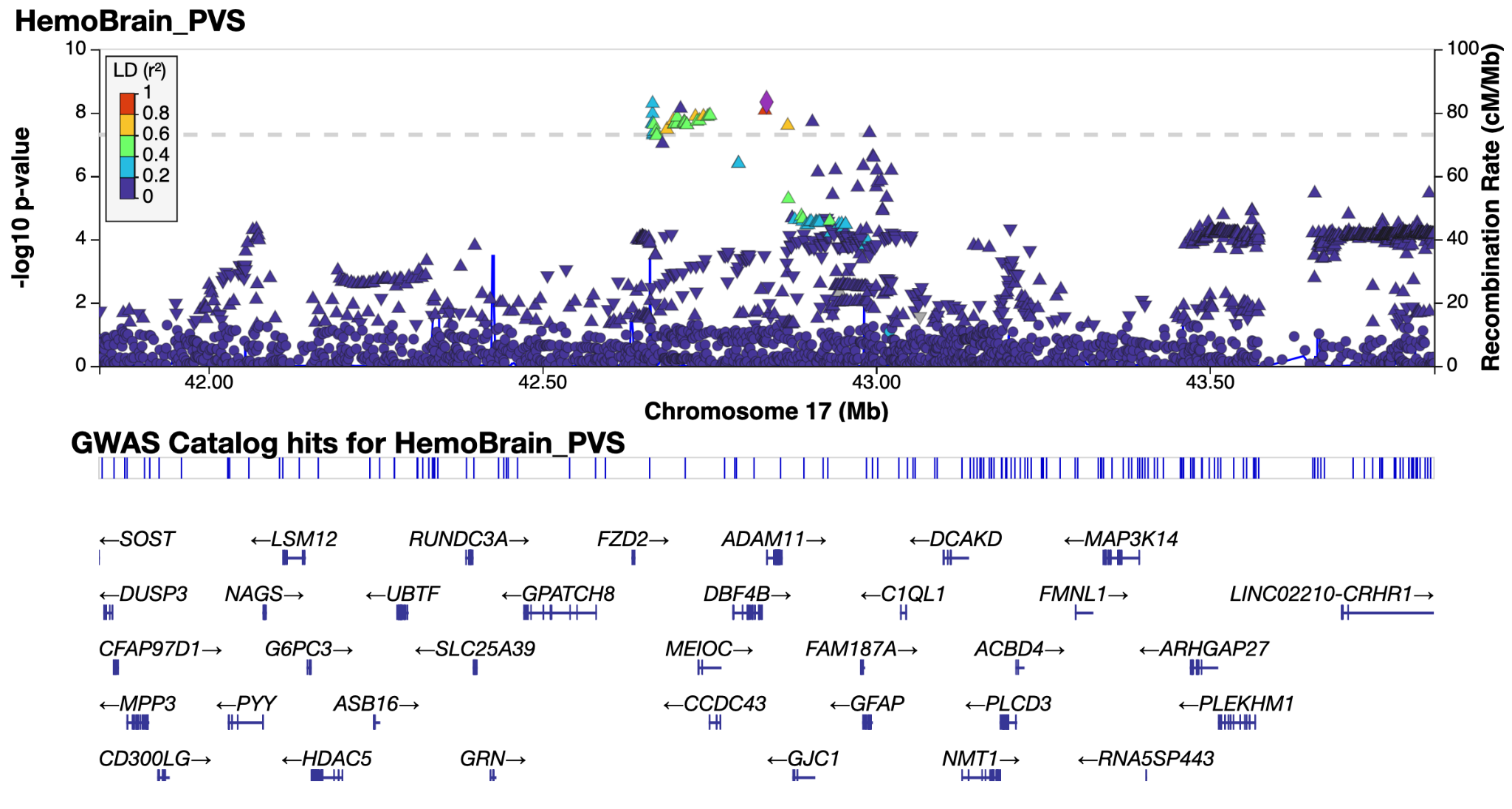

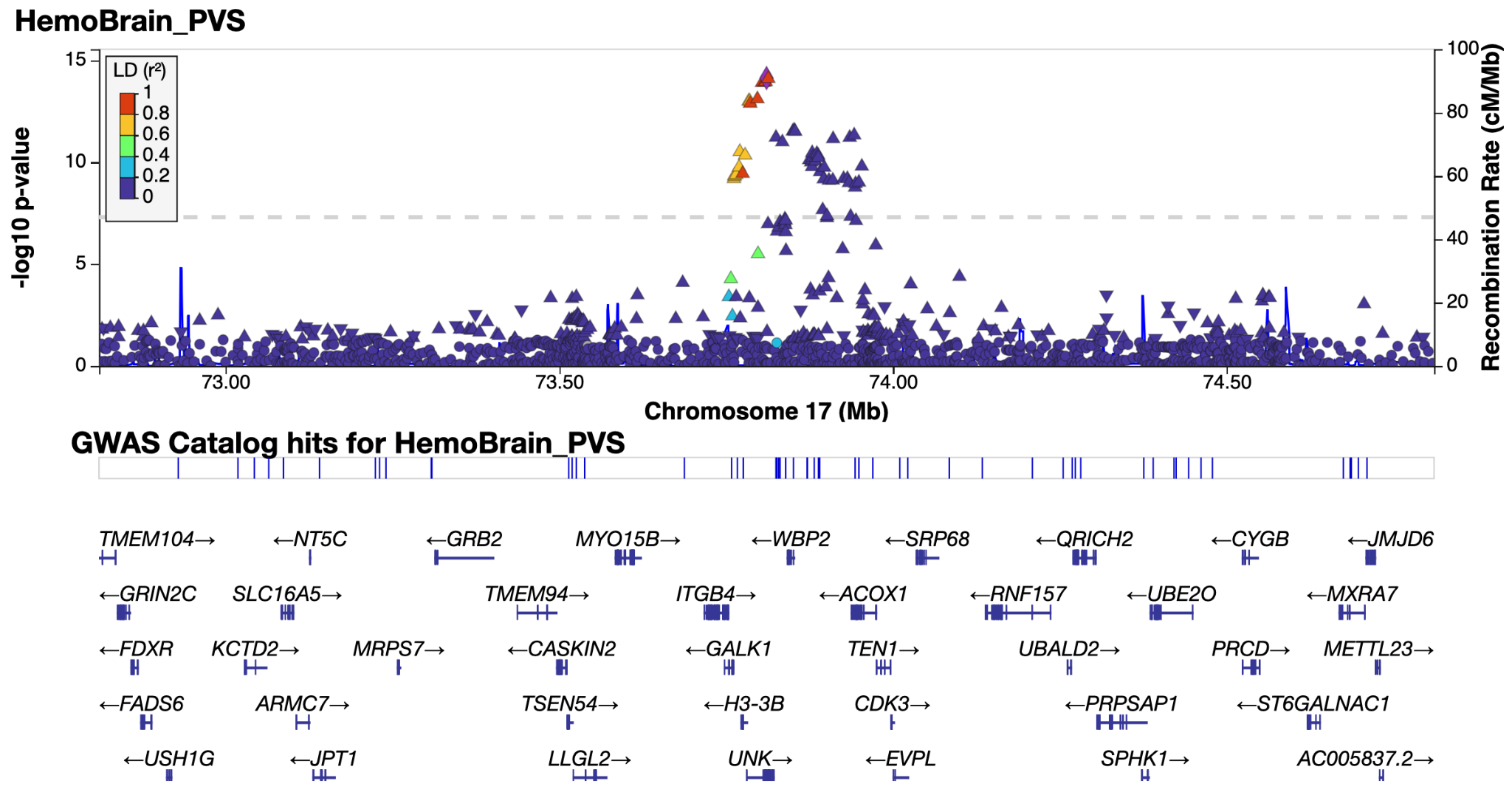

*Supplementary Figure 4.37 & 4.38: LocusZoom**^38^* *plot of genomic loci 37 and 38 - see extended data tables for genomic locus numbering.*

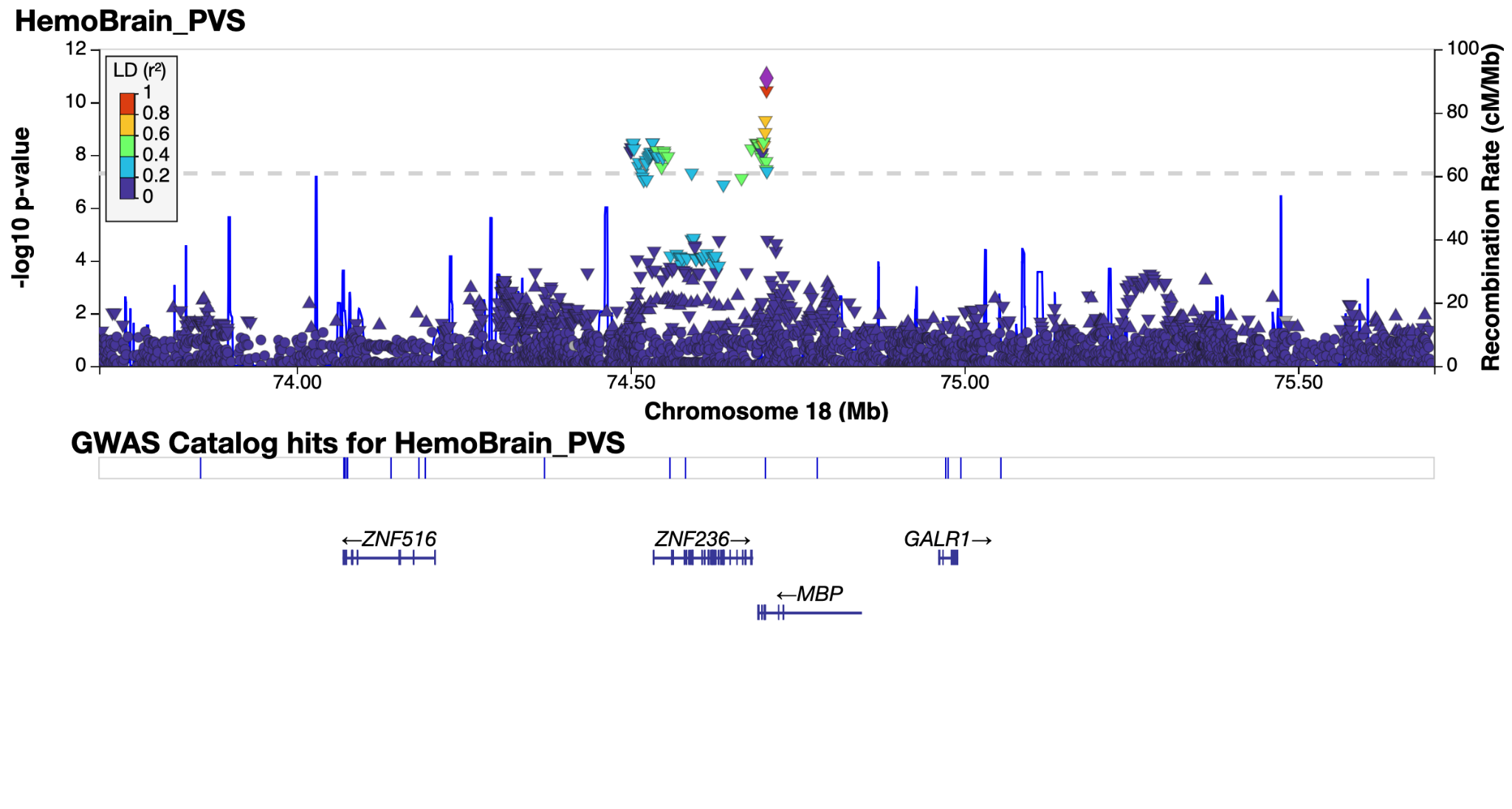

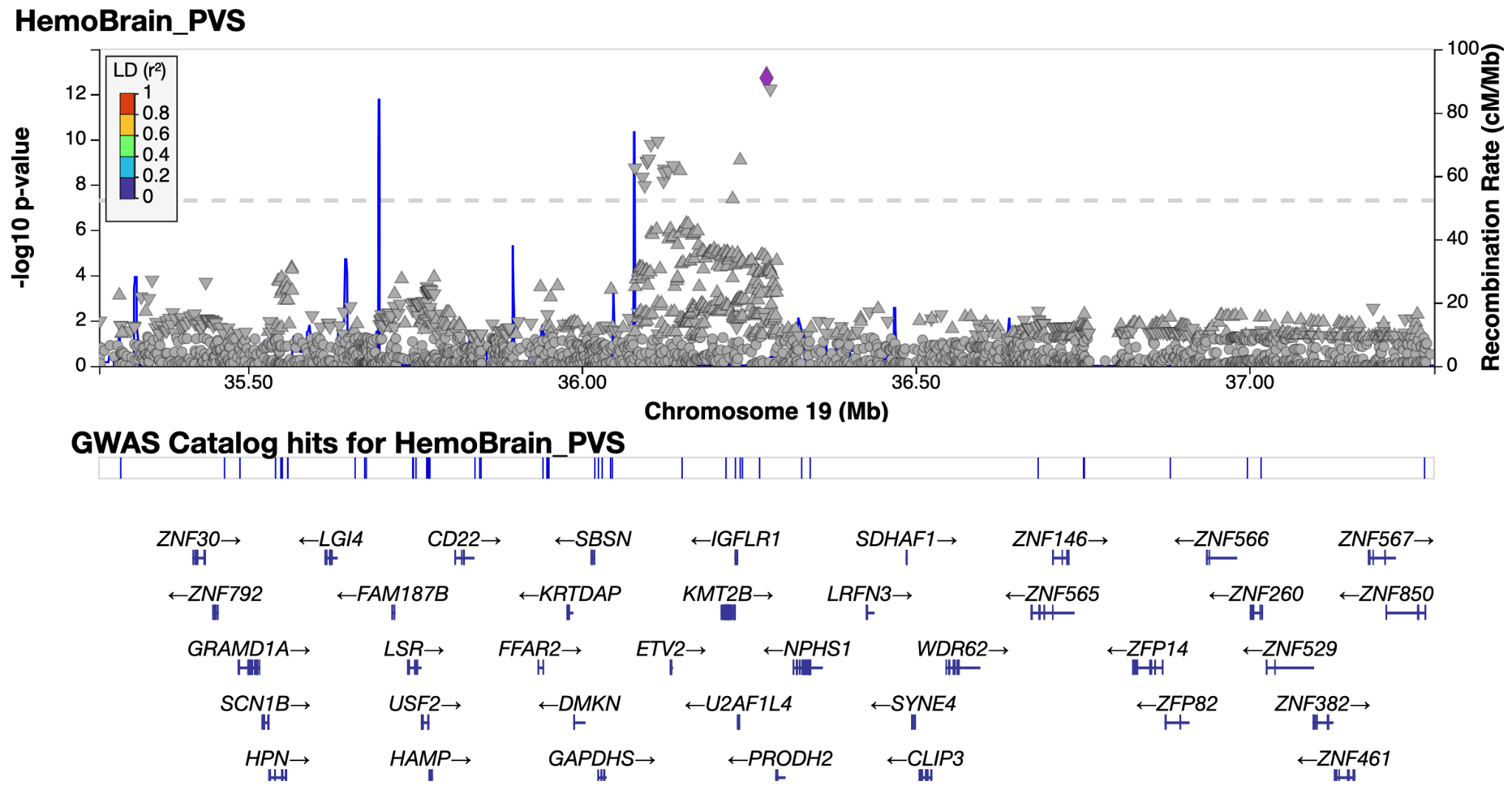

*Supplementary Figure 4.39 & 4.40: LocusZoom**^38^* *plot of genomic loci 39 and 40 - see extended data tables for genomic locus numbering. Non-lead SNP points are in gray as lead SNP is not in LD reference panel (1k genomes).*

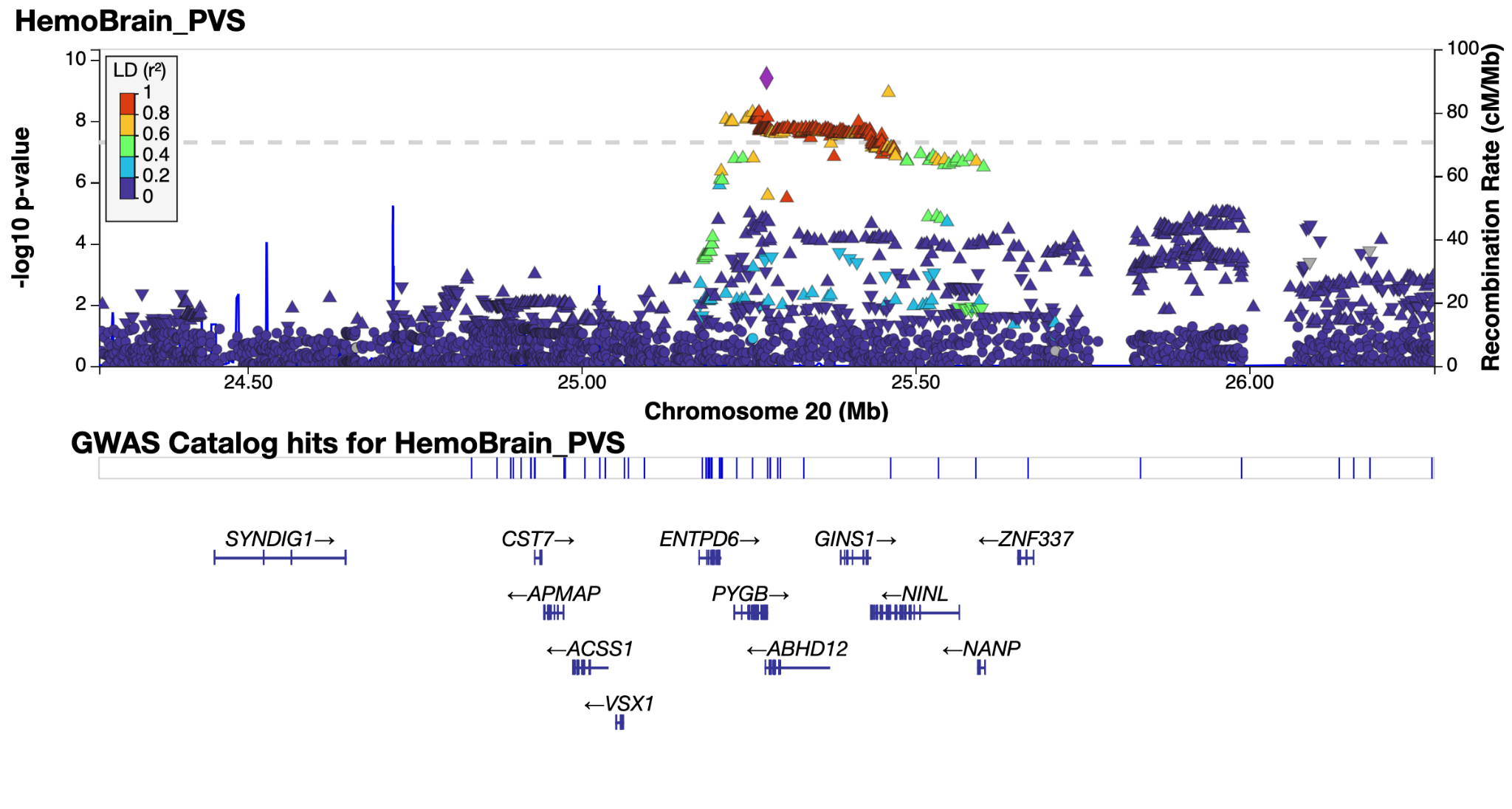

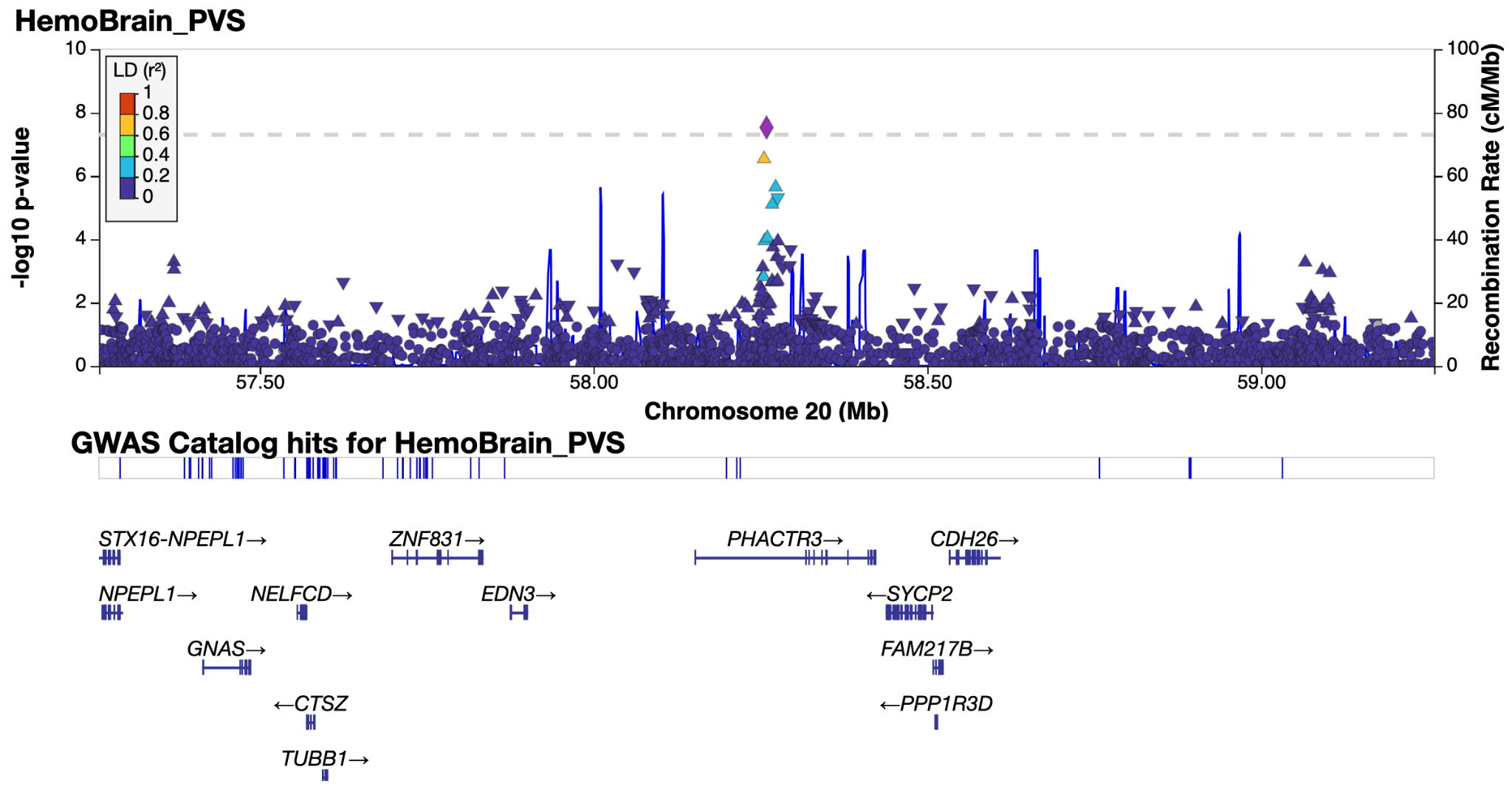

*Supplementary Figure 4.41 & 4.42. LocusZoom**^38^* *plot of genomic loci 41 and 42 - see extended data tables for genomic locus numbering.*

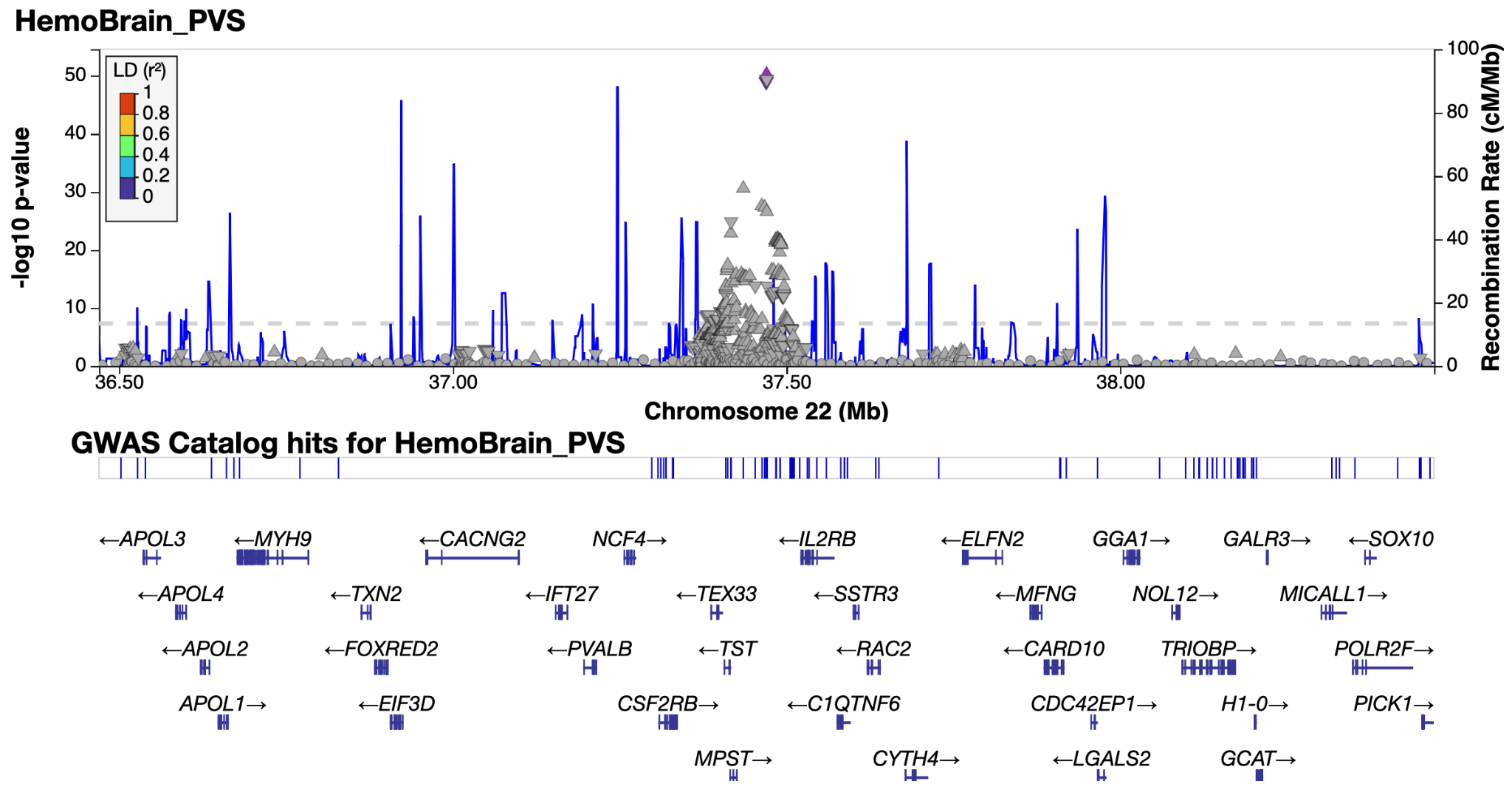

*Supplementary Figure 4.43: LocusZoom**^38^* *plot of genomic locus 43- see extended data tables for genomic locus numbering. Non-lead SNP points are in gray as lead SNP is not in LD reference panel (1k genomes).*

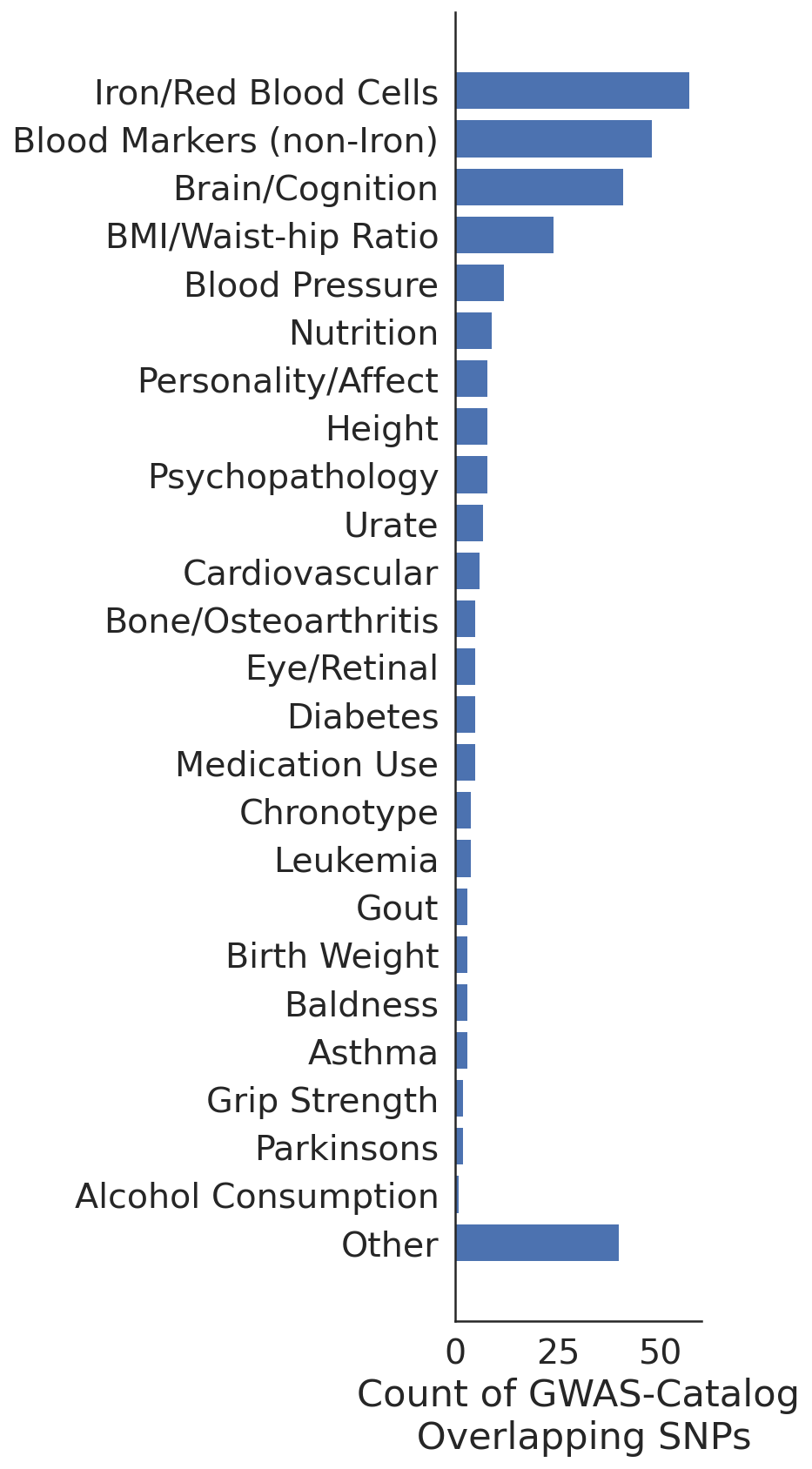

*Supplementary Figure 5: Overlap of independent significant SNPs (r_LD_<0.6) for GWAS of PVS with previously reported SNPs in GWAS-Catalog (e104_r2021-09-15).*

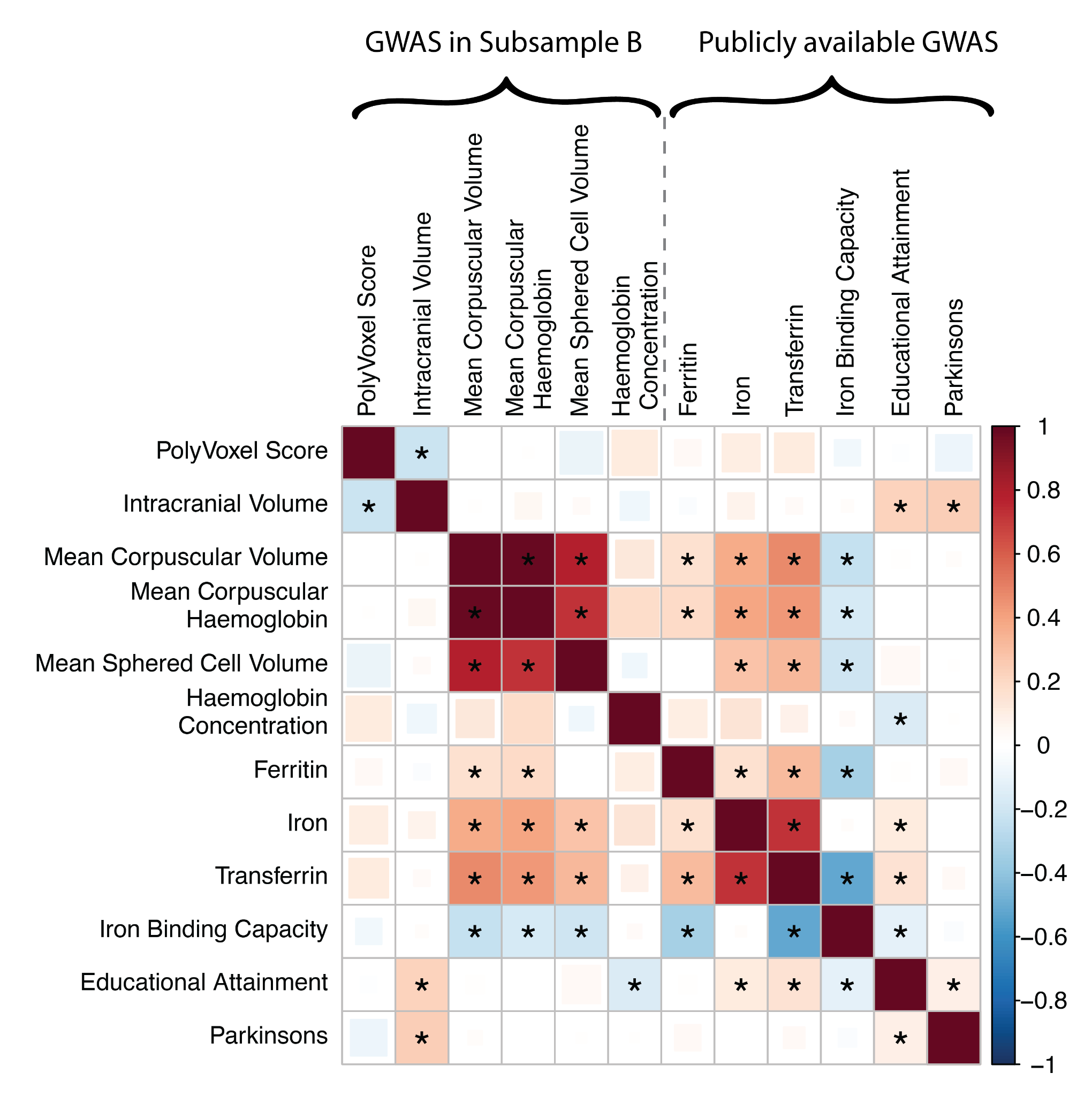

*Supplementary Figure 6: Genetic correlation using LDSC between PolyVoxel Score (PVS) and traits of interest. Asterisk indicates FDR significant correlations.*

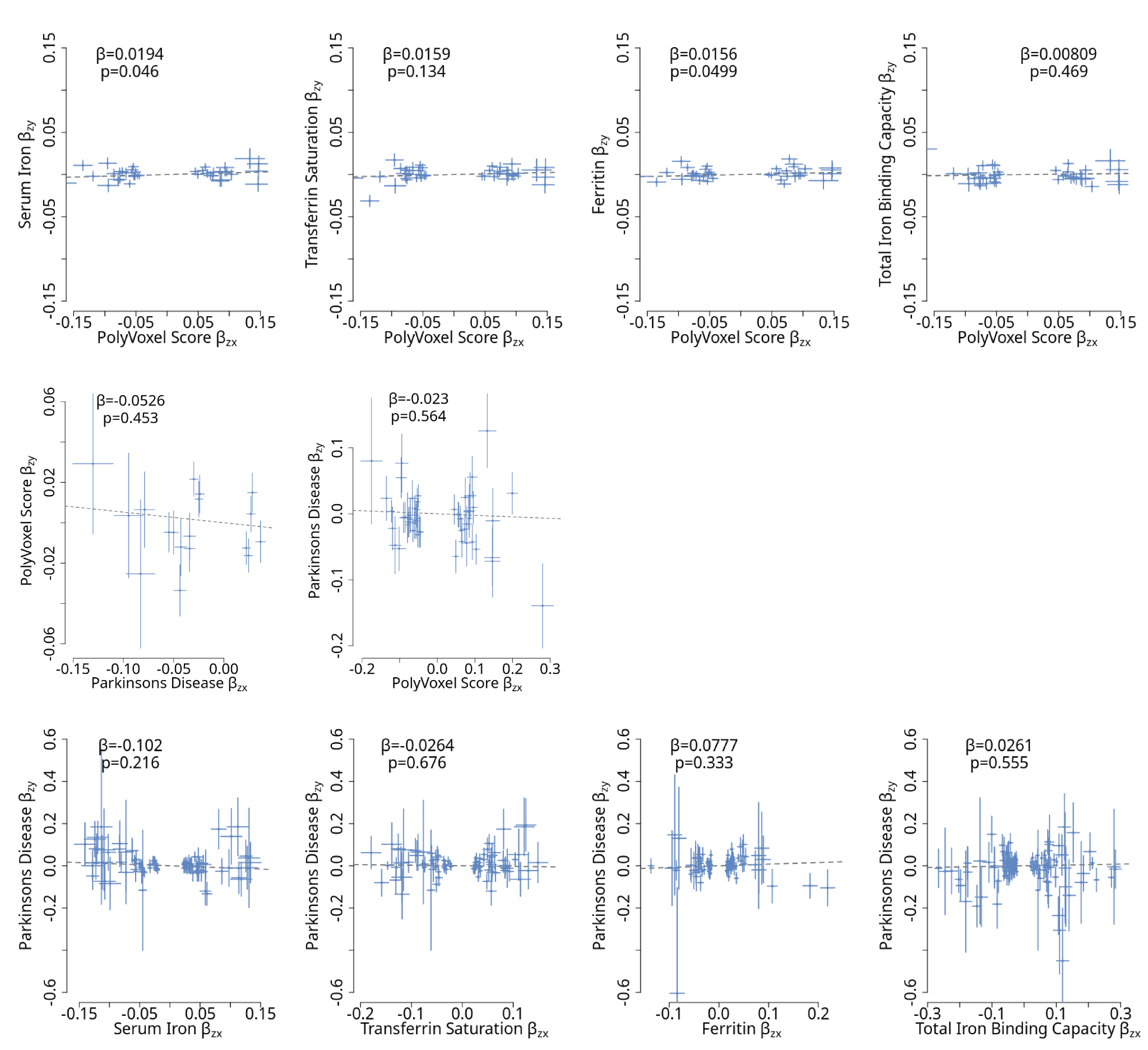

*Supplementary Figure 7: Supplementary mendelian randomization (GSMR) results. For each plot x axis indicates phenotype that is used as exposure (i.e. using variants of that phenotype as instrumental variables) and the y axis indicates the outcome. Top row: reverse direction of results in Figure 3D. Middle row: bidirectional GSMR association between PD and PVS. Lower row: GSMR results treating peripheral blood markers as exposures and PD as outcome.*

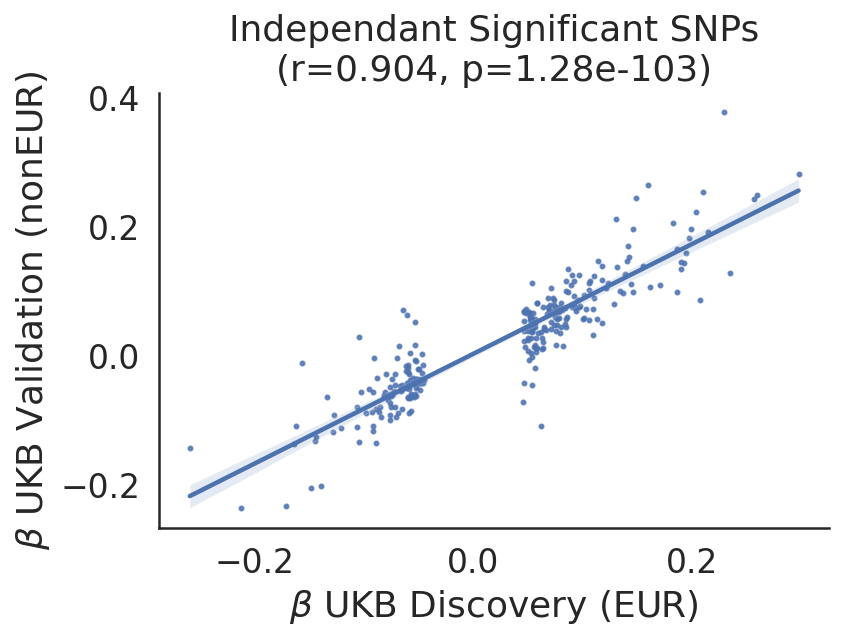

*Supplementary Figure 8: Replication of PVS GWAS discoveries within subsample B of UKB between discovery (30,709 European ancestry individuals) and validation cohorts (4,608 non European ancestry individuals)*

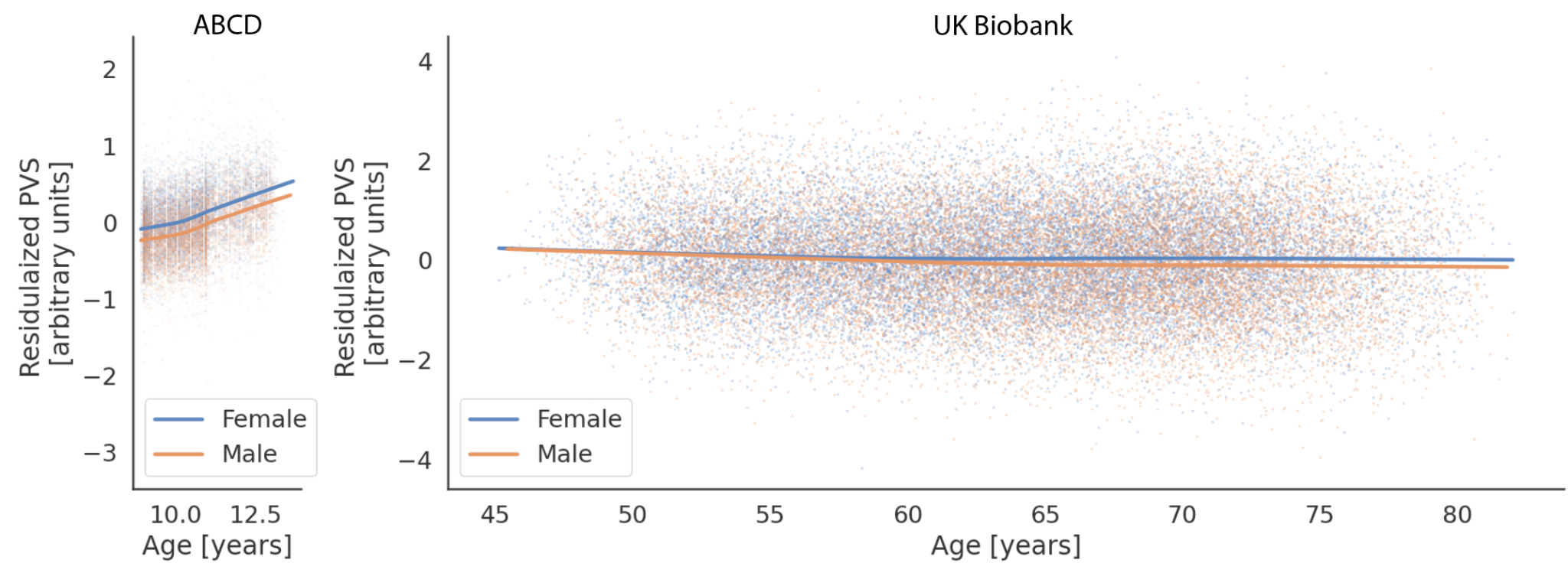

*Supplementary Figure 9: Effect of age on PVS for ABCD (left) and UK Biobank (right). PVS are residualized (see methods for covariates), within each sample so y axes are on different scales - however x axes within each plot have been scaled to match. Higher values on the y axis are in the direction of higher brain iron values.*

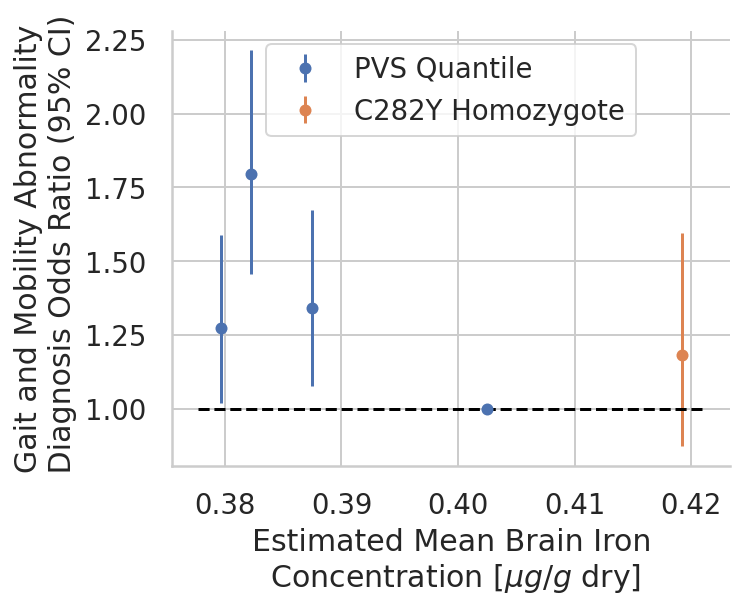

*Supplementary Figure 10: PVS quantile weighted regression to predict Abnormalities of Gait and Mobility [ICD10:R26] in Subsample C, each point represents a categorical factor indicating one of four PVS quantile (blue) or C282Y homozygosity (orange). IPW of 3.63 was used for PD cases in imaging sample (blue) - see methods. Regression (y-axis) was performed using PVS from T2-W, x-axis is an estimate of mean brain iron concentration using T2* imaging for each group.*
